# The fungal intestinal microbiota predict the development of bronchopulmonary dysplasia in very low birthweight newborns

**DOI:** 10.1101/2023.05.29.23290625

**Authors:** Kent A. Willis, Mary Silverberg, Isaac Martin, Ahmed Abdelgawad, Kosuke Tanaka, Ibrahim Karabayir, Brian A. Halloran, Erin D. Myers, Jay P. Desai, Catrina T. White, Charitharth V. Lal, Namasivayam Ambalavanan, Brian M. Peters, Viral G. Jain, Oguz Akbilgic, Laura Tipton, Tamás Jilling, Stephania A. Cormier, Joseph F. Pierre, Ajay J. Talati

**Affiliations:** Division of Neonatology, Department of Pediatrics, Heersink School of Medicine, The University of Birmingham, Birmingham, AL, USA; Microbiome Center, The University of Alabama at Birmingham, Birmingham, AL, USA; Division of Cardiology, Department of Medicine, Wake Forest University School of Medicine, Wake Forest University, Winston-Salem, NC, USA; College of Medicine, The University of Tennessee Health Science Center, Memphis, TN, USA; Division of Neonatology, Department of Pediatrics, College of Medicine, The University of Tennessee Health Science Center, Memphis, TN, USA; Department of Clinical Pharmacy and Translational Science, College of Pharmacy, The University of Tennessee Health Science Center, Memphis, TN, USA; Epidemiological Cardiology Research Center, Wake Forest University, Winston-Salem, NC, USA; Department of Data Science, Analytics, and Visualization, Chaminade University of Honolulu, Honolulu, HI, USA; Department of Biological Sciences, Louisiana State University, Baton Rouge, LA, USA; Pennington Biomedical Research Center, Baton Rouge, LA, USA; Department of Nutritional Sciences, College of Agricultural and Life Sciences, The University of Wisconsin – Madison, Madison, WI, USA; Department of Obstetrics and Gynecology, College of Medicine, The University of Tennessee Health Science Center, Memphis, TN, USA

## Abstract

**Rationale:** Bronchopulmonary dysplasia (BPD) is the most common morbidity affecting very preterm infants. Gut fungal and bacterial microbial communities contribute to multiple lung diseases and may influence BPD pathogenesis.

**Methods:** We performed a prospective, observational cohort study comparing the multikingdom fecal microbiota of 144 preterm infants with or without moderate to severe BPD by sequencing the bacterial 16S and fungal ITS2 ribosomal RNA gene. To address the potential causative relationship between gut dysbiosis and BPD, we used fecal microbiota transplant in an antibiotic-pseudohumanized mouse model. Comparisons were made using RNA sequencing, confocal microscopy, lung morphometry, and oscillometry.

**Results:** We analyzed 102 fecal microbiome samples collected during the second week of life. Infants who later developed BPD showed an obvious fungal dysbiosis as compared to infants without BPD (NoBPD, *p* = 0.0398, permutational multivariate ANOVA). Instead of fungal communities dominated by *Candida* and *Saccharomyces*, the microbiota of infants who developed BPD were characterized by a greater diversity of rarer fungi in less interconnected community architectures. On successful colonization, the gut microbiota from infants with BPD augmented lung injury in the offspring of recipient animals. We identified alterations in the murine intestinal microbiome and transcriptome associated with augmented lung injury.

**Conclusions:** The gut fungal microbiome of infants who will develop BPD is dysbiotic and may contribute to disease pathogenesis.Conclusions: The gut fungal microbiome of infants who will develop BPD is dysbiotic and may contribute to disease pathogenesis.

## Introduction

Bronchopulmonary dysplasia (BPD) is the most common morbidity of prematurity [1, 2]. Disproportionately affecting the most premature newborns, BPD exacts a devastating toll on the immature lung, impairing alveolarization and disrupting vascularization [3]. Despite the considerable, life-long morbidity and staggeringly high mortality of BPD [1, 2], meaningful interventions to modulate lung injury in preterm newborns remain beyond reach [4]. While environmental triggers, such as exposure to hyperoxia, have been described, modulators of the disease remain elusive.

The gut-lung axis is a recently described phenomenon whereby intestinal commensal microbes may project their influence on lung physiology and pathology [5, 6]. Examples of this connection are described in models of neonatal pneumonia [7, 8] and asthma [9, 10]. Intestinal microbes can influence the lung directly via the production of metabolites and indirectly by informing immune mechanisms that module lung injury and recovery [7, 9]. While previous work has focused on bacteria, other commensal organisms, such as fungi, may prove to be critical stimulators of immunity [9, 11]. In early life, gut fungal communities (the mycobiome) appear causally linked to pulmonary immune development [9]. This may make the mycobiome an important modulator of lung injury in BPD. However, the composition of fungal communities in BPD has not been explored.

We performed a prospective, observational cohort study of very low birthweight (VLBW) preterm newborns to determine if the gut mycobiota composition predicts BPD development. The principal outcome was moderate to severe BPD or death before BPD adjudication. To interrogate causality, we also performed fecal microbiota transplant (FMT) into antibiotic-pseudohumanized mice. Due to their importance to immune development, we hypothesized that the composition of the mycobiome would predict BPD in VLBW infants and that this effect would be transferable to *in vivo* models of BPD. Here, we show the gut-lung axis may influence the development of BPD and that dysbiotic gut mycobiota can augment BPD-like illness via FMT.

## Methods

*Detailed methods are provided in the online supplement*.

### Study Subjects

This study was a prospective observational cohort study of inborn, VLBW (< 1500g) preterm newborns without known congenital anomalies or immunodeficiencies conducted at a Level IIIB academic neonatal ICU (MiBPD study: NCT03229967). We defined BPD by the NICHD Workshop definition [12]. For dichotomous analyses, ‘BPD’ represents the composite outcome of Grade II to III BPD or death before 36 weeks of gestation, with the infants without BPD defined as ‘NoBPD’ [13]. Because critically ill premature infants often do not produce stool or only pass meconium during the first week of life, this study utilized the first non-transitional stool produced during the second week of life. Infants that either did not stool or only passed a meconium stool during the second week of life were excluded from this analysis. At the time of this study, probiotics were not administered in this unit. A second cohort from a different Level IV NICU provided the samples for FMT to create antibiotic-pseudohumanized mice.

### Animal experimental protocol

To create pseudo-humanized mice with a BPD or NoBPD-associated microbiome engrafted throughout the critical period for BPD development [14–16], we purchased conventional specific pathogen-free (SPF) C57Bl6/J mice from the Jackson Laboratory. We randomized them into cages of four female mice each. After acclimatization, we exposed the mice to a week of systemically absorbable antibiotics (penicillin VK, cefoperazone sodium, and clindamycin sodium 1g/L), followed by gastric gavage of 200 uL of polyethylene glycol 3350 at 425 g/L, then another week of non-absorbable antibiotics (ertapenem sodium, vancomycin sodium, and neomycin sodium 1g/L). With the native microbiota thus disrupted, we engrafted human microbiota from infants with BPD or NoBPD by FMT. We bred these mice to generate neonatal offspring we termed BPD-FMT or NoBPD-FMT. Pooled litters of neonates with the same exposure were then randomized to either normoxia (FiO_2_ 0.21) or hyperoxia (FiO_2_ 0.85) exposure from postnatal day (P) 3-14 using a standardized model of BPD [17, 18].

### Bacterial and fungal metagenomic sequencing

For the human cohorts, the first non-transitional stool produced during the second week of life was collected in oxygen-reduced PBS with a sterile spatula immediately following evacuation into a clean diaper. For mouse samples, we collected the whole left lung and terminal ileum [19]. Microbial DNA was extracted, amplified, and sequenced as described [20, 21]. Negative and positive controls were collected and passed through the DNA extraction and sequencing steps. Sequence data were processed with *QIIME 2* [22] and *DADA2* [23], decontaminated using *PERFect* [24], and analyzed in R. Details regarding data processing and ecological analyses are available in the online supplement.

### Statistical analysis

Statistical analyses were performed in SPSS (IBM), Prism 9 (GraphPad), Python, or R. Animal experiments are representative of 2-3 pooled mouse litters per human donor with at least 3 human donors per experimental group and analyzed by two-way ANOVA with a Šidák correction.

## Results

We performed a prospective, observational cohort study of 144 VLBW infants in neonatal intensive care. 102 infants produced their first non-meconium stool during the second week of life, from which we characterized the intestinal microbiota. Cohort characteristics are described in table 1. One infant was excluded due to inadequate sequencing quality, and one for incomplete clinical data. The remaining 42 infants either did not stool or only passed a meconium stool during this period and were excluded. Characteristics of excluded infants, which did not differ from the study cohort, are in table S2.

**Table 1.** Cohort Clinical Characteristics and Dwmograohics.

| Table 1. Cohort Clinical Characteristics and Demographics |  |  |  |
| --- | --- | --- | --- |
|  | PPRD | BPD | P Value |
| *n* | 64 | 38 |  |
| Gestational age (weeks) | 29.0 (27.6 - 30.8) | 25.7 (25 - 27.8) | < 0.001 |
| Birthweight (g) | 1145 (840 - 1330) | 775 (670 - 860) | < 0.001 |
| Female sex assigned at birth | 37 (57.8%) | 14 (36.8%) | 0.041 |
| Antenatal Steroids (No) | 8 (17%) | 4 (13.3%) | 0.33 |
| Surfactant (None) | 27 (43.5%) | 1 (2.6%) | <0.001 |
| Apgar score (1 min) | 6 (3 - 7) | 4 (2 - 5) | < 0.001 |
| Apgar score (5 min) | 7 (6 - 8) | 6 (5 - 7) | < 0.001 |
| Small for gestational age* | 6 (9.4%) | 8 (21.1%) | 0.098 |
| Cesarean section | 28 (59.6%) | 20 (66.7%) | 0.531 |
| Prolonged rupture of membranes‡ | 7 (15%) | 5 (16.7%) | 0.834 |
| Critical Risk Index for Babies II (CRIB II) score | 7 (4 - 9) | 12 (9 - 14) | < 0.001 |
| Antenatal antibiotic exposure | 22 (46.8%) | 15 (50%) | 0.785 |
| Duration of antenatal antibiotic exposure (d) | 1 (0 - 1) | 0 (0 - 1) | 0.542 |
| Postnatal antibiotic exposure | 55 (85.9%) | 38 (100%) | 0.015 |
| Duration of postnatal antibiotic exposure (d) | 2 (1 - 2) | 2 (1 - 4) | 0.016 |
| Respiratory Support on Day 3 |  |  |  |
| Mechanical ventilation | 20 (37%) | 33 (89.2%) | < 0.001 |
| Fraction of inspired oxygen | 0.21 (0.21 - 0.21) | 0.25 (0.21 - 0.34) | < 0.001 |
| Predicted risk of severe BPD† | 4.40% (1.68 - 10.14%) | 10.70% (8.73 - 14.49%) | < 0.001 |
| Predicted risk of death† | 2.84% (1.36 - 6.30%) | 10.90% (7.69 - 16.00%) | < 0.001 |
| Respiratory Support on Day 7 |  |  |  |
| Mechanical ventilation | 15 (28.3%) | 30 (83.4%) | < 0.001 |
| Fraction of inspired oxygen | 0.21 (0.21 - 0.23) | 0.29 (0.24 - 0.30) | < 0.001 |
| Predicted risk of severe BPD† | 4.35% (1.54 - 9.40%) | 11.09% (8.22 - 21.50%) | < 0.001 |
| Predicted risk of death† | 3.35% (1.1 - 5.52%) | 10.21% (7.01 - 13.2%) | < 0.001 |
| Endotracheal intubation | 41 (66.1%) | 37 (100%) | < 0.001 |
| Ventilation duration (d) | 1 (0 - 4) | 34 (22 - 50) | < 0.001 |
| Non-invasive ventilation duration (d) | 5 (1 - 17) | 14 (1 - 38) | 0.053 |
| Oxygen exposure duration (d) | 13 (4 - 41) | 92 (30 - 119) | < 0.001 |
| Supplemental oxygen on day 28 | 23 (37.1%) | 33 (91.7%) | <0.001 |
| Death | 0 (0%) | 6 (15.8%) | < 0.001 |
| Data are presented as median (interquartile range) or *n* (%) with significance testing by Mann-Whitney U or Pearson chi-squared test, as appropriate. BPD, Bronchopulmonary dysplasia; PPRD, Post-prematurity respiratory disease. * < 3rd percentile, ‡ > 18 h, † NICHD Neonatal Outcomes Estimator |  |  |  |

### Dysbiosis of the gut microbiota in infants with BPD

We first compared the intestinal microbiota of preterm infants who later developed BPD to infants who did not develop BPD (NoBPD cohort). As shown in figure 1, the fungal intestinal microbiota (mycobiome) of BPD infants differed in community diversity, composition, and interconnectivity from NoBPD infants. We did not identify differences in the bacterial microbiome.

**Figure 1.**
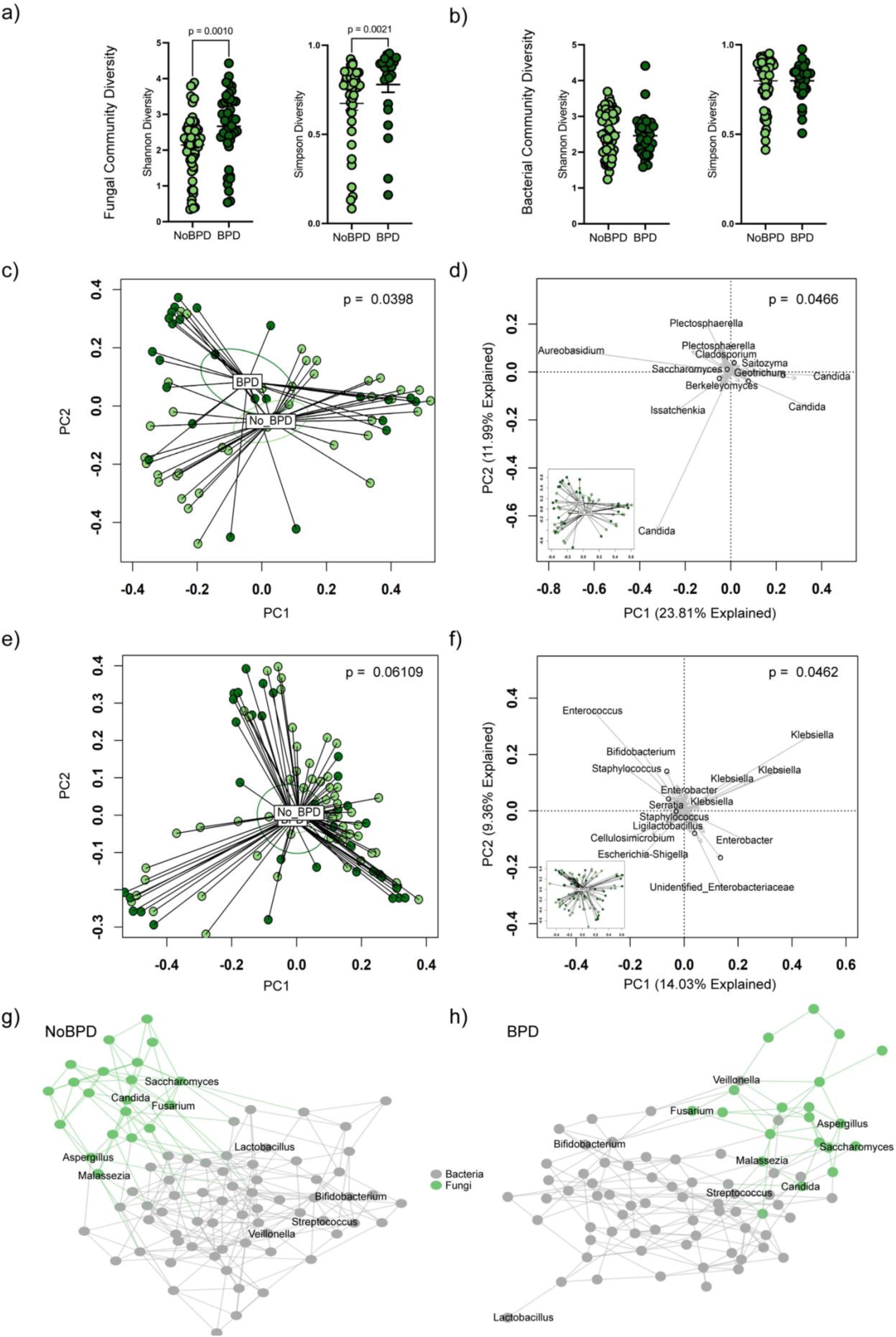
The fungal gut microbiota is altered in infants that will develop bronchopulmonary dysplasia. (*a*) Fungal alpha diversity as quantified by the Shanon and Simpson indices of infants with moderate to severe bronchopulmonary dysplasia or death prior to 36 weeks gestation (BPD) to those without (NoBPD). (*b*) Bacterial alpha diversity. (*c*) Principal coordinates analysis (PCoA) of Bray-Curtis dissimilarity displaying differences in fungal beta diversity (p = 0.0398, permutational multivariate analysis of ANOVA, PERMANOVA. p = 0.4393, permutational multivariate analysis of dispersion, PERMDISP). Ellipses indicate the 95% confidence interval. (*d*) Biplot of principal components analysis (PCA) with fungal community composition driven by amplicon sequence variants (ASVs) aligning to the genera *Candida* and *Aureobasidium* (p = 0.0466). (*e*) PCoA of Bray-Curtis dissimilarity displaying bacterial beta diversity (p = 0.06109, PERMANOVA. p = 0.365, PERMDISP). (*f*) Biplot of PCA of bacterial community composition (p = 0.0462). (*g* and *h*) SPIEC-EASI network analysis shows that BPD infants formed sparser multikingdom networks than NoBPD infants. See also supplemental figures 2 and 3.

For alpha diversity, the within-group variation exceeded the between-group differences for both the mycobiome and the microbiome. The alpha diversity of the mycobiome was greater in BPD as quantified by the Shannon and Simpson diversity indices (*p* = 0.0010 and 0.0021, Mann-Whitney, figure 1a). The alpha diversity of the bacterial microbiome was similar (figure 1b).

We used several complementary methods to quantify beta diversity. As visualized by principal coordinates analysis, disease status produced divergent clustering of the mycobiome (figure 1c). The mycobiome is differently composed in BPD infants (*p* = 0.0209, permutational multivariate ANOVA (PERMANOVA)). We verified this was not a false positive due to unequal dispersion with permutational multivariate analysis of dispersion (PERMDISP, *p* = 0.4393). We performed biplot analysis of principal components analysis graphs to identify which amplicon sequence variants (ASVs) produced these compositional differences. We determined that multiple ASVs belonging to the genera *Candida* and an ASV aligning to *Aureobasidium* made the most significant contributions to the structure of the mycobiome. However, the microbiome was not statistically different (*p* = 0.06109, PERMANOVA, *p* = 0.365, PERMDISP, figure 1d). Biplot analysis showed *Enterococcus* and multiple *Klebsiella* ASVs contributed to the overall bacterial community structure.

To quantify the interconnectivity of these microbial communities, we used a novel cross-domain extension of SPIEC-EASI [11] to build multikingdom interaction networks (figure 1g-h). Compared to NoBPD infants, BPD infants have fewer fungal than bacterial edges but a similar number of nodes (figure S2e). Together, this creates multikingdom interaction networks that are sparser for BPD infants, with fewer fungi-to-fungi and fungi-to-bacterial interactions than NoBPD infants.

We used DESeq2 to identify differentially abundant genera. For the mycobiome, the genus *Candida* was more abundant in NoBPD infants, while *Mortierella* was increased in BPD infants (figure S2). For the bacteriome, we detected increases in the Family Staphylococcaceae and the genus *Veillonella*. Together, these analyses indicate the gut mycobiome of infants that will develop BPD is unique. It was characterized by a lower abundance of *Candida* and a higher number of rarer fungal taxa, including *Mortierella*, arranged in more sparse and less interconnected multikingdom networks.

### Fungal dysbiosis predicts BPD development

To test the predictive potential of the mycobiota, we first developed random forest machine-learning models using the mycobiome or microbiome as an exploratory analysis (figure S4). Next, we developed a LightGBM model using clinical variables known to predict the development of BPD from the Neonatal BPD Outcomes Predictor [25, 26] and higher-order taxa microbiota data (figure 2). After five-fold cross-validation and Bayesian hyperparameter tuning, the model predicted the risk of developing BPD in the unseen validation set with an area under the curve of 0.81 (CI^95%^ 0.61-1.00), with a sensitivity of 80% and specificity of 89%, positive predictive value of 89%, and an F1-score of 0.89 (figure 2). For the composite outcome of severe BPD or death, a second model obtained an area under the curve of 0.89 (CI^95%^ 0.76-1.00), with a sensitivity of 75% and specificity of 86%, a positive predictive value of 90%, and F1-score of 0.82. Intriguingly, in variable importance analysis, several fungal phyla’s relative abundance strongly contributed to the model (figure S4e), including the minor phyla Mortierellomycota and Rozellomycota and the major phyla Ascomycota and Basidiomycota. The fungal taxa outperformed several well-described clinical characteristics and all bacterial phyla except Bacteroidata.

**Figure 2.**
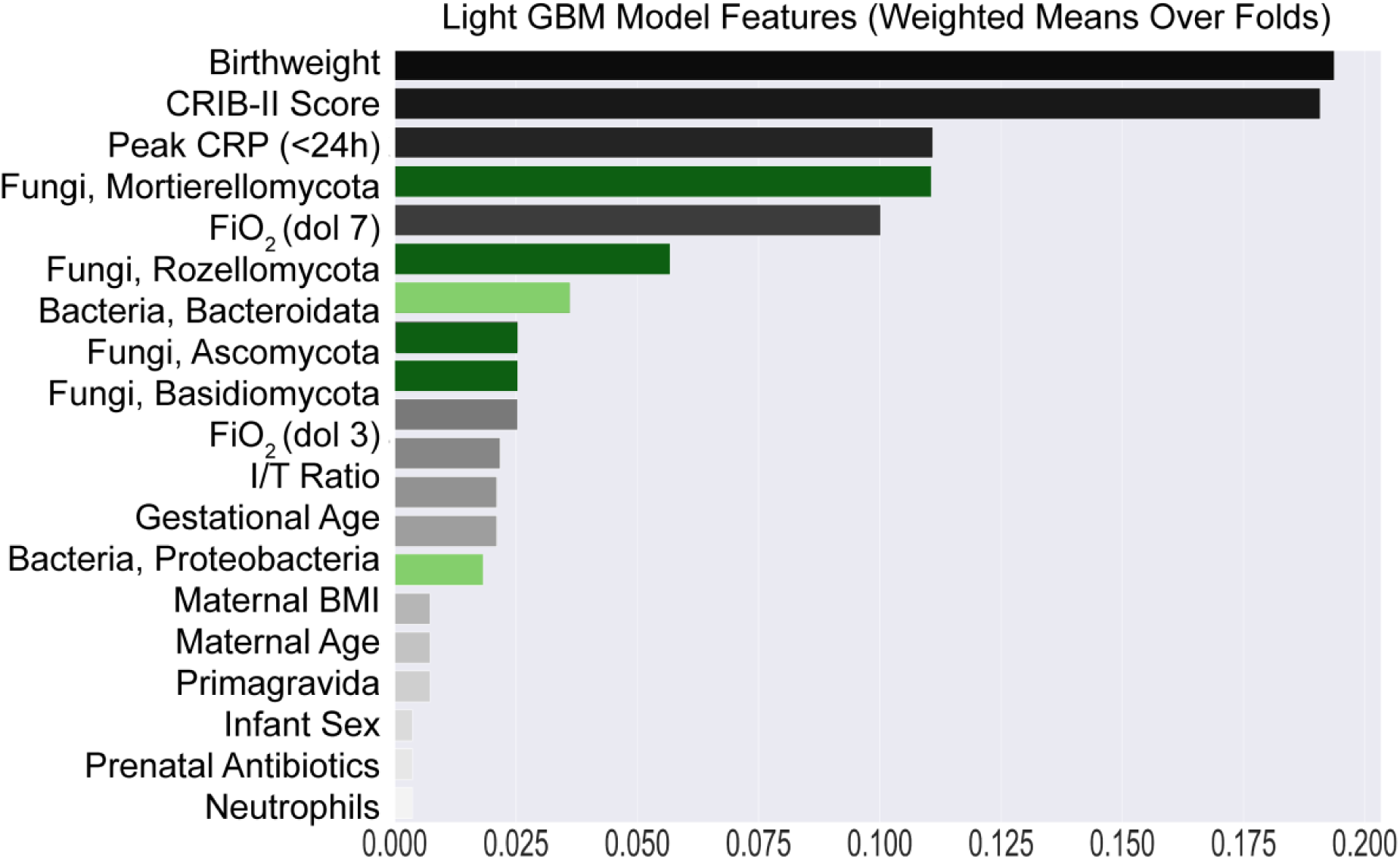
The mycobiota predicts the development of BPD. (*a*) A light gradient-boosted machine learning model (LightGBM) with Bayesian hyperparameter tuning developed from clinical data and phylum-level microbiome data indicates that key members of the mycobiome predict the development of BPD. CRIB-II, critical risk index for babies-II. CRP, c-reactive protein. FiO_2_ (dol 7), average fraction of inspired oxygen on the 7th day of life. FiO_2_ (dol 3), average fraction of inspired oxygen on the 3rd day of life. I/T, immature to total lymphocyte ratio on admission complete blood count. BMI, body mass index.

### Sex-specific differences in the composition of the mycobiome

Due to known sex-specific differences in BPD [27, 28], we predetermined to test if sex-specific features exist in the neonatal mycobiome (figures 3 and S4-5). Shannon and Simpson diversity of the mycobiome and bacteriome did not differ between those assigned female (AFAB) or male at birth (AMAB) figure 3a-b). Fungal community composition differed for all groups (*p* = 0.0407, PERMANOVA; *p* = 0.5957, PERMDISP, figure 3c). AFAB infants with BPD and NoBPD clustered closer than infants AMAB (figure 3c). DESeq2 identified differences in the abundance of the fungal genera *Mortierella* and *Pseudeurotium* in AMAB infants who later developed BPD (figure S6a-b). SPIEC-EASI network analysis identified greater interconnectivity of infants with NoBPD regardless of sex assigned at birth (figure 3e-h, S6e-f). Infants AMAB had fewer fungal edges than infants AFAB, but the number of edges was reduced only in infants AMAB that would develop BPD (figure S6c). These differences produced sparser networks in infants AMAB, with the least interconnected networks in infants AMAB with BPD. From these analyses, we conclude that sex-specific differences in the composition of the mycobiome are the strongest in infants AMAB that will develop BPD.

**Figure 3.**
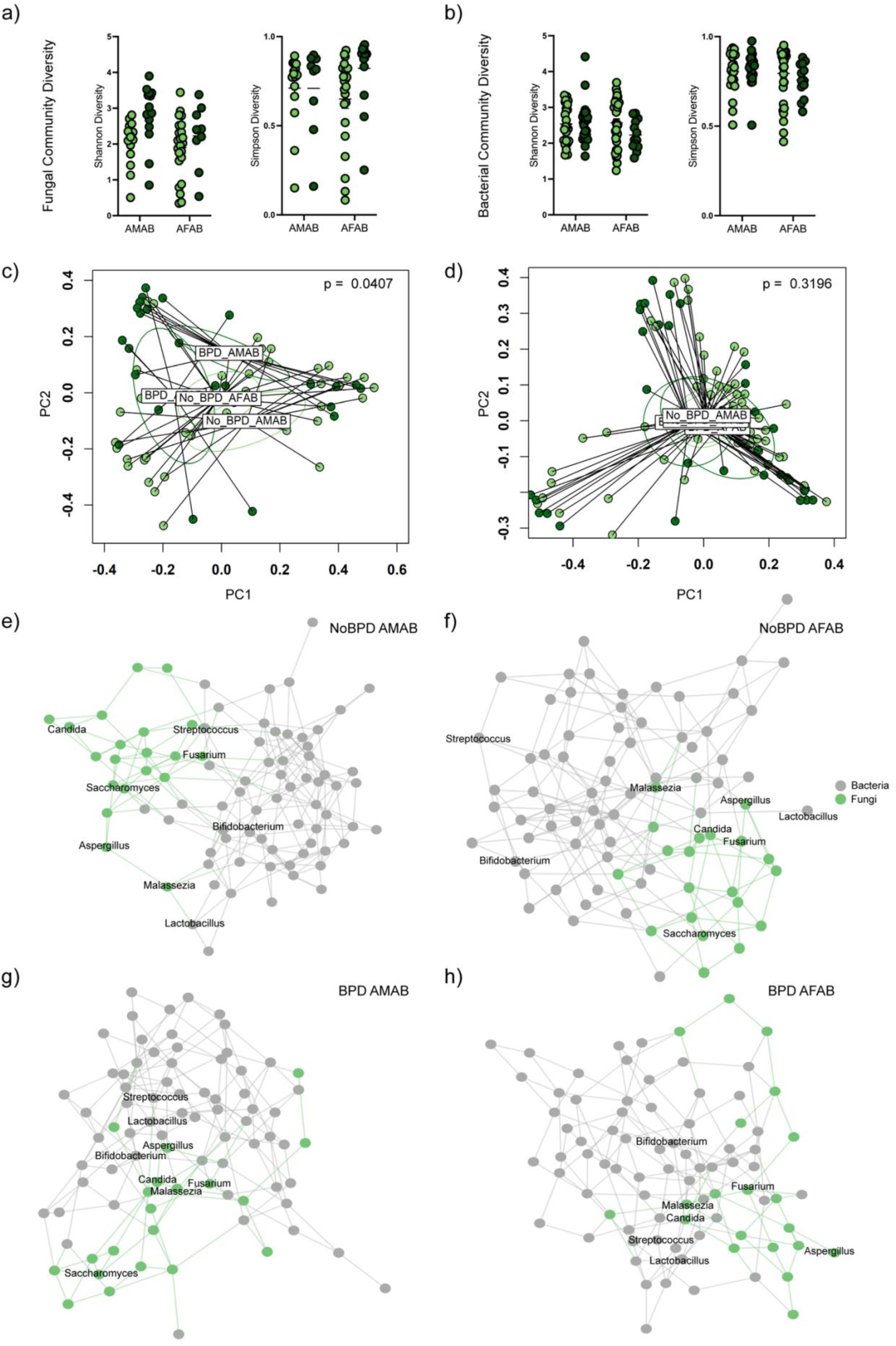
**Sex-specific differences in infants that will develop BPD**. (*a*) Fungal alpha diversity in infants in infants assigned male (AMAB) or female at birth (AFAB) with NoBPD or BPD. (*b*) Bacterial alpha diversity. (*c*) Principal coordinates analysis (PCoA) of Bray-Curtis dissimilarity showing differences in the mycobiome for each sex and disease state (p = 0.0407, PERMANOVA, p = 0.5957, PERMDISP). Ellipses indicate the 95% confidence interval. (*d*) PCoA of Bray-Curtis dissimilarity of the bacterial microbiome (p = 0.3196, PERMANOVA. p = 0.6439). (*e*-*h*) SPIEC-EASI network analysis shows that BPD infants AMAB formed the sparsest networks. See also supplemental figures 5 and 6. Neutrophils, neutrophil count on admission complete blood count. See also supplemental figure 4.

### Fecal microbiota transfer transmits a severe phenotype of BPD

To test if alterations in the gut mycobiome that we found preceded the development of BPD could augment lung injury, we transferred the gut microbiome from infants with either BPD or NoBPD into mice. Because the effects of bacterial and fungal members of the microbiome are interdependent, we transferred intact microbial communities. Following antibiotic preparation, recipient maternal mice were randomly orally inoculated with anoxic fecal samples from donor infants. These mice were bred, and the resulting newborn mice were exposed to hyperoxia to induce BPD-like lung injury (figure 4a).

**Figure 4.**
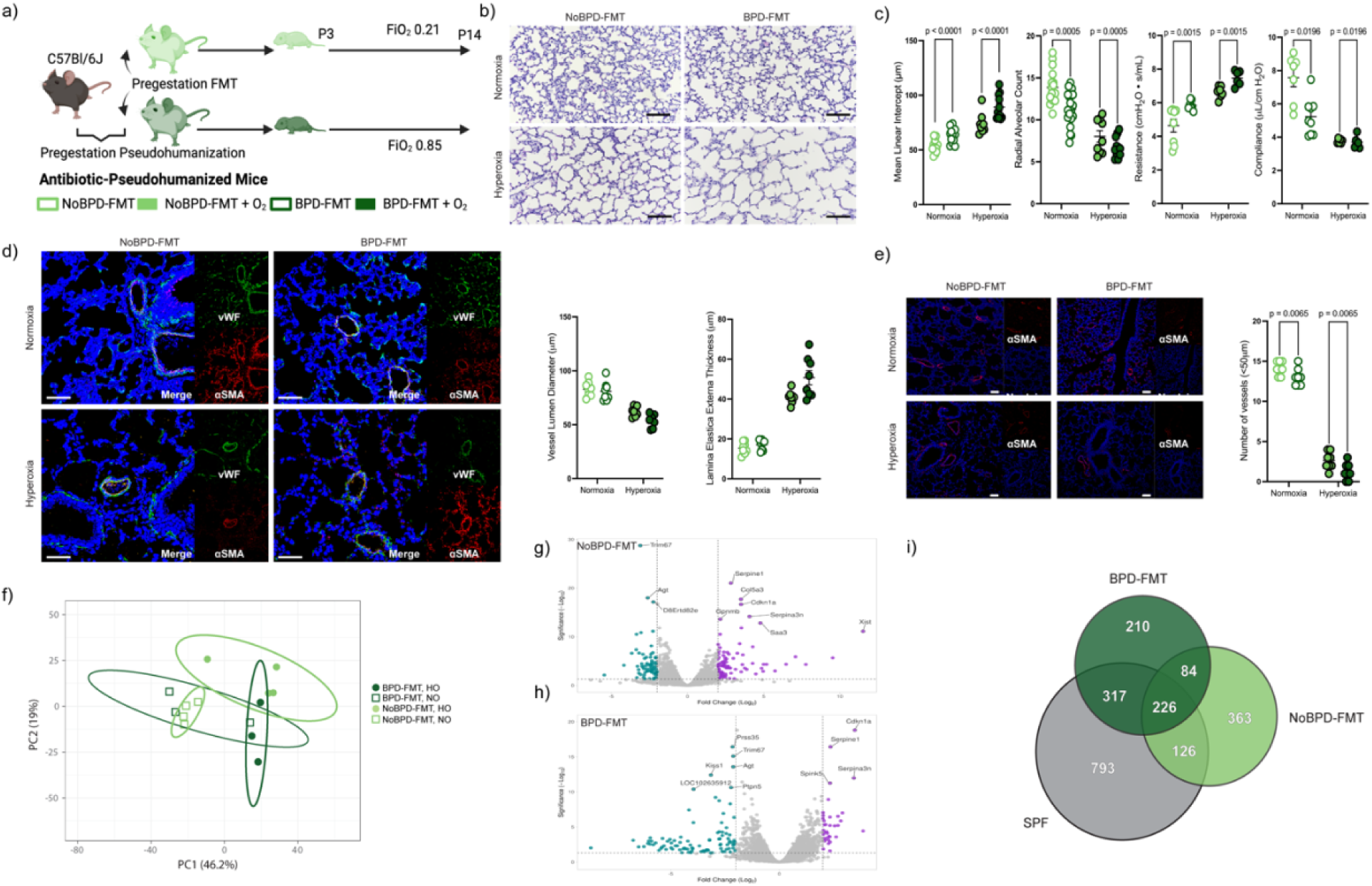
Fecal microbiota transfer from infants with BPD augments lung injury in mice. (*a*) Experimental schematic. (*b*-*d*) Histomorphological analysis and forced oscillometry of the lungs of 14-day-old neonatal mice born to pseudo-humanized mice with fecal microbiota transport from BPD-associated microbiomes (BPD-FMT) or NoBPD-associated microbiomes (NoBPD-FMT). Scale bars, 100 µm. (*d*-*e*) Immunohistochemistry of alpha-smooth muscle actin and von Wilibrand’s Factor showing vascular changes consistent with early pulmonary hypertension (*d*) and reduced vascular density of 20-50 µm vessels (*e*). Scale bars, 20 and 50 µm. (*f*-*i*) Hyperoxia exposure induces different transcriptomic profiles in the lungs in BPD-FMT or NoBPD-FMT mice by RNA-seq. (*f*) Principal components analysis of global transcriptome by both disease and FMT. (*g*) Volcano plot of the hyperoxia-induced transcriptomic alterations in NoBPD-FMT. (*h*) Volcano plot of hyperoxia-induced transcriptomic alterations in BPD-FMT. (*i*) Venn diagram showing unique gene regulation in both BPD-FMT and NoBPD-FMT mice as compared to conventional specific pathogen-free mice (SPF). FiO_2_, fraction of inspired oxygen. Schematic created using BioRender.com. See supplemental figures 12-13 specific differences in the development of BPD detected by single-cell transcriptomics (34, 49–51), this is an interesting finding worth further investigation.

Neonatal mice that received BPD-associated microbiomes (BPD-FMT) had more significant histological lung injury when exposed to hyperoxia than mice that received NoBPD-associated microbiomes (NoBPD-FMT) (Greater mean linear intercept and reduced radial alveolar count, *p* < 0.0001 and *p* = 0.0005, two-way ANOVA, figure 4b-c). Functionally, BPD-FMT mice had increased pulmonary resistance (*p* = 0.0015) and decreased compliance (*p* = 0.0196). We used immunohistochemistry and morphometry to measure neointimal changes consistent with pulmonary hypertension (figure 4d) and identified BPD-FMT mice had reduced vascular density of 20-50 µm diameter vessels (*p* = 0.0065, figure 4e).

We next used RNA-seq to quantify transcriptomic changes in the lungs of these mice. Principal components analysis confirmed unique global transcriptomic changes in the lungs of mice by both FMT and hyperoxia exposure (figure 4f). Volcano plots show a clear difference in the response of NoBPD-FMT (figure 4g) and BPD-FMT (figure 4h) to hyperoxia exposure. We identified unique alterations in lung injury (table S5), lectins and other fungal interacting genes (table S6), and antimicrobial peptides (figure S12, table S7). To identify genes uniquely regulated in mice exposed to FMT, we compared their lung transcriptome to hyperoxia or normoxia-exposed conventional mice (figure 5i). 210 genes were uniquely regulated in BPD-FMT mice (table S8) and 363 genes in NoBPD-FMT mice (table S9) as compared to 793 uniquely regulated genes in the conventional mice. Acute phase response and pathogen-activated cytokine storm signaling was increased BPD-FMT but not in NoBPD-FMT mice by pathway analysis (figure S13). Conventional mice had increased angiogenic, interleukin-6, and -17 signaling that was not observed in either FMT group.

**Figure 5.**
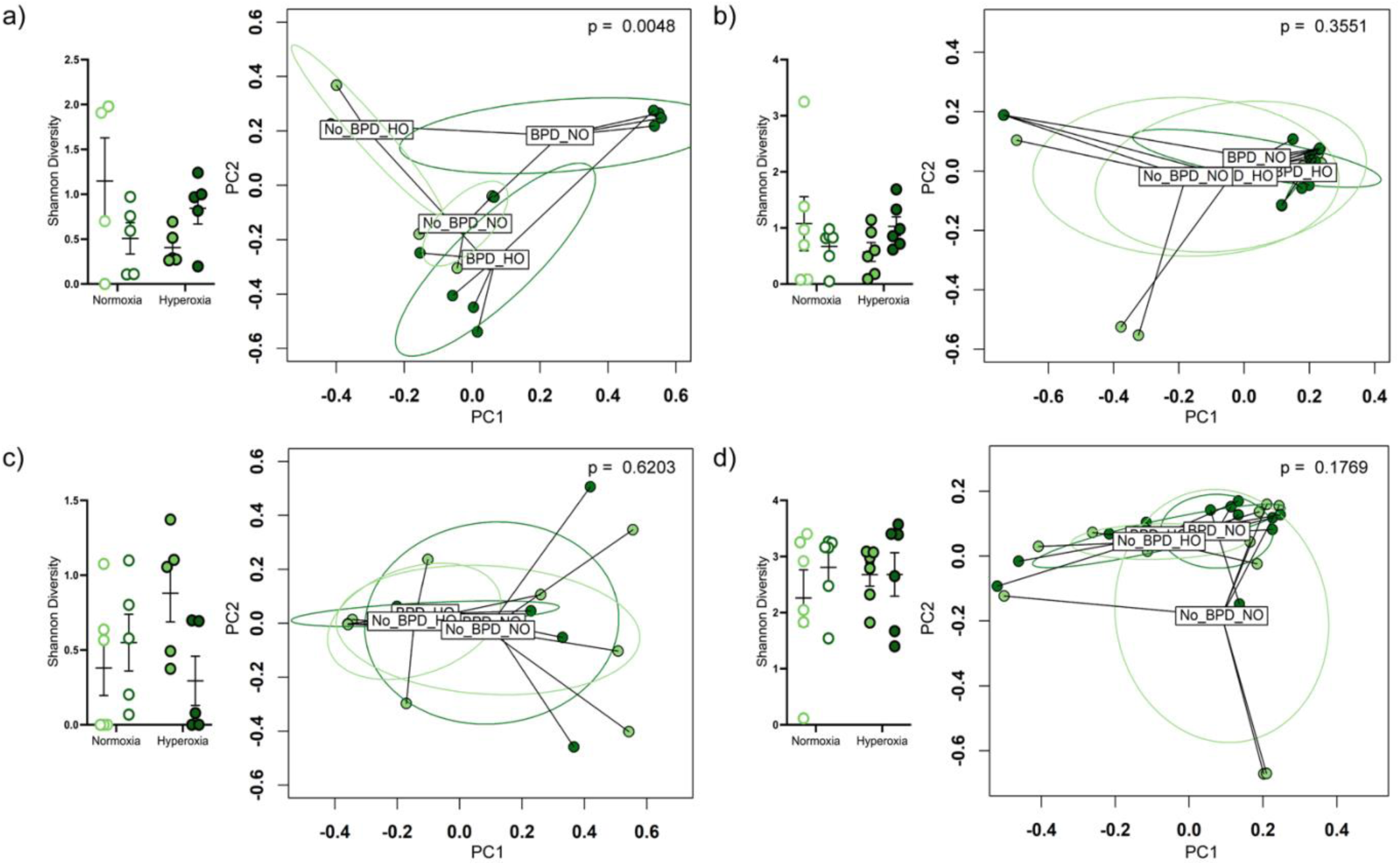
Fecal microbiota transfer alters the lung and gut microbiome of recipient mice. (*a*) Alpha and beta diversity of the gut fungal and (*b*) gut bacterial microbiome in 14-day-old neonatal mice. (*c*) Alpha and beta diversity of the lung microbiome. (*d*) Lung alpha and beta diversity of the bacterial microbiome. BPD_NO, BPD normoxia. BPD_HO, BPD hyperoxia. NoBPD_NO, NoBPD normoxia. NoBPD_HO, BPD hyperoxia. Ellipses indicate the 95% confidence interval.

### Fecal microbiota transfer alters the neonatal mouse microbiome

We also characterized the mouse lung and gut microbiomes to identify how FMT and hyperoxia exposure influence their ecology. In the mouse gut and lung, 79% and 69% of the genera were shared with human donor microbiomes. Beta but not alpha diversity was significantly different for the gut mycobiome (*p* = 0.0062, PERMANOVA figure 5a) and microbiome (*p* = 0.0167, figure 5b). In the lungs, only the microbiome was uniquely composed (*p* = 0.0103, PERMANOVA, figure 5c-d). From these analyses, we conclude that a BPD-associated mycobiome is transferable to neonatal mice and results in augmented lung injury from hyperoxia exposure.

## Discussion

BPD is the most common morbidity and severe chronic respiratory disease in newborns [4]. Considerable heterogeneity exists in its clinical phenotype, and effective means to predict and modulate disease remain unavailable because the underlying etiology remains unclear. Here, we sought new answers in the gut. We found that in preterm neonates who would eventually develop BPD, the fungal intestinal microbiota differed several weeks before diagnosis from neonates who did not develop BPD (NoBPD cohort). Furthermore, these differences were driven by fungal commensals. With FMT, we showed that gut microbiota transferred from the BPD cohort could augment lung injury as compared to microbiota transferred from the NoBPD cohort. Our findings suggest the gut-lung axis may play a crucial role in the etiology of BPD.

Fungi are vital members of the human microbiota [29]. However, unlike bacterial commensals, their non-pathological and non-parasitic functions remain poorly understood [29]. So, while the microbiome has recently emerged as a contributor to clinical heterogeneity in adult patients in intensive care [30–38], our knowledge of fungal commensals in newborn respiratory disease is limited [39]. Here, in one of the most extensive studies of the multikingdom intestinal microbiome in preterm newborns, we show that the composition of gut mycobiome during the second week of life predicts the development of BPD. The abundance of certain higher-order fungal taxa was similarly predictive of the risk of developing BPD as well-established clinical criteria [25, 26, 40]. Intriguingly, we also identified robust sex-specific differences between infants who would develop BPD that were not observed in infants who did not develop BPD. In light of recent sex-specific differences in the development of BPD detected by single-cell transcriptomics [27, 41–43], this is an interesting finding worth further investigation.

Collectively, our analyses found that the composition of the intestinal mycobiota of infants that would develop NoBPD was more uniform, whereas those that eventually developed BPD had more disparate mycobiota. This suggests that a particular pattern of mycobiota development may be necessary to impart resistance to the development of BPD, and failure to do so in various ways is associated with disease development. A more robust health-associated microbiome with a less specific disease-associated microbiome is a well-described ecological pattern termed the Anna Karenina phenomenon [44]. This paradigm holds for other aspects of the pathogenesis of BPD [45]; therefore, this finding fits within our broader understanding of the disease.

A previous study of the bacterial microbiota identified a divergence over time between the bacterial community composition in the gut and lungs of preterm infants that would eventually develop BPD [46], suggesting that a gut-lung axis may influence the development of BPD. While the mechanisms underlying the gut-lung axis require further exploration in BPD, work in other childhood diseases has suggested some potential mechanisms. Mouse models of childhood asthma suggest intestinal fungi are more immunostimulatory than bacteria, while intestinal bacteria are more responsible for producing metabolites detectable in the host’s serum [9]. Gut intestinal fungi, mainly the yeast *Candida albicans*, are known to provoke strong immune responses in the host [47]. Indeed, monocolonization with *Candida* can stimulate postnatal immune development [48]. The greater abundance of *Candida*-dominated communities we observed in NoBPD infants is intriguing in this regard.

Limitations of this study include that we transplanted whole fecal samples in the mouse FMT experiment. While we analyzed the microbiota transferred to the neonatal mice, it remains unclear which components of this complex mix of microbes, metabolites, and nutrients modulate lung injury. Future experiments may benefit from a multiomic approach to identify and validate potential candidate microbes or metabolites. Similarly, it is currently not feasible to confine the transfer of microbes to only one organ system. Quantifying how spillover from FMT might affect other host organs, such as the lungs, remains challenging. A strength of this study was the rigorous exploration of the multikingdom microbiota using the most up-to-date bioinformatic techniques [39] and the demonstration that the mycobiome effect was transferable by FMT.

### Conclusions

In this prospective, observational cohort study of VLBW preterm newborns, baseline compositional differences in the gut mycobiome predicted the eventual development of BPD. Furthermore, we showed that this dysbiotic mycobiota could augment lung injury in a mouse model of BPD. These findings suggest that the gut mycobiota can modulate lung injury and represent a therapeutic target in newborn lung disease.

Collectively, our analyses found that the composition of the intestinal mycobiota of infants that would develop PPRD was more uniform, whereas those that eventually developed BPD had more disparate mycobiota. This suggests that a particular pattern of mycobiota development may be necessary to impart resistance to the development of BPD, and failure to do so in various ways is associated with disease development. A more robust health-associated microbiome with a less specific disease-associated microbiome is a well-described ecological pattern termed the Anna Karenina phenomenon (52). This paradigm holds for other aspects of the pathogenesis of BPD (53); therefore, this finding fits within our broader understanding of the disease.

A previous study of the bacterial microbiota identified a divergence over time between the bacterial community composition in the gut and lungs of preterm infants that would eventually develop BPD (54), suggesting that a gut-lung axis may influence the development of BPD. While the mechanisms underlying the gut-lung axis require further exploration in BPD, work in other childhood diseases has suggested some potential mechanisms. Mouse models of childhood asthma suggest intestinal fungi are more immunostimulatory than bacteria, while intestinal bacteria are more responsible for producing metabolites detectable in the host’s serum (11). Gut intestinal fungi, mainly the yeast *Candida albicans*, are known to provoke strong immune responses in the host (55). Indeed, monocolonization with *Candida* can stimulate postnatal immune development (56). The greater abundance of *Candida*-dominated communities we observed in PPRD infants is intriguing in this regard.

Limitations of this study include that we transplanted whole fecal samples in the mouse FMT experiment. While we analyzed the microbiota transferred to the neonatal mice, it remains unclear which components of this complex mix of microbes, metabolites, and nutrients modulate lung injury. Future experiments may benefit from a multiomic approach to identify and validate potential candidate microbes or metabolites. Similarly, it is currently not feasible to confine the transfer of microbes to only one organ system. Quantifying how spillover from FMT might affect other host organs, such as the lungs, remains challenging. A strength of this study was the rigorous exploration of the multikingdom microbiota using the most up-to-date bioinformatic techniques (47) and the demonstration that the microbiome effect was transferable by FMT.

### Conclusions

In this prospective, observational cohort study of VLBW preterm newborns, baseline compositional differences in the fungal intestinal microbiome predicted the eventual development of BPD. Furthermore, we showed that this dysbiotic microbiota could augment lung injury in a mouse model of BPD. These findings suggest that the gut microbiota can modulate lung injury and represents a therapeutic target in newborn lung disease.

## Author Contributions

Conceptualization: KW, NA, BP, SC, JP, AT. Clinical Sample and Data Collection: KW, MS, IM, EM, JD. Experimentation: KW, MS, AA, BH, KT, CW. Analysis: KW, IM, IK, OA, LT. Manuscript: KW. All authors have approved the final manuscript as submitted.

## Data Availability

Raw sequencing files are deposited at the National Center for Biotechnology Information Sequence Read Archive (BioProject: PRJNA982880). Processed data files are available at github.com/WillisLungLab/MiBPD

## Inclusion and Diversity

We support the inclusive, diverse, and equitable conduct of research. Multiple authors of this paper self-identify as underrepresented ethnic or gender minorities, or as a member of the LGBTQA+ community. While citing references scientifically relevant to this work, we also actively worked to promote gender balance in our reference list.

## Data Availability

All data produced in the present study are available upon reasonable request to the authors. Raw MiSeq and RNAseq data are deposited at the NCBI sequence read archive (BioProject: PRJNA982880). Processed data files and custom code are available at github.com/WillisLungLab/MiBPD

https://www.ncbi.nlm.nih.gov/bioproject/?term=PRJNA389800

## Acknowledgments

We thank the infants, their parents, and the study coordinators, without whom this study would have been impossible. Confocal microscopy was performed at the UAB High-Resolution Imaging Facility with the assistance of Robert Grabski.

## ONLINE DATA SUPPLEMENT

### Key Resource Table

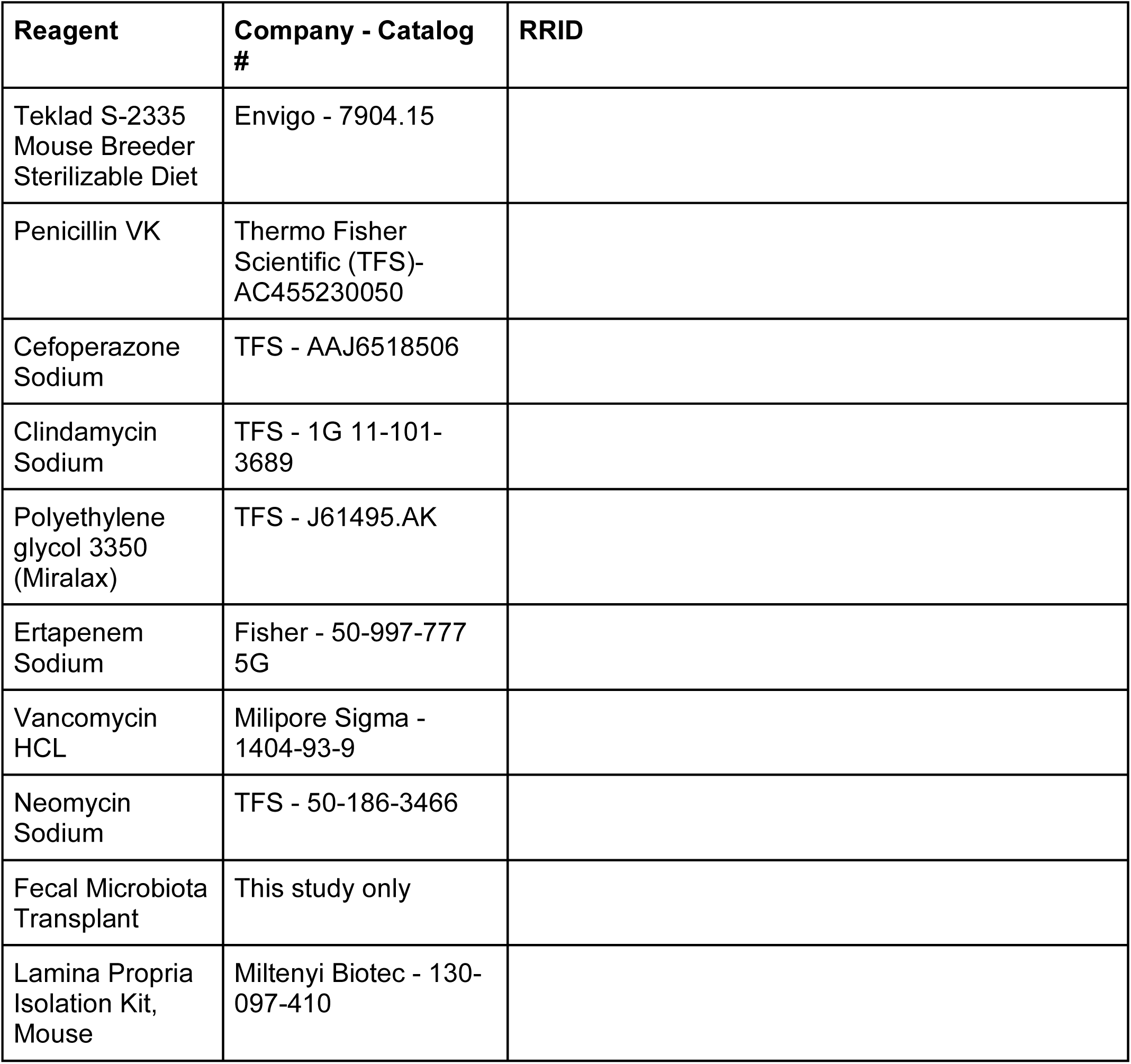
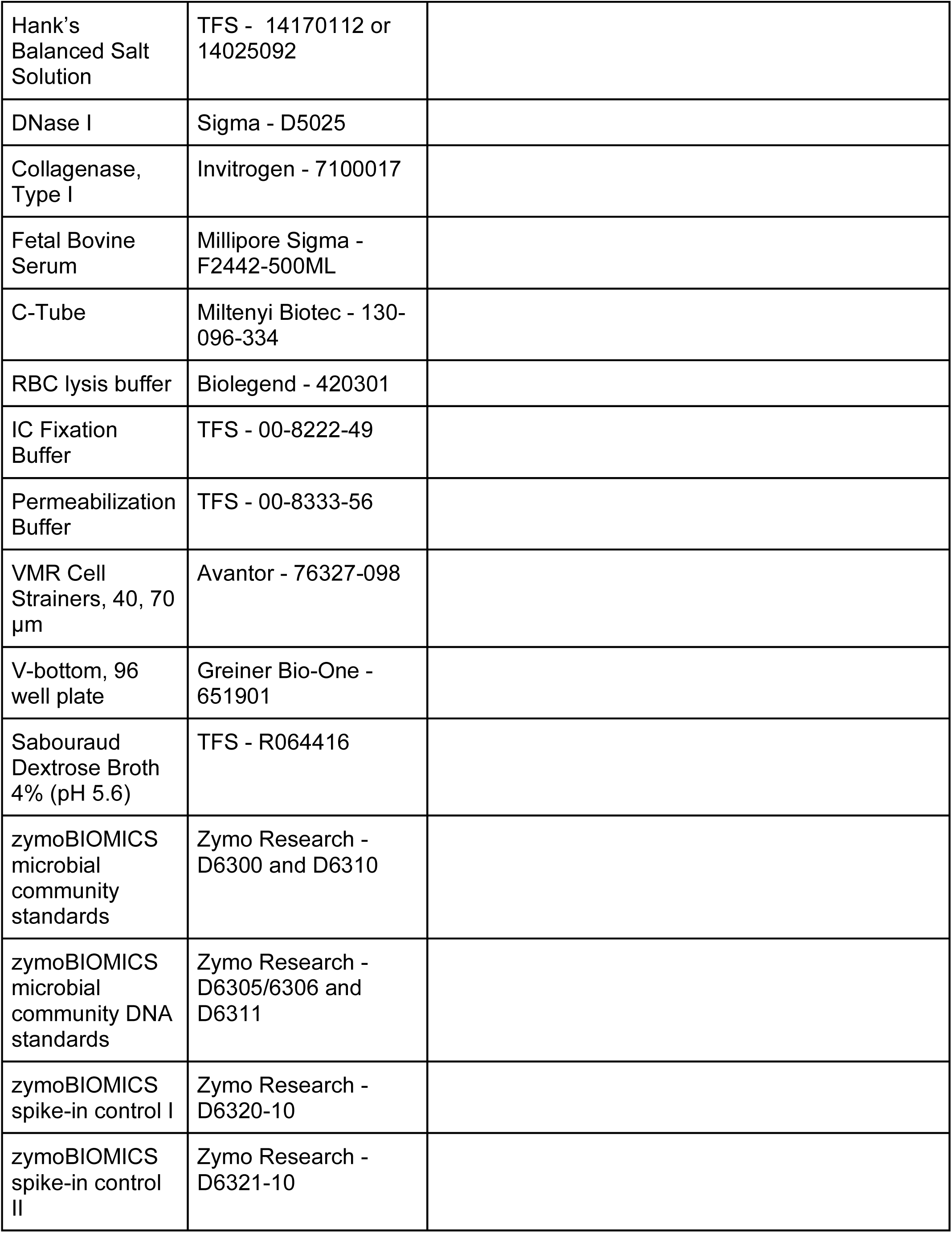
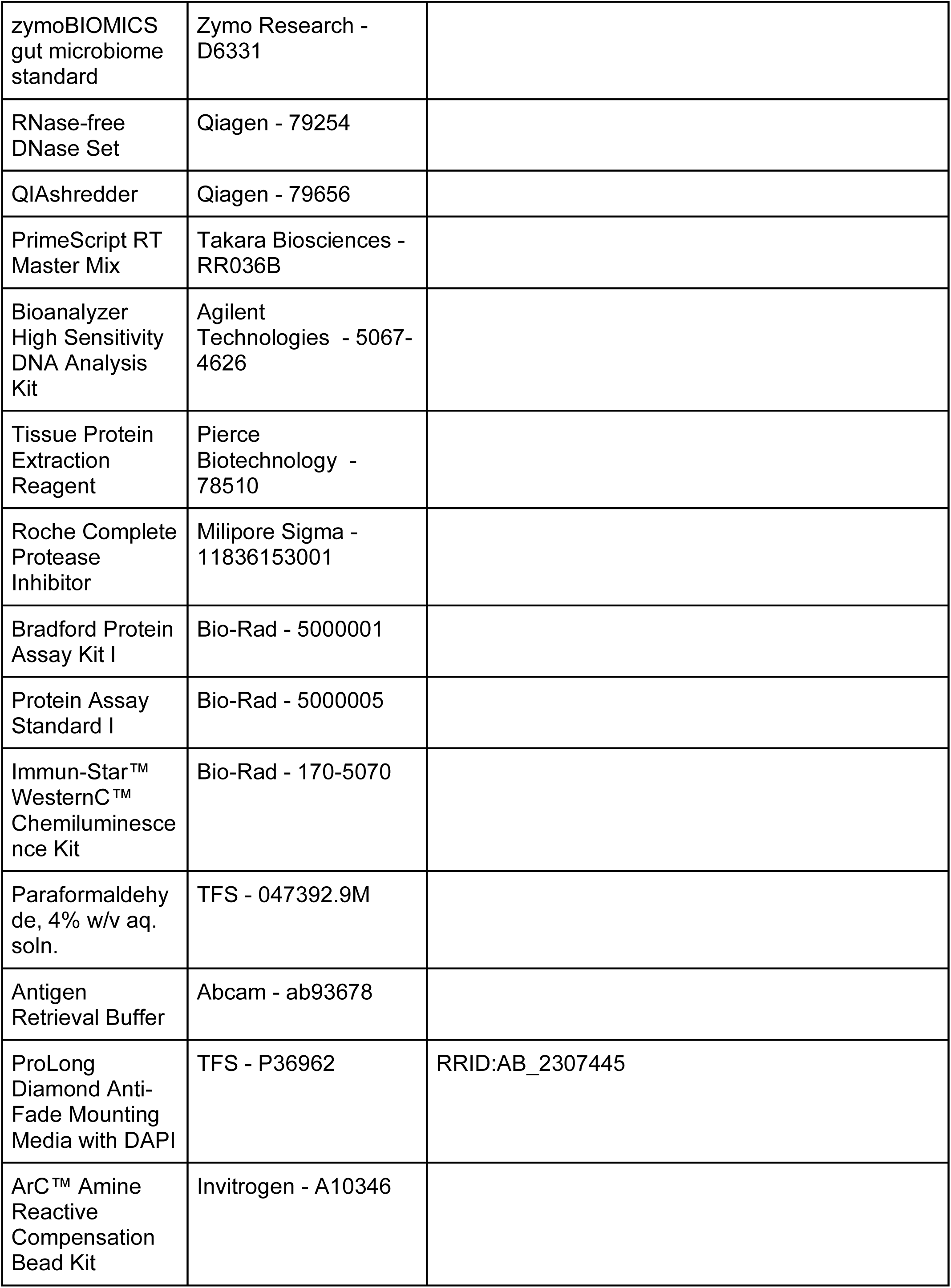
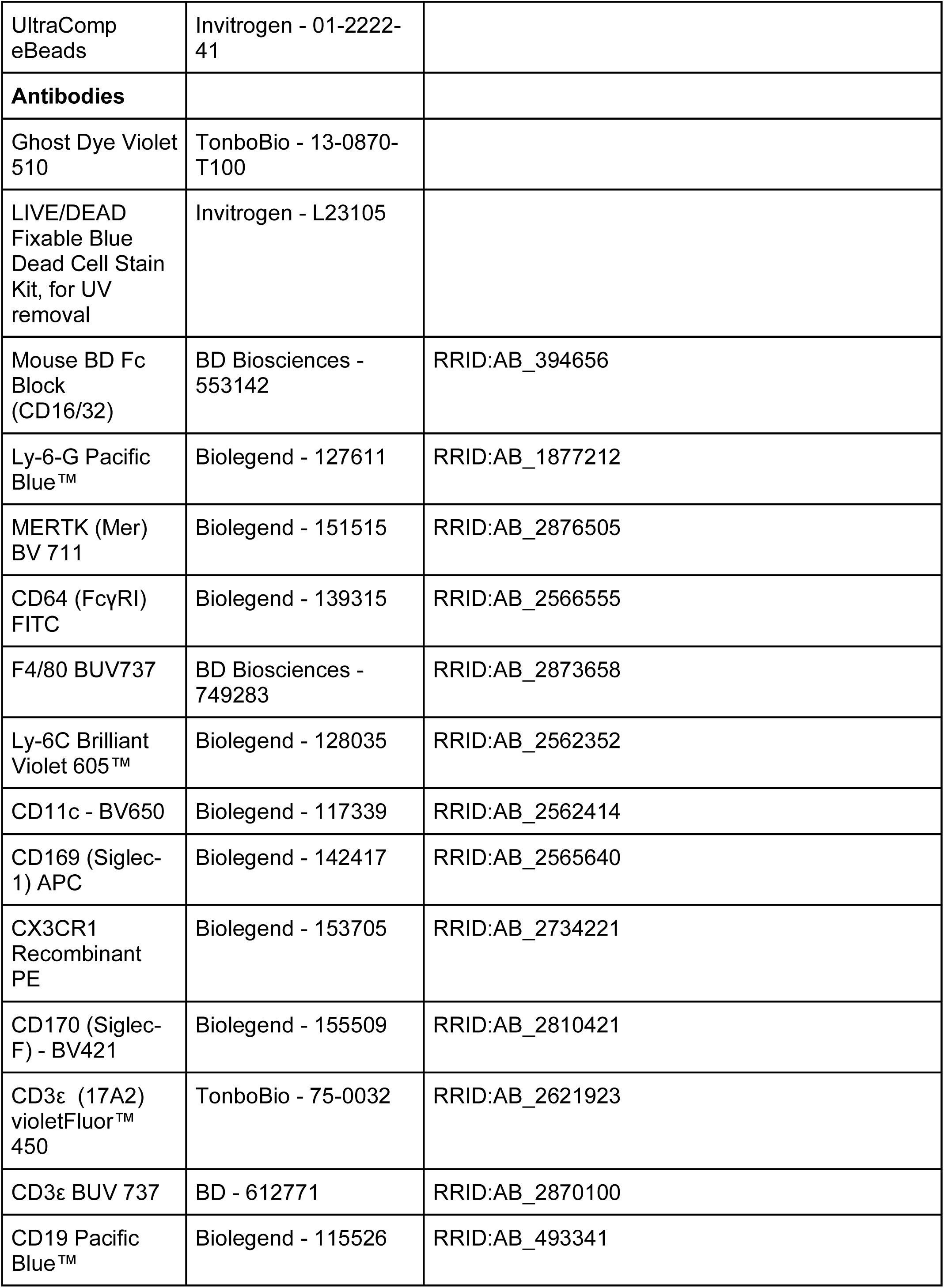
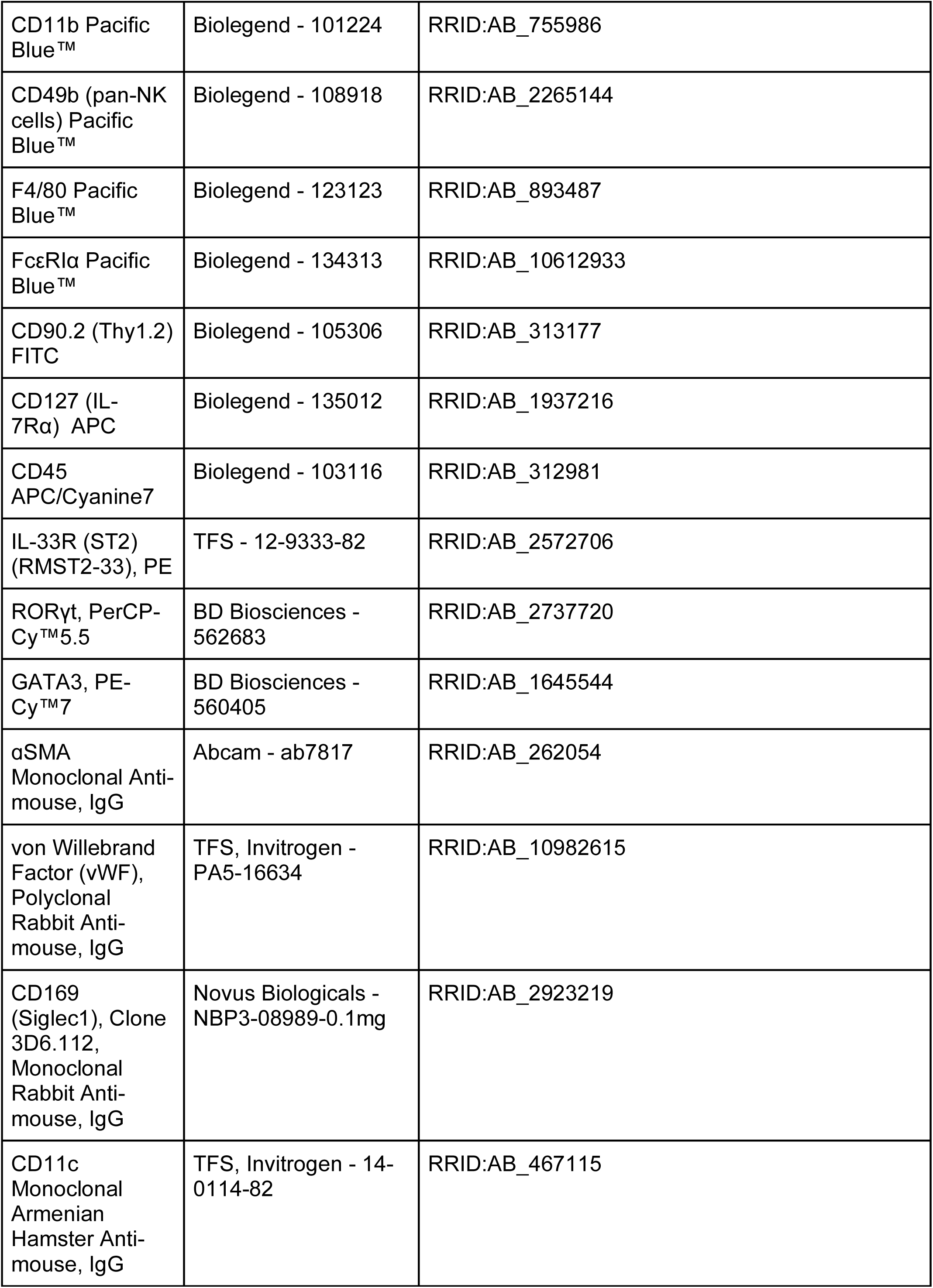
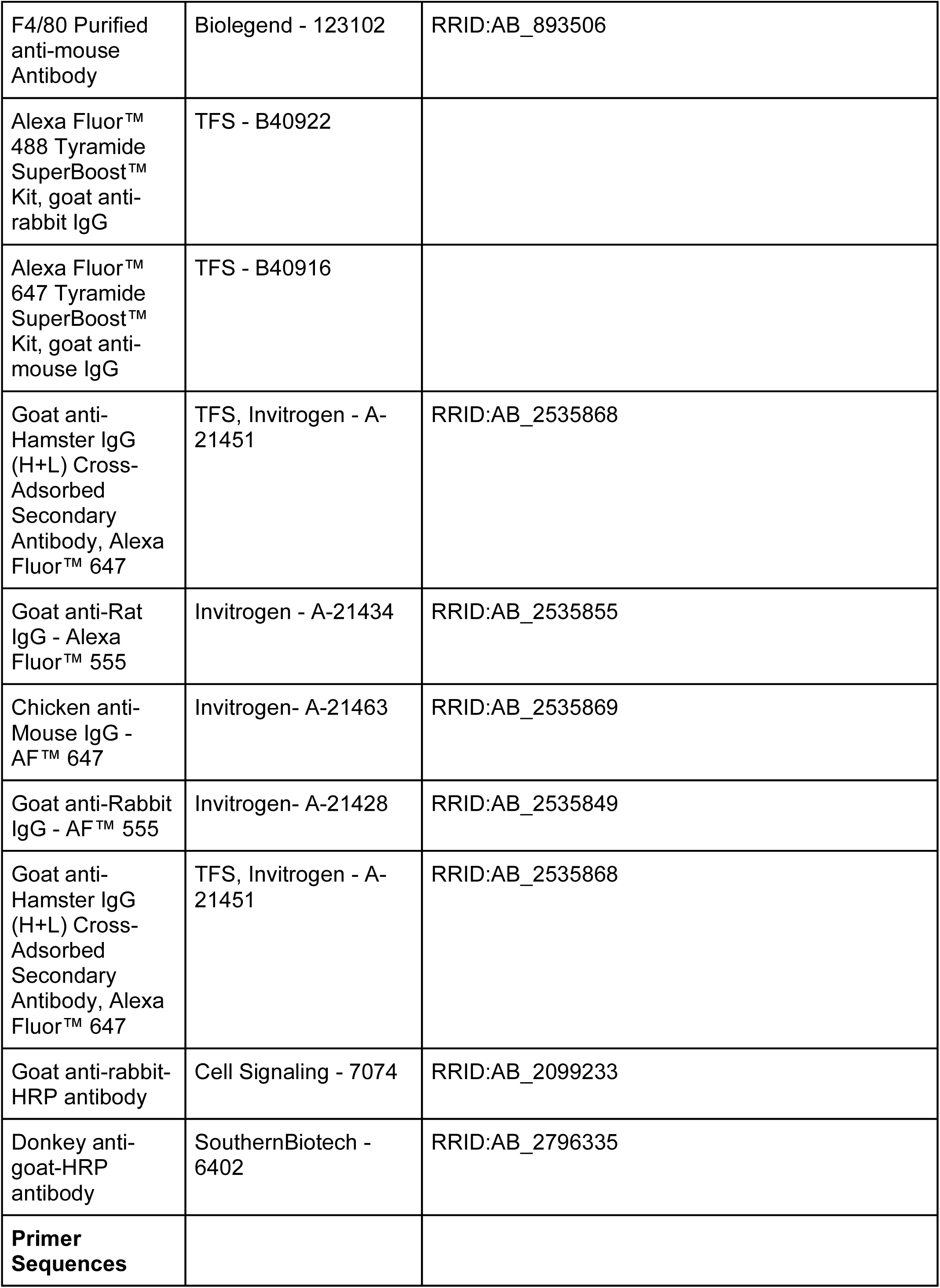
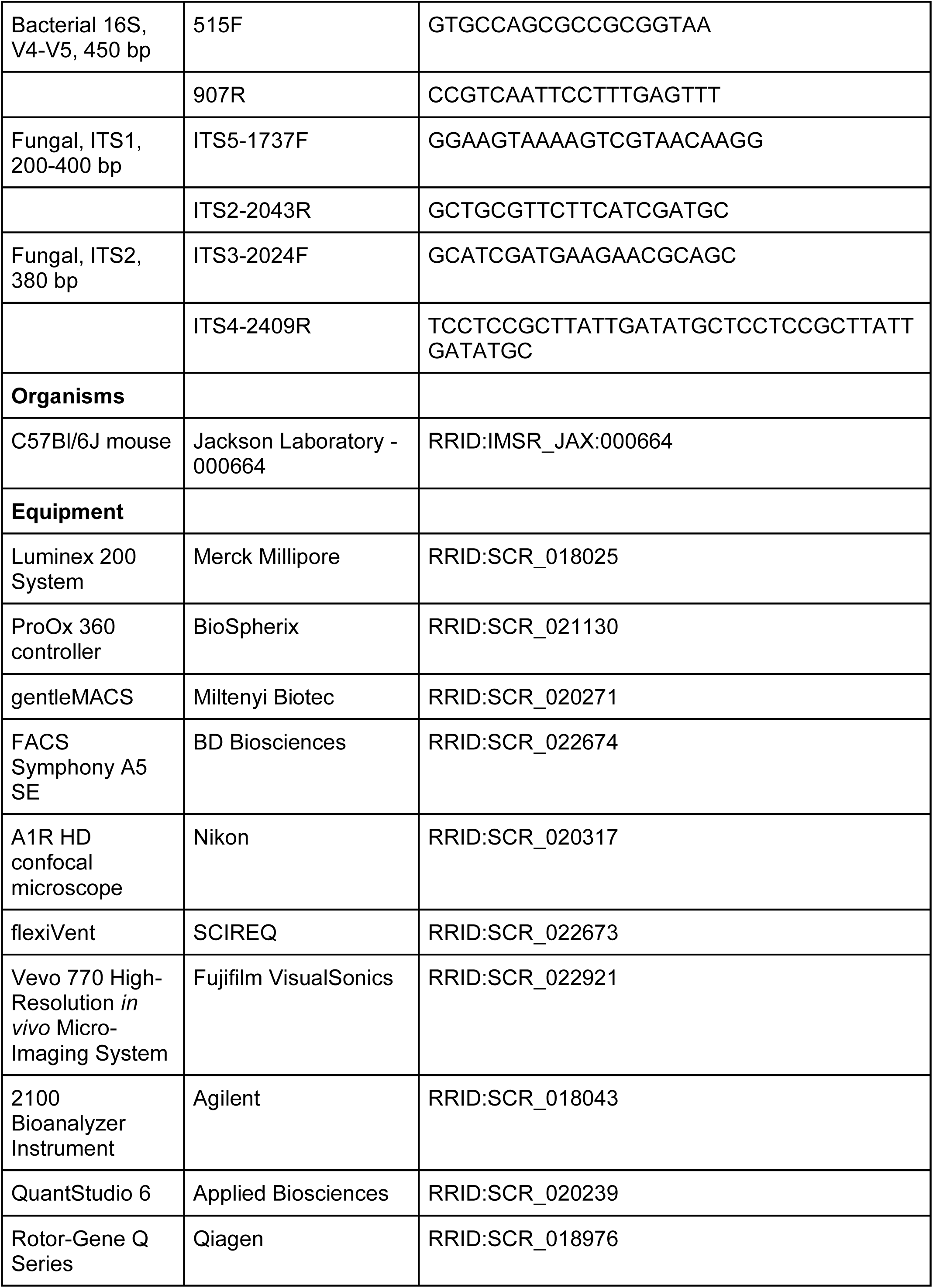
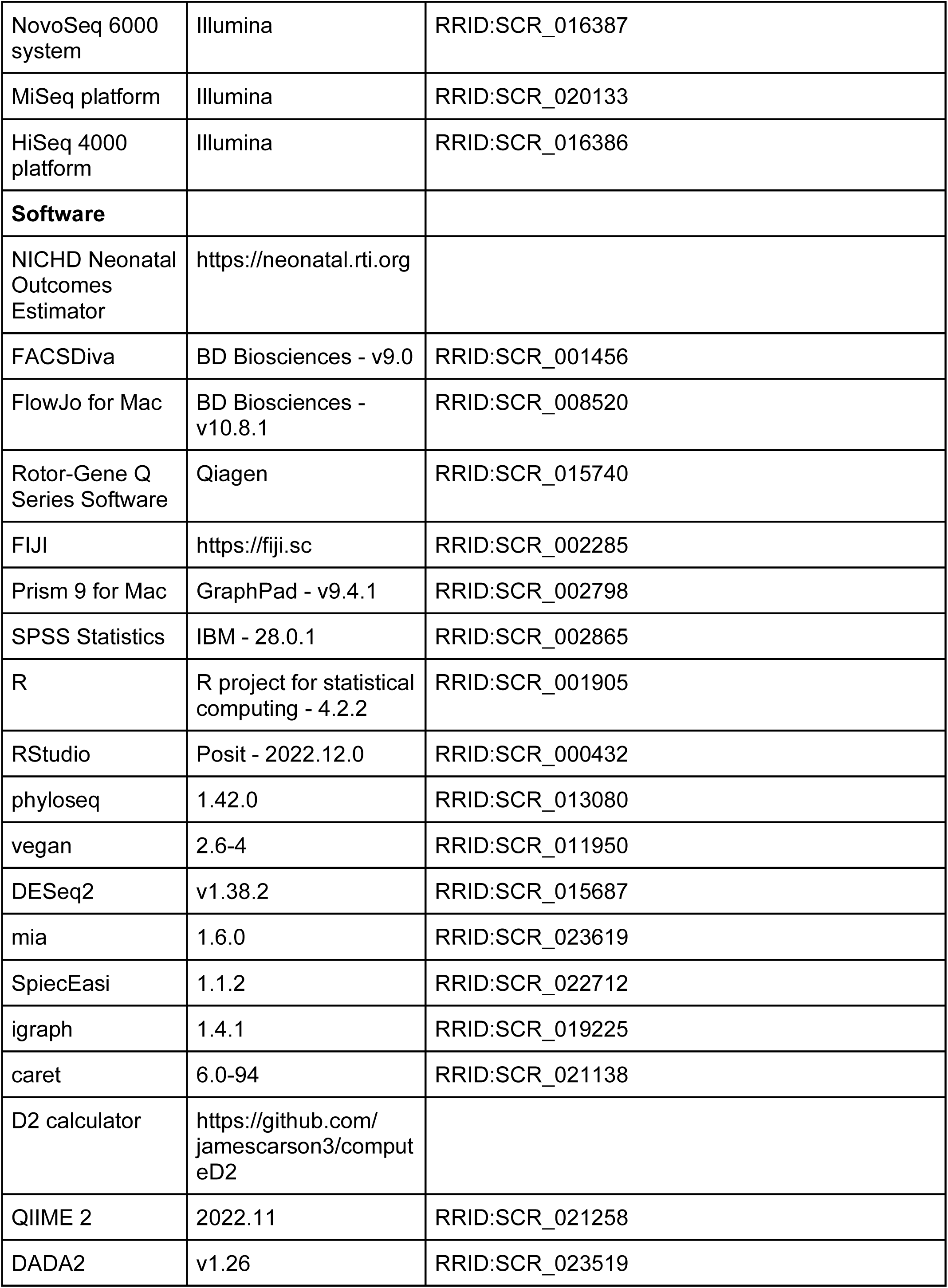
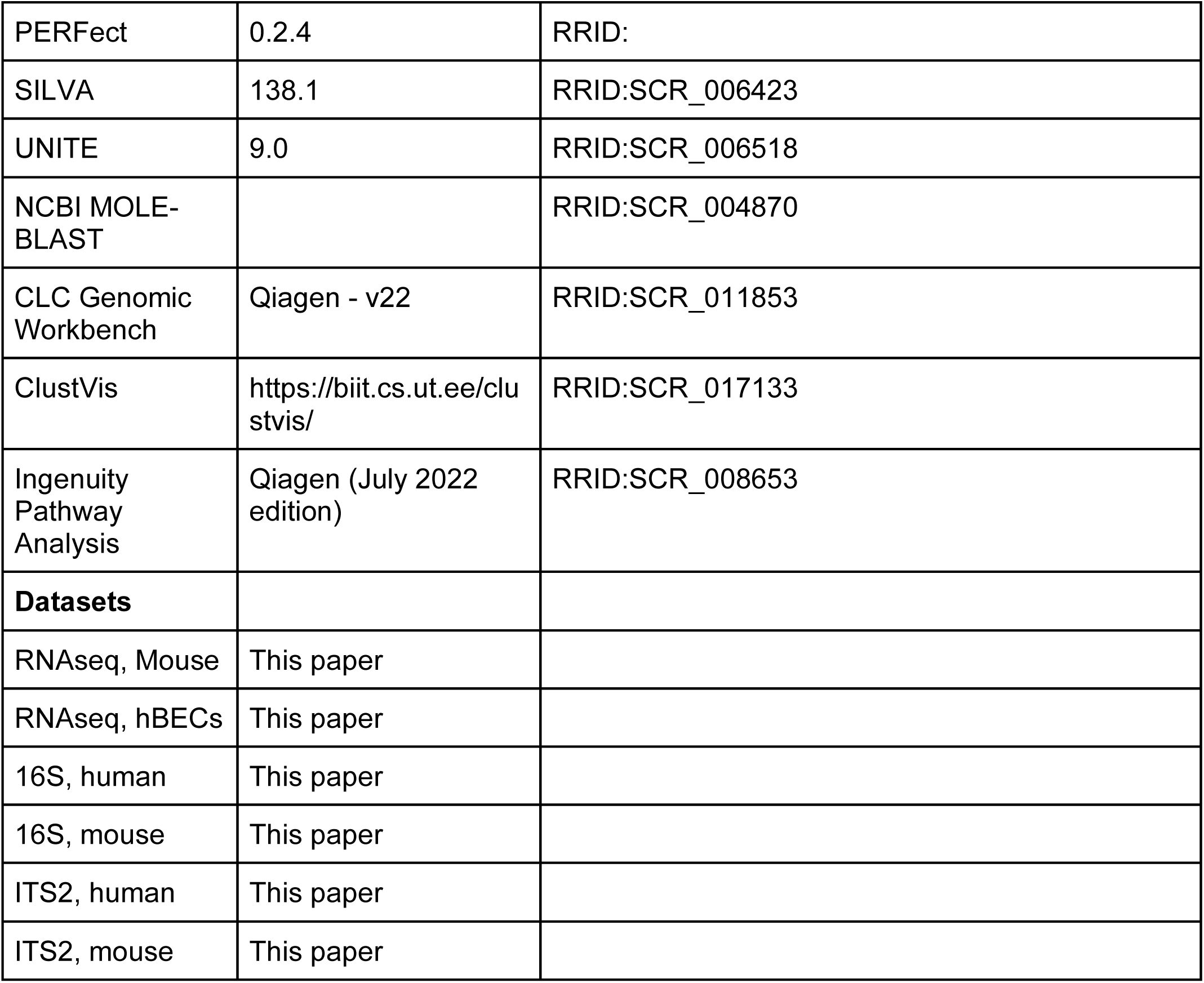

### Resource Availability

Lead Contact: Further information and requests for resources and reagents should be directed to and will be fulfilled by the lead contact, Kent Willis.

Materials Availability: This study did not generate any unique reagents or transgenic animals.

Data Availability:

- Raw RNA-seq, 16S and ITS2 sequencing data are deposited at the NCBI Sequence Read Archive (Access Number: PRJNA982880).
- Processed data files and original code are available at github.com/WillisLungLab/MiBPD.
- Any additional data required to reanalyze the data presented in this paper are available from the lead contact upon request.

### Experimental Model and Subject Details

Study Subjects:

University of Tennessee Health Science Center cohort: This study was approved by the Institutional Review Board of The University of Tennessee under protocol 17-05311-XP and conducted in accordance with the Declaration of Helsinki. We performed a prospective observational cohort study at the regional NICU at Regional One Health (Memphis, TN, USA) from July 2017-January 2020, pre-registered with clinicaltrials.gov (NCT03229967). The Microbial Influence on the Development of Bronchopulmonary Dysplasia (MiBPD) study approached the mothers of all inborn very low birthweight infants (VLBW, < 1500g) without known major congenital anomalies or immunodeficiency for potential consent for enrollment during the first week of life unless they were enrolled in a concurrent trial of probiotics that enrolled < 10 infants during this period. Mothers provided written consent on behalf of themselves and assent for the inclusion of their newborn prior to enrollment. Samples were collected weekly for the first month of life. Meconium samples collected within the first 72 hours have been described[1]. Analysis in this study was constrained to fecal samples collected during the second week of life. Demographic and clinical data were collected to describe the clinical course and to identify the severity of bronchopulmonary dysplasia (BPD) according to the NICHD workshop definition [2]. The risk of developing BPD was predicted using the NICHD Neonatal BPD Outcomes Estimator (neonatal.rti.org, 2022 updated formula)[3]. Human subjects data is reported in accordance with the extended STROBE guidelines [4].

University of Alabama at Birmingham cohort: This study was approved under protocol IRB-300003994 by the IRB at the University of Alabama at Birmingham (UAB, Birmingham, AL, USA). Samples from this cohort were collected in oxygen-reduced PBS immediately prior to FMT to the mice. The samples were used for the generation of the pseudohumanized mouse model and were collected from August 2021 to June 2022.

#### Animals

Animal experiments were performed at UAB under IACUC protocol 22042. 8-week-old female C57Bl/6J mice (Stock Number 000664, RRID:IMSR_JAX:000664) were purchased from the Jackson Laboratory and allowed to acclimate in specific-pathogen free housing with 4 animals per cage, on a 12-hour light/dark cycle, at 20-24C, with *ad libitum* access to irradiated rodent chow (Teklad 7904, Envigo) in the vivarium at the UAB for two weeks. Animal studies were performed in accordance with the recommendations in the Guide for the Care and Use of Laboratory Animals of the National Institutes of Health under protocol 22042, approved by the Institutional Animal Care and Use Committee at UAB. Animal experimentation is reported in accordance with the ARRIVE guidelines [5].

#### Generation of pseudo-humanized mice

Pseudo-humanized mice were generated by fecal microbiota transplant (FMT) from human infant donors with or without severe BPD, following antibiotic preconditioning, as previously described [6–8], generating mice we named NoBPD-FMT and BPD-FMT, respectively. Briefly, 9-week-old dams were subjected to antibiotic administration for two weeks in their drinking water, which they were allowed to access *ad libitum*. The first week consisted of systemically absorbable antibiotics: penicillin VK, cefoperazone sodium, and clindamycin sodium (all at a concentration of 1mg/mL). This was followed by a bowel cleansing regimen by oral-gastric gavage with 200 uL of polyethylene glycol 3350 at 425 g/L. This was repeated twice. Mice were placed on the second week of antibiotic therapy in the drinking water, which consisted of the non-absorbable antibiotics ertapenem sodium, vancomycin sodium, and neomycin sodium (all at a concentration of 1mg/mL). Cage bottoms were changed daily and after each gavage of polyethylene glycol, as mice are coprophagous and could reinoculate themselves with stool. After two weeks of antibiotic treatment, female mice were given stool from neonatal human donors with or without BPD. The stool was preserved in oxygen-reduced sterile PBS. We then performed FMT. Mice in each group were gavaged twice with 200 uL of sample from the same human donor. Stool samples were collected after antibiotic therapy, after FMT engraftment, or at the time of harvest. After engraftment, dams inoculated with the same donors FMT were bred simultaneously to generate neonatal mice for further experimentation. On day 2 after birth, the pups were inoculated with the same donor FMT to create BPD-FMT and NoBPD-FMT mice, respectively. Each experiment was conducted with at least two litters arising from the same human donor and a minimum of three human donors in each experimental group (6 litters=3 donors/group).

#### Neonatal hyperoxia exposure model

We utilized our established, standardized hyperoxia-exposure model of BPD [9, 10]. We time-mated the mice to produce multiple simultaneous birth cohorts with the same perinatal exposure. On the day of birth, pups from two consecutively born litters with the same perinatal exposure were pooled, and the pups were re-distributed evenly between the two dams. Litter sizes for all experiments were adjusted to 5–8 pups per treatment group to minimize nutritional effects on lung development. One pooled litter was randomly assigned to hyperoxia [fraction of inspired oxygen (FiO_2_) 0.85]) and the other to air (FiO_2_ 0.21). Pups were exposed continuously for 11 days, starting on the third day after birth (P3-14). Oxygen concentrations were maintained using a ProOx monitor (Biospherix). Dams were rotated daily between the two pooled litters to limit the complications of hyperoxia. Growth was monitored daily for all pups throughout the experiment.

## Method Details

### Data collection

Clinical data were collected from the medical record that described clinical illness severity, infant characteristics, maternal and infant demographics, the extent of prematurity, respiratory parameters, and known microbiome modulators when the infant reached 36 weeks’ corrected gestational age or was discharged. BPD was adjudicated at 36 weeks corrected gestational age using the NICHD workshop definition [2]. We quantified clinical illness severity with the Critical Risk Index for Babies II (CRIB II)[11]. The risk of developing BPD was predicted using the NICHD Neonatal BPD Outcomes Estimator (neonatal.rti.org, 2022 updated formula)[3].

### Patient and public involvement

Neither patients nor the public were involved in designing this research study.

### RNA-seq

Bulk RNA-seq was performed on isolated tissue from the left middle lobe using established methods [12]. Briefly, 25 mg of tissue was collected in RLT buffer (Qiagen) with 1% (v/v) 2-Mercaptoethanol (BME, Sigma Aldrich), homogenized using a QIAshredder (Qiagen), and isolated with an RNeasy Kit (Qiagen) with on-column removal of DNase. DNA purity and concentration were validated with the Bioanalyzer High Sensitivity DNA Analysis chip (Agilent Technologies) at the Heflin Genomics Core Facility. Samples were sent to Novogene for RNA library preparation and transcriptome sequencing on the HiSeq 2500 platform (Illumina) using a 150-nucleotide paired-end read length [12].

### FlexiVent pulmonary function testing

The P14 mice used for this analysis were not used for other analyses. Forced oscillometry was performed as described [13, 14]. Briefly, we performed a tracheostomy on the sedated mice with the appropriate cannula and secured it with two 3-0 sutures. We performed the neonatal pulmonary function assessment program on the flexiVent system (SCIREQ). We calibrated the system with the tracheal cannula prior to each mouse.

### Histology and immunohistochemistry

We gravity inflated the right lung of each mouse to 20 cm H2O with 4% formalin (Thermo Fisher Scientific), and were placed into 100% ethanol within 16 h. After paraffin embedding, we cut 4-5 m slides at 3 random levels through the block that were stained with hematoxylin and eosin for morphometry. At each level, three slides were made from adjacent sections for immunohistochemistry. These slides were deparaffinized and rehydrated with graduated concentrations of ethanol. We performed antigen retrieval with heated 10 µM Sodium Citrate Buffer with 20% Tween. Slides were fenestrated, blocked in 1% BSA and anti-CD16/32 (BD Biosciences) for 30 min, then incubated overnight with either anti-vWF and or anti-αSMA at 4C in incubation buffer (1% BSA, 1% normal donkey serum, 0.3% Triton X-100, and 0.01% sodium azide in PBS). The next day, they were washed in PBS, blocked again, counterstained with the appropriate secondary antibodies, washed, and finally counterstained with ProLong Diamond Anti-Fade Mounting Media with DAPI (Thermofisher Scientific). Images were captured using an A1R HD inverted confocal microscope (Nikon).

### Bacterial and fungal microbiota analysis

The first non-transitional stool sample collected during the second week of life was used to assess the human gut microbiota. In mice, liquid nitrogen snap-frozen homogenized whole left lung or a 1 cm section of terminal ileum were used because of the advantages in quantifying the adherent mucosal microbiota [15]. We used ZymoBIOMICS whole organism and DNA microbial community standards (Zymo Research) as well as sterile-filtered PBS to create pairs of positive and negative controls appropriate for each step of the sample collection and isolation procedure. Microbial DNA was extracted using the ZymoBIOMICS DNA Miniprep Kit (Zymo Research) with the addition of a lyticase (200 Units per 1 mL sample volume) predigestion with 10-minute bead bashing on a Fisher Vortex Genie 2 (Thermo Fisher Scientific) at RT, then 37 C with gentle agitation for 20 min, as previously validated to facilitate extraction of fungal DNA [16]. Extracted DNA was sequenced using the MiSeq platform (Illumina) at Novogene (human samples) or Argonne National Laboratory (mouse samples) using 16S V3-4 (bacterial) rRNA with primers 515F-806R and the internal transcribed spacer (ITS) 2 (fungal) rRNA gene primers. Primer design was chosen based on prior published optimization for mycobiome amplification in respiratory samples [17, 18]. Quantitative RT-PCR of the ITS1, ITS2, and 16S genes was performed as described [19], using TaqMan probes (Thermo Fisher Scientific) validated with ZymoBIOMICS controls (Zymo Research) and additional human and mouse samples. QC was performed at the UAB Genomics Core.

### Bioinformatics

Microbiomics - We used QIIME 2 [20] to import and trim ITS2 and 16S rRNA gene amplicon sequences. DADA2 [21] was used to denoise and cluster amplicon sequence variants (ASVs). ASV taxonomy was assigned with the RDP naive Bayesian Classifier using the SILVA 138.1 and UNITE 9.0 databases, for bacteria and fungi, respectively. All ASVs aligning to the division Chytridiomycota or unassigned at the phylum level were then removed because confirmatory analysis with BLAST revealed these sequences to be either host mitochondrial or plant material contaminates. Contamination source modeling was then performed using the R package *PERFect* [22], using permutation filtering with p-value ordering, at an alpha value of 0.05.

Decontaminated ASV tables were then exported into R where they were normalized, transformed, and analyzed with the following packages: *vegan*, *phyloseq*, *DESeq2*, *SpiecEasi* (Sparse Inverse Covariance Estimation for Ecological Association Inference), *tidyverse, igraph, mia,* and *caret*. Alpha diversity was quantified with the Shannon and Simpson Indices. We displayed beta diversity with both principal components analysis of Euclidean distances and principal coordinates analysis of Bray-Curtis dissimilarity matrices. Significance testing was performed using PERMANOVA, and evaluated confounding by differential dispersion using PERMDISP. Feature selection was performed using DESeq2 and Random Forest using *ranger* and *caret*. Unsupervised clustering was performed using Dirichlet-Multinomial Mixture modeling using the packages *mia* and *DirichletMultinomial*. Feature selection was performed using the *fitted* function within *DirichletMultinomial*. Mapping of clinical data to DMM clusters was performed using Canonical Correspondence Analysis (CCA) using the *vegan* package.

RNAseq - Raw sequencing data was aligned, processed and normalized, then differential gene expression was calculated using the CLC Genomic Workbench (Qiagen). Genes with adjusted p-value (q) < 0.05 and log(FoldChange) > 0.301 were considered differentially expressed. Heatmaps and principal components analyses were graphed using ClustVis. Differentially expressed genes were input into Ingenuity IPA (Qiagen) to identify significantly regulated networks and pathways.

### Machine learning models

Gradient boosting algorithms utilize a group of shallow trees and assemble them into a single prediction score to improve their predictive properties [23]. We developed a Light Gradient Boosted Machine (LightGBM) [24] model to predict BPD using both clinical demographics at birth and the corresponding microbiome data. Clinical parameters were curated from those previously validated in the Neonatal BPD Outcome Predictor [3], and included gestational age at birth, maternal-self-reported race, birth weight, temperature on admission to the NICU, and mode of ventilation. Since the sample size of the dataset is modest in comparison to the number of unique variables in the microbiota data, we only utilized the kingdom and phylum-level microbiota data to maintain generalizability and reduce the risk of over-fitting. To construct the model, we split the dataset into two sets: 80% into a model-building discovery set and 20% as a holdout validation set. We used 5-fold cross-validation in the discovery set to tune the hyperparameters of the algorithm. We then assessed the accuracy of the training phase using the validation set. A Bayesian algorithm [25] was chosen for hyperparameter tuning. To tune the model, we adjusted the number of trees, feature fraction, learning rate, and the maximum depth of the trees. In addition, the training phase was stopped if there was no consecutive improvement in the validation set. Finally, the LightGBM model from the cross-validation was performed on the holdout set, and we calculated the resulting AUC of the ensemble model.

We also implemented the default variable importance analysis of the LightGBM algorithm to evaluate the contribution of the variables into models, in which their contribution was weighted over folds according to the model’s performance on the corresponding validation set. We also evaluated the directional effect of the variables on the BPD risk prediction score within a 95% confidence interval (inverse or direct relationship) by systematically increasing by one unit for categorical variables and one standard deviation for continuous variables.

Statistical analysis: Descriptive statistical analysis of the human data was performed in SPSS Statistics (IBM). Murine data was analyzed primarily in R and Prism 9 (GraphPad, Inc.).

## Supplemental Information

### Extended Data

#### The fungal microbiota predicts BPD development

To test the predictive potential of the mycobiota, we first developed random forest machine-learning models using fungal or bacterial microbiome as exploratory analyses (supplemental figure S4). The fungal model (supplemental figure S4a-b), which had an area under the receiver operating curve of 0.71, identified several rare fungal genera, such as *Penicillium*, *Gamszarea*, and *Verticillium* as the most discriminatory taxa. In the bacterial model, which had a receiver operating curve of only 0.58, *Enterococcus* and *Acinitobacter* were the most discriminatory taxa (supplemental figure S3c-d).

#### Distinct fungal community clusters are linked to clinical outcomes

Having shown that fungal microbiota features are associated with BPD development, we sought to identify other clinical characteristics that may contribute to heterogeneity in the composition of the neonatal mycobiome. First, we used PERMANOVA to screen potential additional contributors to fungal community composition. Of the potential fungal microbiome covariates we examined none remained significant after adjustment for false discovery (supplemental table S3, figure S6). When we tested potential bacterial covariates we made similar observations (supplemental table S4).

#### Heterogeneity in the mycobiome

We next used unsupervised clustering to explore heterogeneity in fungal community features using Dirichlet-Multinomial Mixtures (DMM) modeling [26] (supplemental figure S8). In contrast to our previous approach, this method identifies clusters based on their microbial composition, which can then be associated with clinical characteristics. This permits the identification of microbial heterogeneity within disease states that can go otherwise undetected. In this study, Laplace analysis for the whole mycobiome identified 5 clusters as the optimal fit point (supplemental figure S9a). Membership in Cluster 1 of the fungal DMM model (supplemental figure S8a-c) was driven by the relative abundance of an *Aureobasidium* ASV and a *Saccharomyces* ASV (supplemental figure S9b). Membership in Cluster 2 was driven by a *Candida* ASV (supplemental figure S9b). Cluster 3 was driven primarily by an *Aureobasidium* ASV. Cluster 4 was driven by *Candida* and *Aureobasidium* ASVs and to a lesser extent by an *Issatchenkia* ASV. Finally, Cluster 5 was almost entirely driven by a *Candida* ASV. We then used canonical correspondence analysis (CCA) to map clinical characteristics that may be associated with the fungal composition profiles identified by the DMM components (supplemental figure S9c). Clusters 1 and 3 shared similar vectors and were closely associated with BPD and increased illness severity (CRIB-II). Cluster 2 was in opposition, and therefore associated with opposing markers of illness severity such as increasing gestational age (GA), birthweight (BW) and male sex-assigned at birth (AMAB). The limited number of samples in Cluster 5 were at right angles to the majority of these clinical variables and were associated with low fungal evenness (arbitrarily defined one ASV contributing greater than half of the relative abundance).

#### Dirichlet-Multinomial Models of NoBPD and BPD infants

Laplace analyses performed independently within the BPD or NoBPD cohorts identified 2 clusters as the best fit for each of these mycobiome subsets (supplemental figures S9d and g). In the NoBPD DMM model, membership in Cluster 1 was driven by *Saccharomyces*, *Aureobasidium*, and to a lesser extent a *Candida* ASV (supplemental figure S9h). Membership in Cluster 2 was driven by three different *Candida* ASVs and a *Saccharomyces* ASV. CCA suggests that membership in Cluster 1 associated with the the opposite clinical variables as NoBPD Cluster 1 (BW, GA and AMAB), with the exception of fungal abundance (supplemental figure S9i). In the BPD DMM model, membership in Cluster 1 was driven by *Saccharomyces* and *Candida* ASVs (supplemental figure S9e). Membership in Cluster 2 was primarily driven by an *Aureobasidium* ASV. On CCA, Cluster 1 was associated closely with BW and to a lesser extent with GA and death (supplemental figure S9f).

#### Bacterial DMM models

DMM modeling using the bacterial microbiome data is shown in supplemental figures S10 and S11. Of note, Laplace clustering for the whole microbiome indicated optimal clustering with 4 clusters (supplemental figure S11a), while clustering for the NoBPD and BPD infants was optimal with 2 clusters, each (supplemental figures S11d and g). We then performed variable importance analysis and identified bacterial ASVs with significant contributions to component formation (supplemental figure S11). Clustering in the whole microbiome model is driven by *Enterococcus*, *Enterobacter*, *Cellulosimicrobium*, *Escherichia-Shigella* and multiple *Klebsiella* ASVs. Clustering in NoBPD infants was driven by *Enterococcus, Staphylococcus* and multiple *Klebsiella* ASVs, while Enterococcus and Enterobacter and *Klebsiella* ASVs drove the BPD infant clusters. CCA showed Cluster 2 of the overal bacterial model was associated with BPD, critical illness, AMAB and positive CRP (supplemental figure S9c). Together, the DMM clustering between the fungal and multikingdom microbiome models is not indicative of any significant overlap.

**Figure S1.**
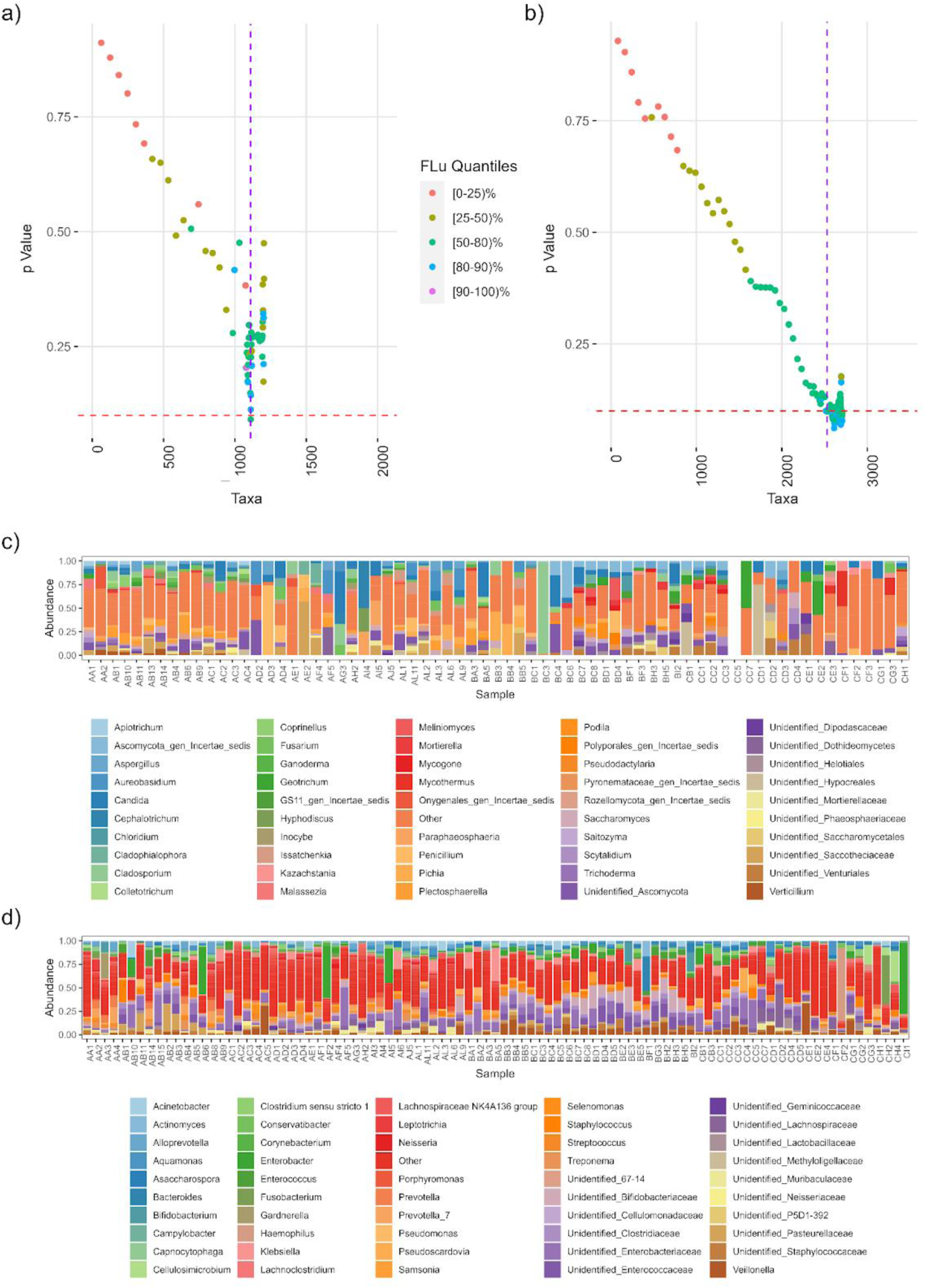
Source contamination modeling. a) Fungal taxa filtering loss plots. b) Bacterial taxa filtering loss plots. Please see tables of filtered taxa at github.com/WillisLungLab/MiBPD/SourceContamination. c) Average abundance of filtered fungal taxa. d) Average abundance of filtered bacterial taxa.

**Figure S2.**
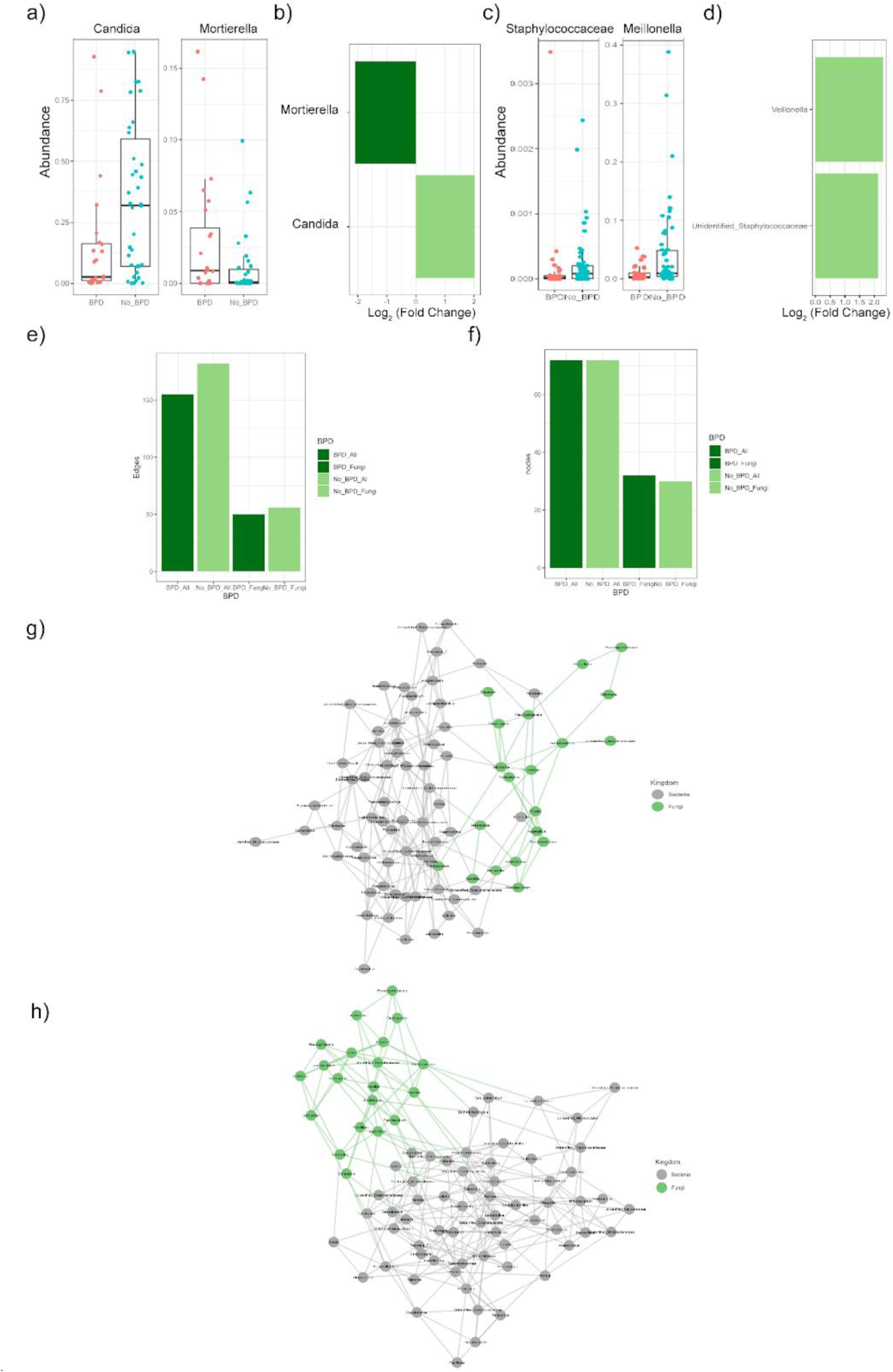
The BPD-associated mycobiome is unique. (a, b) Relative abundance of key fungi by DESeq2. (c,d) Relative abundance of key bacteria by DESeq2. (d) Number of edges in each SPIEC-EASI network. (e) Number of nodes in each SPIEC-EASI network. (e) Complete SPIEC-EASI BPD network with all genera labeled. (F) Complete NoBPD network with all genera labeled.

**Figure S3.**
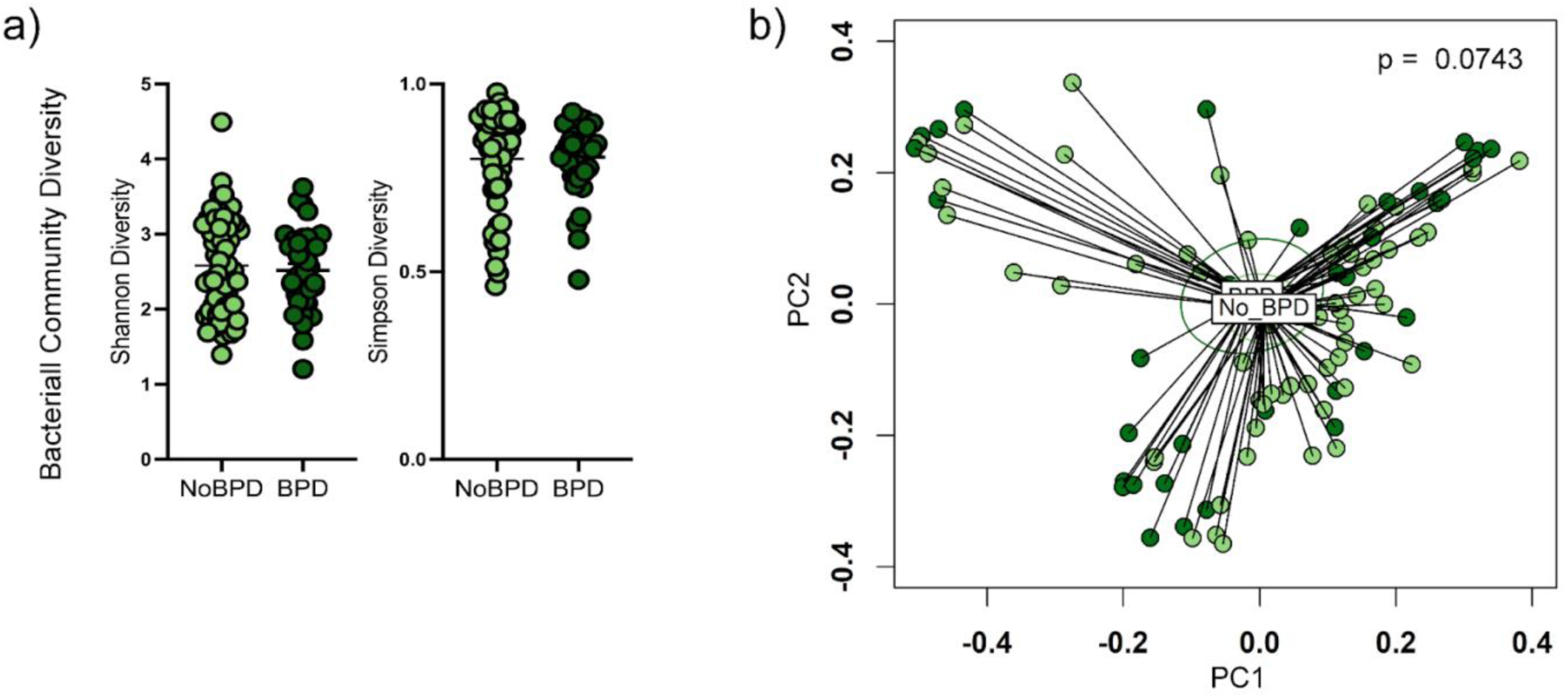
The overall composition of the multikingdom microbiome. (a) Alpha diversity. (c) Beta diversity (PCoA of Bray-Curtis dissimilarity, p = 0.0743, PERMANOVA, p = 0.3749, PERMDISP). Ellipses indicate the 95% confidence interval.

**Figure S4.**
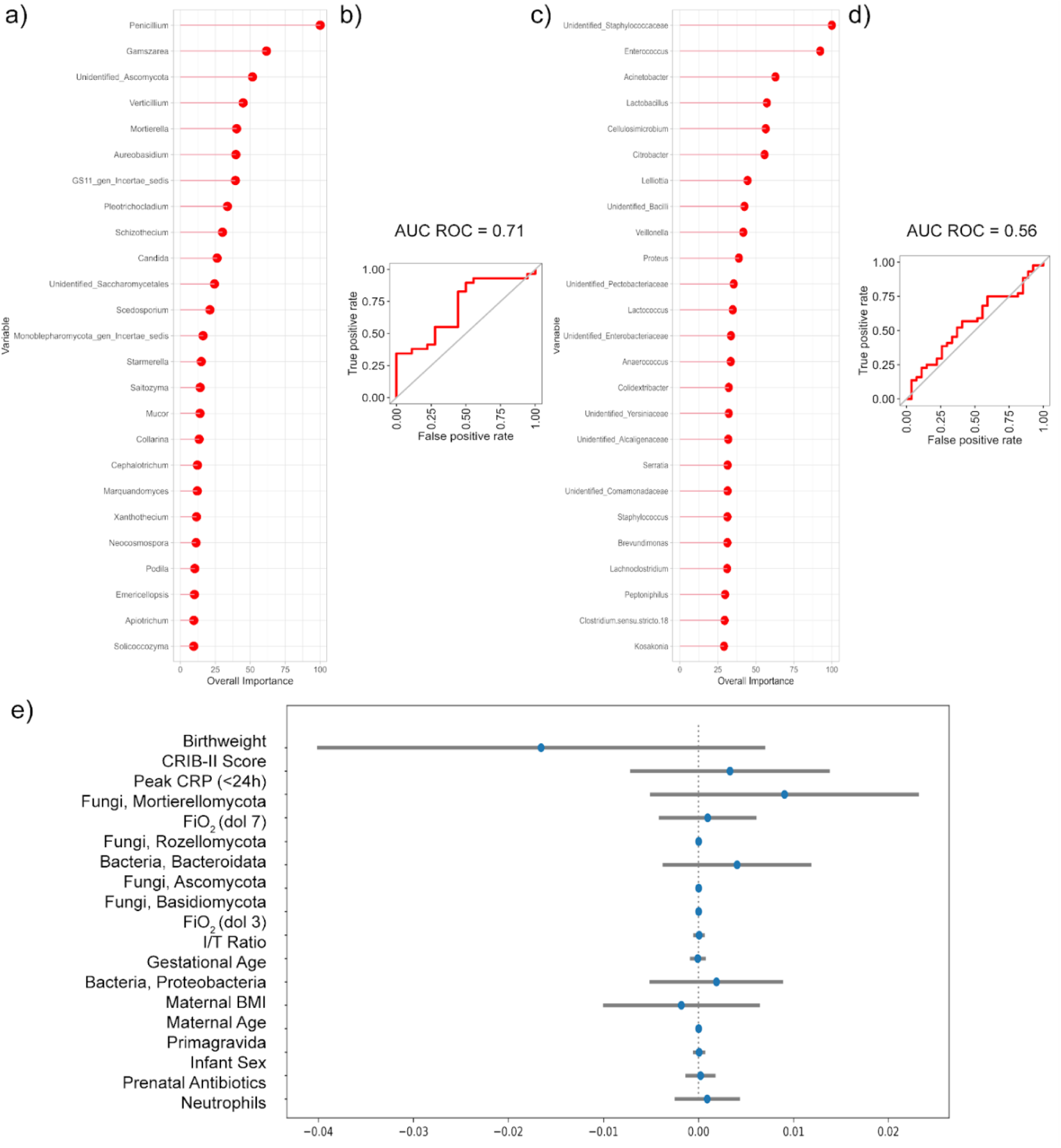
BPD status is predictable using machine-learning models of the fungal microbiota several weeks prior to the adjudication of the clinical diagnosis at 36 weeks corrected gestational age. (a-b) Exploratory random forest machine learning model using genera level mycobiome and clinical data (Should be considered exploratory due to high risk of overfitting). (c-d) Exploratory random forest machine learning model using genera-level bacterial microbiota and clinical data. (E) Directional variable importance analysis for the multikingdom light gradient boosted machine learning model.

**Figure S5.**
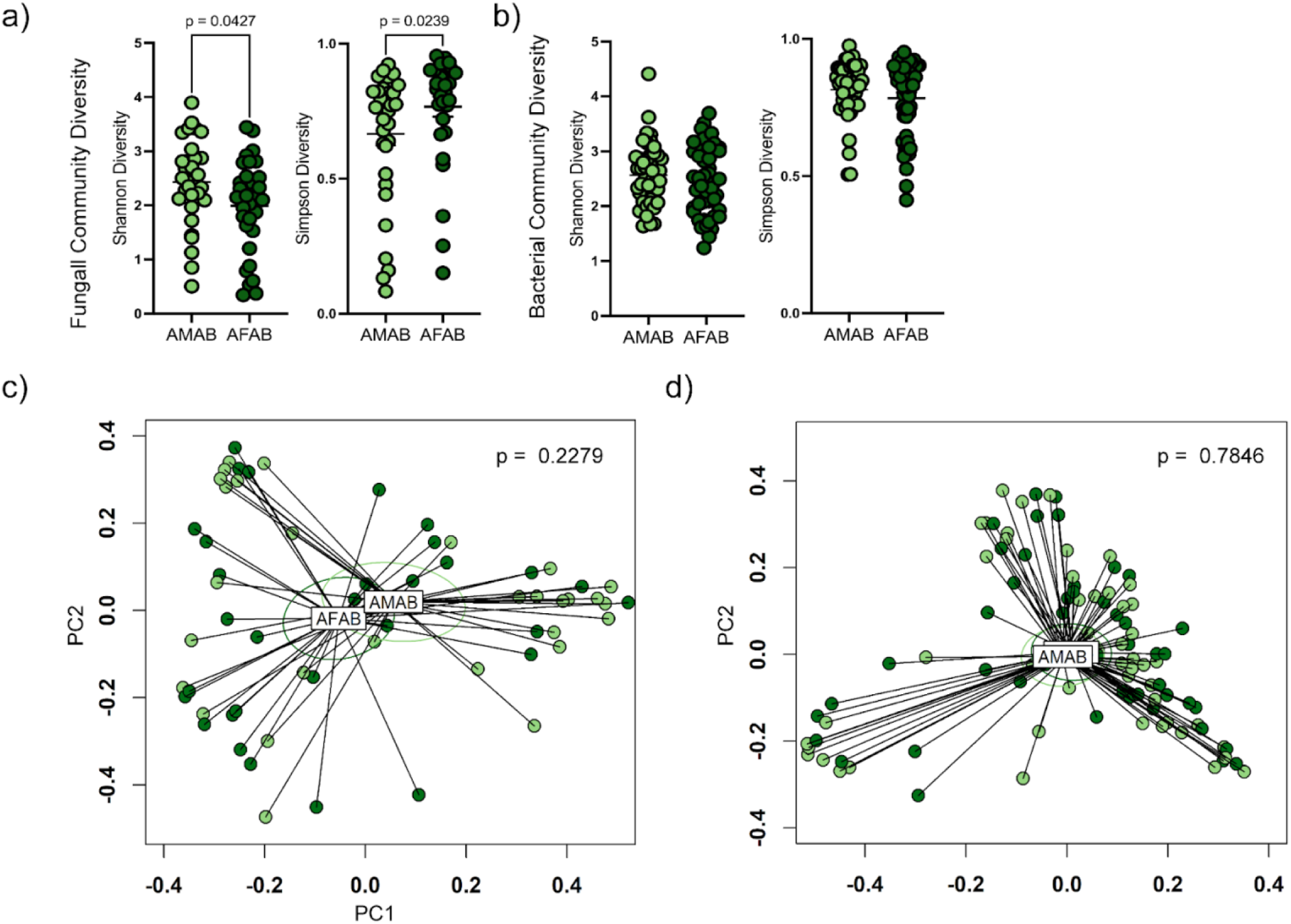
Mycobiome features differences by sex assigned at birth. (a) Fungal alpha diversity. (b) Bacterial alpha diversity. Shannon diversity was decreased in infants assigned female at birth (AFAB) as compared to infants assigned male at birth (AMAB, p = 0.0427, Mann-Whitney). Simpson diversity was increased in infants AFAB as compared to those AMAB (p = 0.0239, Mann-Whitney). (c) Principal coordinates analysis of Bray-Curtis dissimilarity of fungal beta diversity (p = 0.2279, PERMANOVA; p = 0.9464, PERMDISP). (d) Principal coordinates analysis of Bray-Curtis dissimilarity of bacterial beta diversity (p = 0.7846, PERMANOVA; p = 0.3957, PERMDISP).

**Figure S6.**
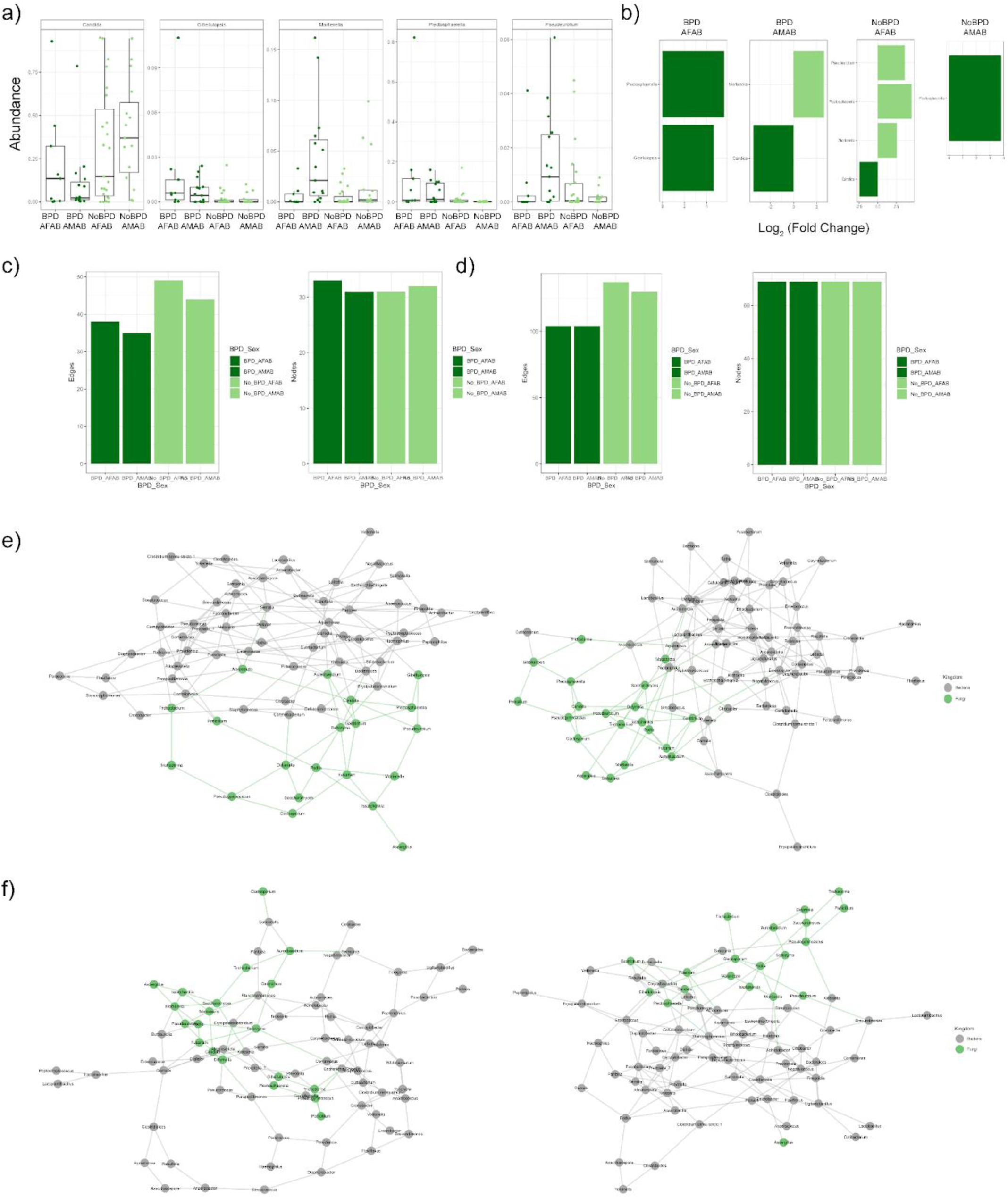
Microbiota alterations by disease status and sex assigned at birth. (a-b) DESeq2 selected features of the mycobiota. (c) SPIEC-EASI network edges. (d) SPIEC-EASI network nodes. (e) SPIEC-EASI networks for NoBPD infants assigned male at birth (AMAB) or assigned female at birth (AFAB). (f) SPIEC-EASI networks for BPD infants.

**Figure S7.**
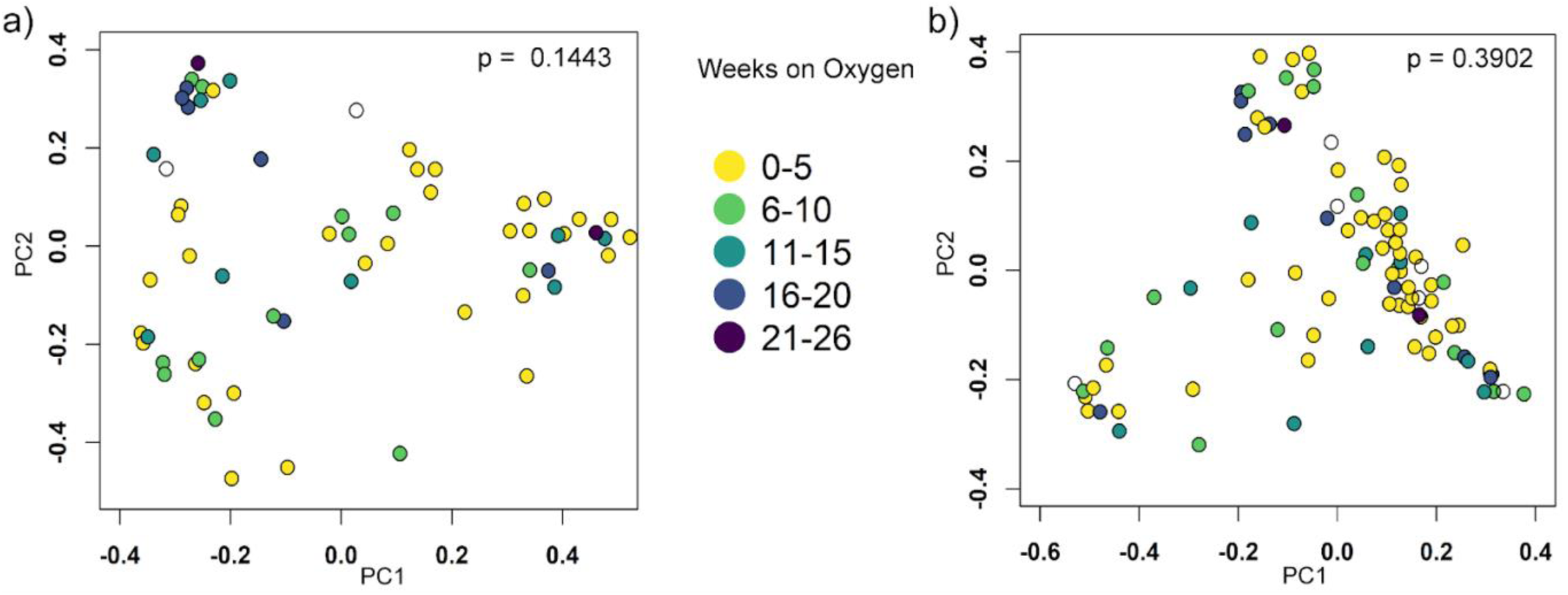
Oxygen concentration influences the composition of the intestinal microbiome. (A) PCoA of Bray-Curtis dissimilarity showing that the composition of the intestinal mycobiome is significantly associated with total weeks of exposure to oxygen (p = 0.1443, PERMANOVA). (B) PCoA of Bray-Curtis dissimilarity showing that the composition of the intestinal microbiome is not associated with total weeks of exposure to oxygen (p = 0.3902, PERMANOVA).

**Figure S8.**
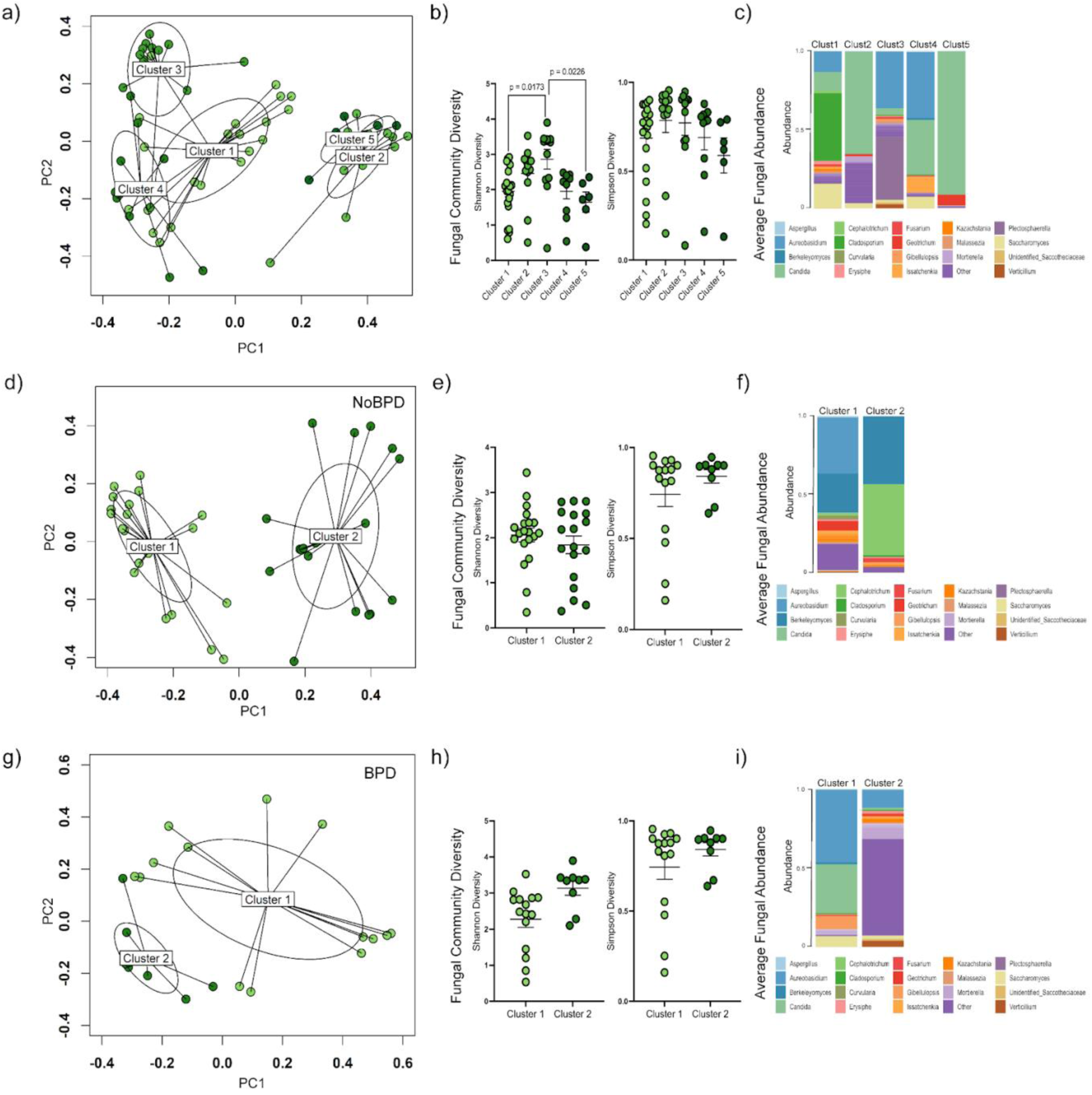
Unsupervised clustering analysis of heterogeneity in the mycobiome. (a) Dirichlet-Multinomial Mixture (DMM) model of the mycobiome. (b) Fungal community diversity analyses indicate higher microbial richness in Cluster 3. (c) Relative abundance of fungi in each DMM cluster. (d-f) A two-cluster DMM model in NoBPD infants, with the highest richness in Cluster 2. (g-i) A two-cluster DMM model in BPD infants with the highest microbial richness in Cluster 2. Clust1-5, Cluster 1-5. Ellipses indicate the 95% confidence interval.

**Figure S9.**
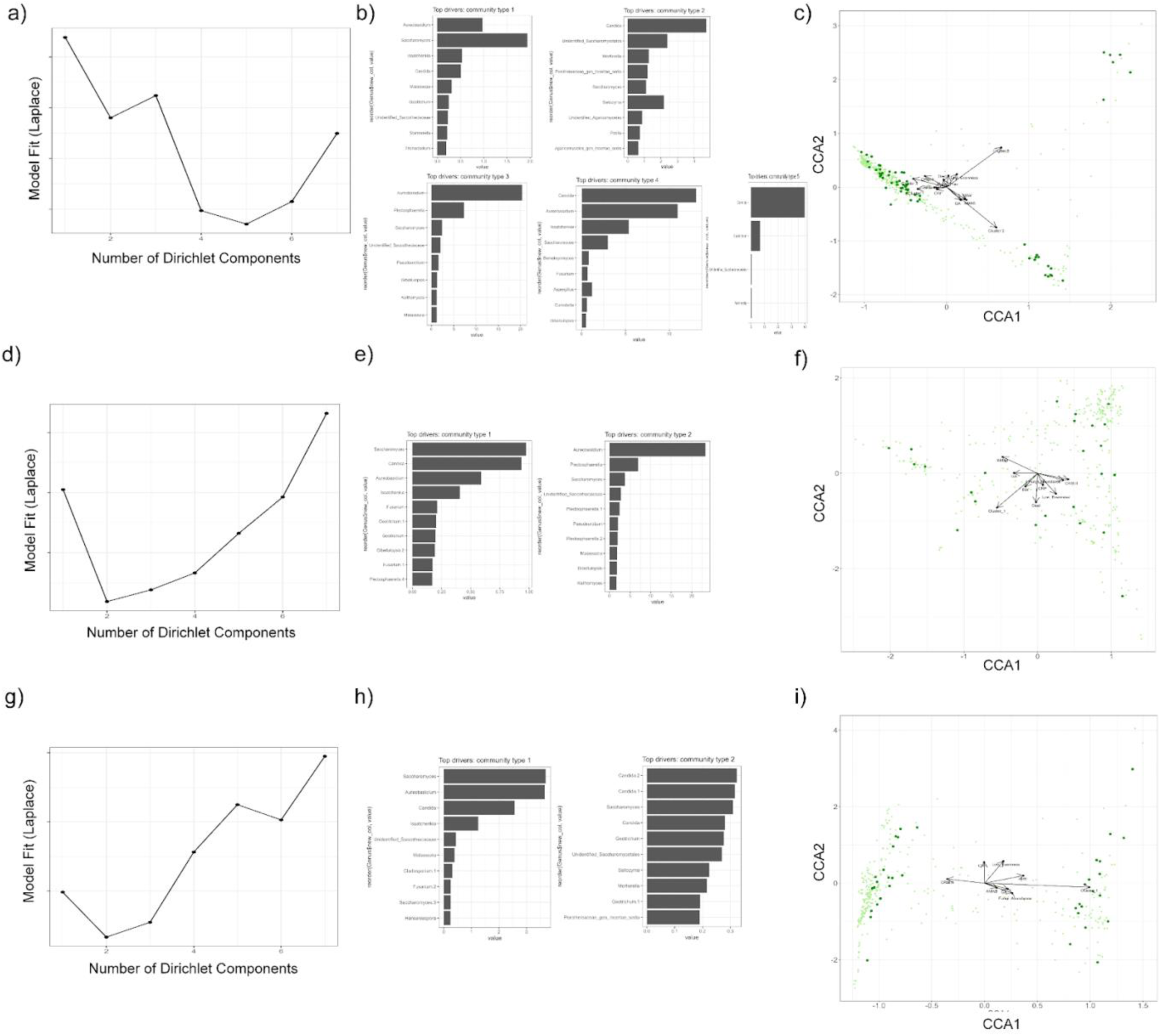
Dirichlet-Multinomial Mixture (DMM) modeling of the mycobiome. (a) Laplace analysis of the mycobiome indicates optimal model fit points with 5 clusters. (b) Variable importance analysis of the fungal DMM model. (c) Canonical Correspondence Analysis (CCA) for the DMM model. (d) Laplace analysis of mycobiome samples from NoBPD infants shows an optimal fit point at 2 clusters. (e) Variable importance analysis of the NoBPD DMM model. (f) CCA for the NoBPD DMM model. (g) Laplace analysis of mycobiome samples from BPD infants shows an optimal fit point at 2 clusters. (h) Variable importance analysis of the BPD DMM model. (i) CCA for the BPD DMM model.

**Figure S10.**
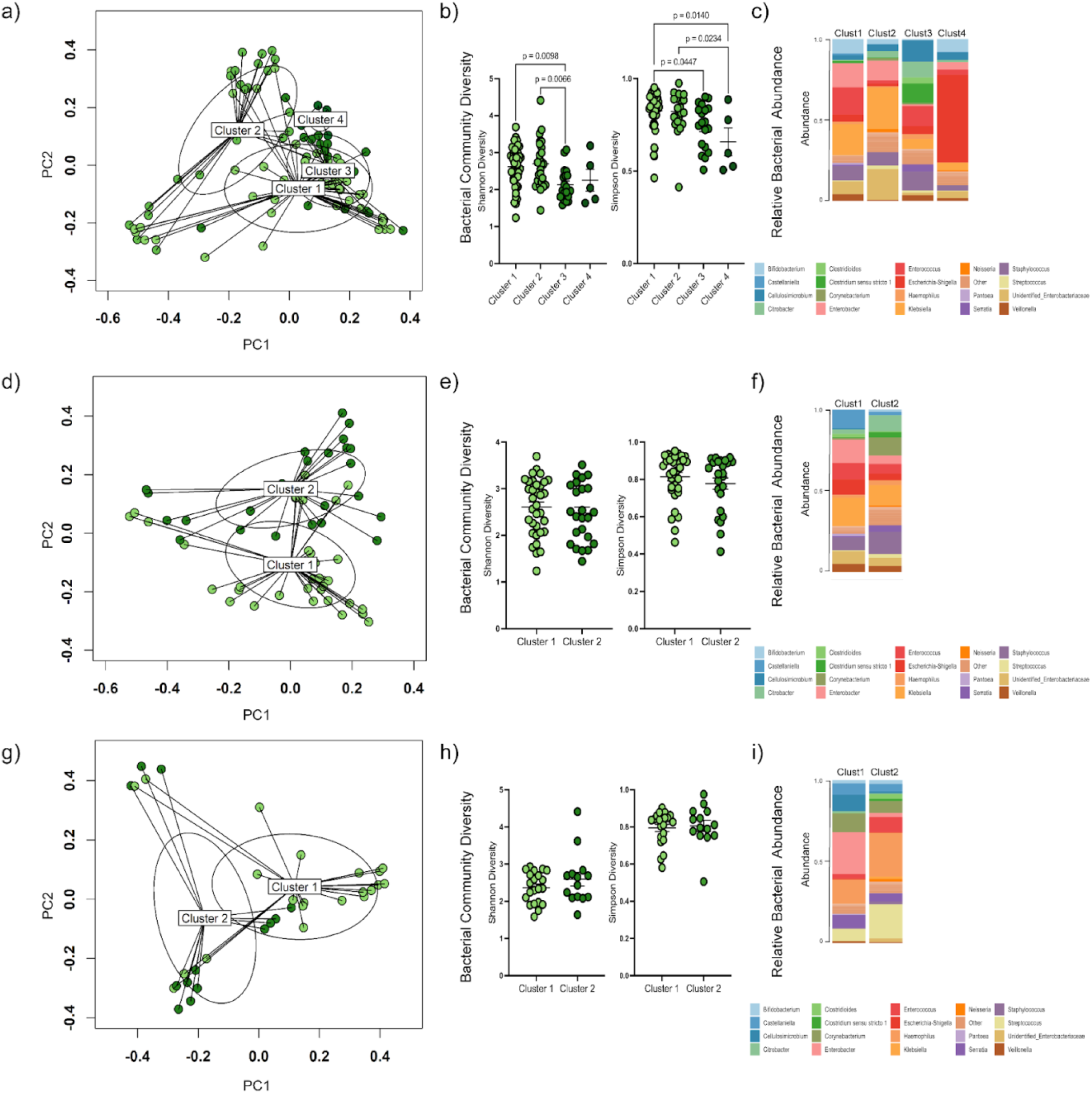
Dirichlet-Multinomial Mixture (DMM) modeling of the bacterial microbiome. (a) Principal Coordinates Analysis (PCoA) of the bacterial microbiome DMM model. (b) Alpha diversity of the bacterial microbiome DMM model. (c) Relative abundance of the bacterial microbiome in the DMM components. (d) PCoA of the DMM model of the bacterial microbiome of infants with NoBPD. (e) Alpha diversity of the multikingdom microbiome. (f) Relative abundance of the bacterial microbiome in the NoBPD DMM model. (g) PCoA of the DMM model of the bacterial microbiome of infants with BPD. (h) Alpha diversity of the bacterial microbiome. (i) Relative abundance of the multikingdom microbiome in the BPD DMM model. Ellipses indicated the 95% confidence interval.

**Figure S11.**
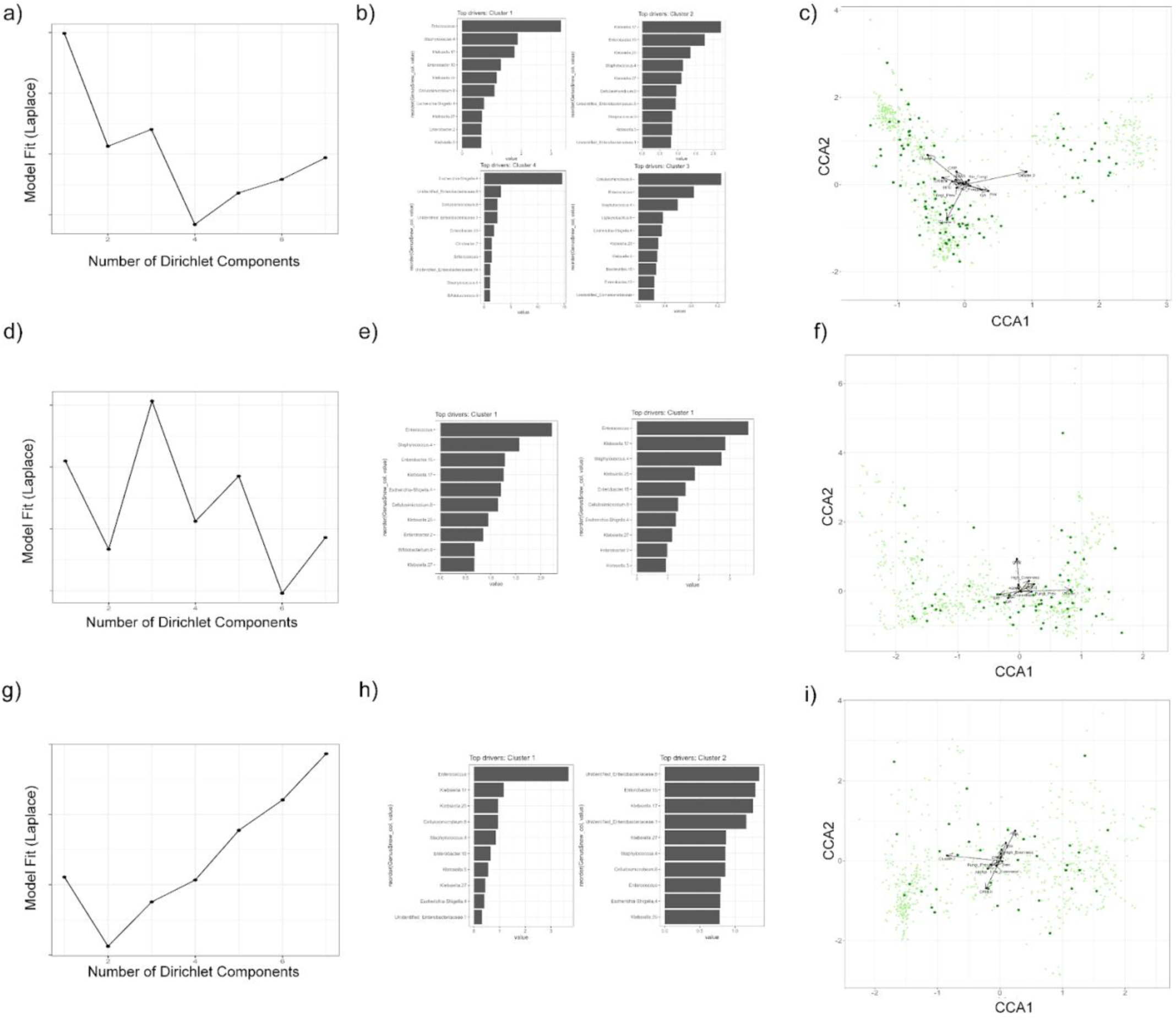
DMM modeling of the bacterial microbiome. (a) Laplace analysis of the mycobiome indicates an optimal model fit point with four components. (b) Variable importance analysis of the multikingdom microbiome DMM model. (c) Canonical Correspondence Analysis (CCA) for the multikingdom microbiome DMM model. (d) Laplace analysis of multikingdom microbiome samples from NoBPD infants showing an optimal fit point at three components. (e) Variable importance analysis of the NoBPD DMM model. (f) CCA for the NoBPD DMM model. (g) Laplace analysis of multikingdom microbiome samples from BPD infants showing an optimal fit point at two components. (h) Variable importance analysis of the BPD DMM model. (i) CCA for the BPD DMM model.

**Figure S12.**
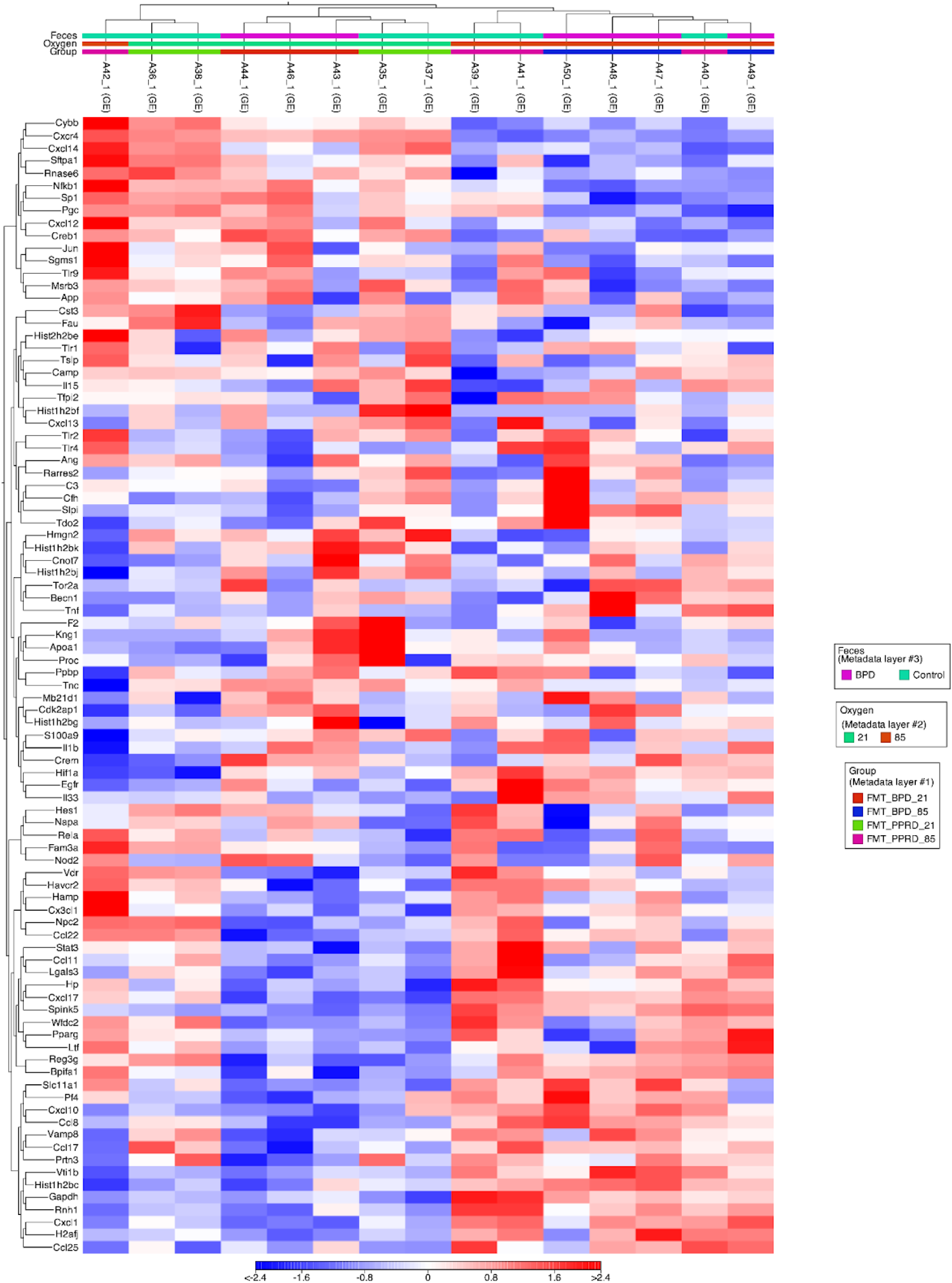
Lung antimicrobial peptide expression. Heatmap of antimicrobial peptides.

**Figure S13.**
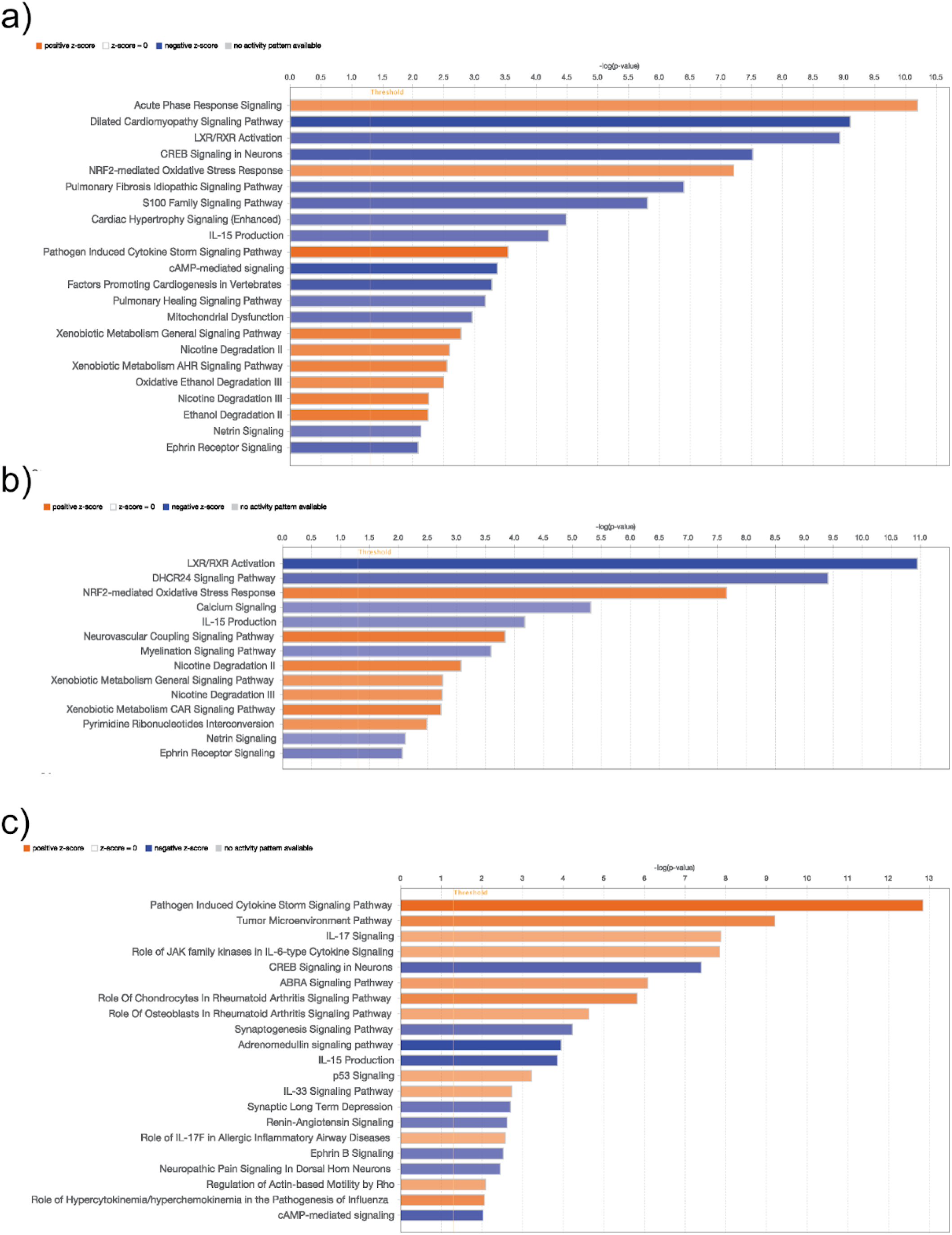
Ingenuity pathway analysis. (A) Pathways in BPD-FMT mice. (B) Pathways in NoBPD-FMT mice. (C) Pathways in specific pathogen-free (SPF) mice.

**Table S1.** Cohort characteristics and demographics.

|  | NoBPD | BPD | n | p |
| --- | --- | --- | --- | --- |
| Median (IQR) or n (%) | 64 | 38 |  |  |
| Gestational age (weeks) | 29.0 (27.6 - 30.8) | 25.7 (25 - 27.8) | 102 | < 0.001 |
| Birthweight (g) | 1145 (840 - 1330) | 775 (670 - 860) | 102 | < 0.001 |
| Female sex assigned at birth | 37/64 (57.8%) | 14/38 (36.8%) | 102 | 0.041 |
| Antenatal Steroids (No) | 8/47 (17%) | 4/30 (13.3%) | 77 | 0.33 |
| Surfactant (None) | 27/62 (43.5%) | 1/38 (2.6%) | 100 | <0.001 |
| Apgar score (1 min) | 6 (3 - 7) | 4 (2 - 5) | 99 | < 0.001 |
| Apgar score (5 min) | 7 (6 - 8) | 6 (5 - 7) | 99 | < 0.001 |
| Small for gestational age* | 6/64 (9.4%) | 8/38 (21.1%) | 102 | 0.098 |
| Cesarean section (1) | 28/47 (59.6%) | 20/30 (66.7%) | 77 | 0.531 |
| Prolonged rupture of membranes** (1) | 7/47 (15%) | 5/30 (16.7%) | 77 | 0.834 |
| Critical Risk Index for Babies II (CRIB II) score | 7 (4 - 9) | 12 (9 - 14) | 95 | < 0.001 |
| Prenatal antibiotic exposure | 22/47 (46.8%) | 15/30 (50%) | 77 | 0.785 |
| Duration of prenatal antibiotic exposure (d) | 1 (0 - 1) | 0 (0 - 1) | 60 | 0.542 |
| Postnatal antibiotic exposure | 55/64 (85.9%) | 38/38 (100%) | 102 | 0.015 |
| Duration of postnatal antibiotic exposure (d) | 2 (1 - 2) | 2 (1 - 4) | 96 | 0.016 |
| Endotracheal intubation | 41/62 (66.1%) | 37/37 (100%) | 99 | < 0.001 |
| Ventilation duration (d) | 1 (0 - 4) | 34 (22 - 50) | 94 | < 0.001 |
| Non-invasive ventilation duration (d) | 5 (1 - 17) | 14 (1 - 38) | 94 | 0.053 |
| Oxygen exposure duration (d) | 13 (4 - 41) | 92 (30 - 119) | 94 | < 0.001 |
| Death | 1/63 (1.6%) | 6/38 (15.8%) | 101 | 0.006 |

Data are presented as median (interquartile range) or n (%) with significance testing by Mann-Whitney U or Pearson chi-squared test, as appropriate.
BPD, Bronchopulmonary dysplasia; NoBPD, Post-prematurity respiratory disease. \* < 3rd percentile, ‡ > 18 h, †NICHD Neonatal Outcomes Estimator

**Table S2.**
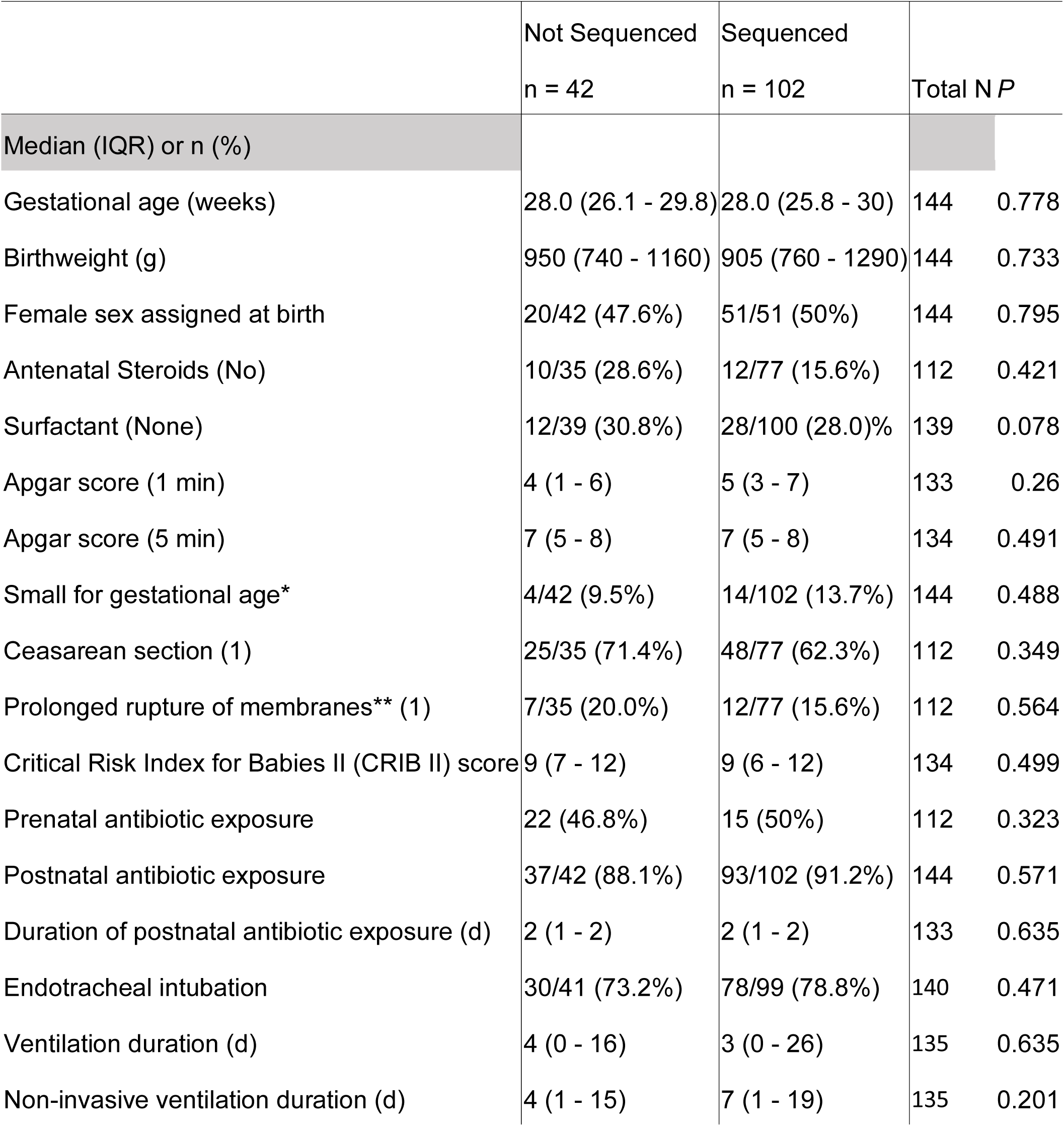
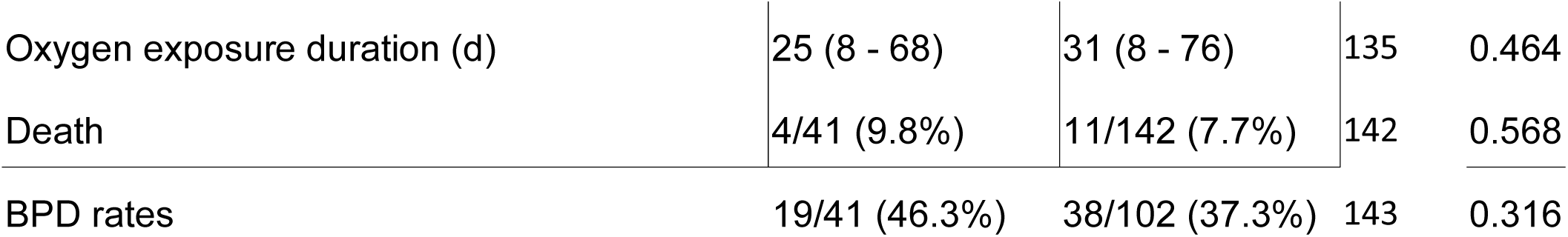
Characteristics and demographics of infants without a stool for analysis.

**Table S3.**
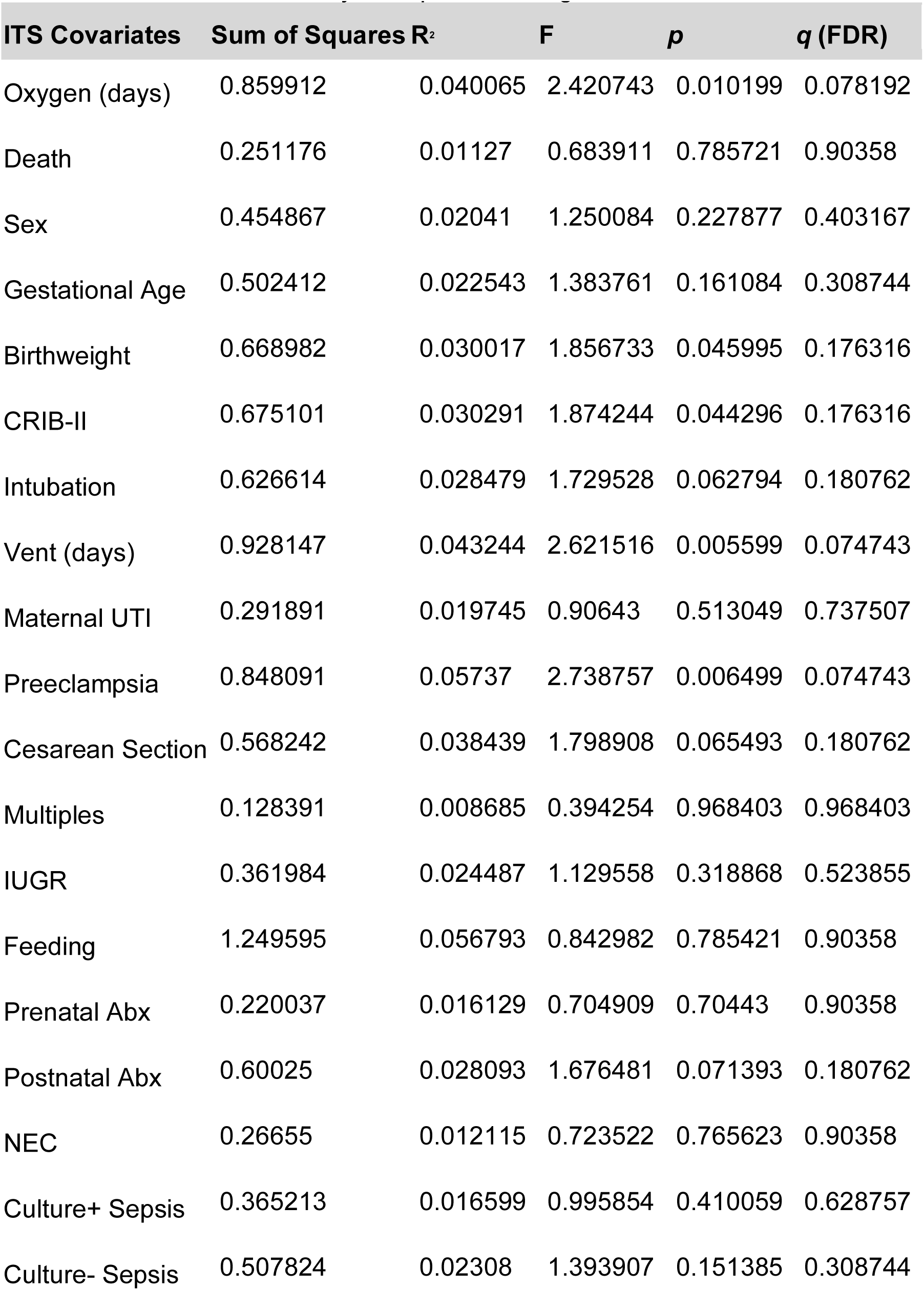
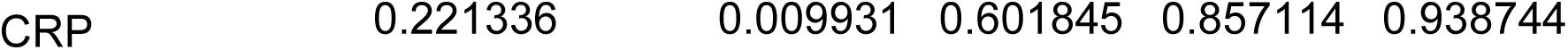
PERMANOVA analysis of potential fungal covariates.

**Table S4.** PERMANOVA analysis of potential bacterial covariates.

| Bacterial Covariates | Df | Sum of Sqs | R<sup>2</sup> | F | p | q (FDR) |
| --- | --- | --- | --- | --- | --- | --- |
| Death | 1 | 0.174478 | 0.005424 | 0.496284 | 0.986401 | 0.986401 |
| Sex | 1 | 0.259828 | 0.008077 | 0.74103 | 0.784622 | 0.859347 |
| Gestational age | 1 | 0.583988 | 0.018155 | 1.682631 | 0.037496 | 0.172483 |
| Birthweight | 1 | 0.738424 | 0.022956 | 2.138061 | 0.005999 | 0.10349 |
| CRIB-II | 1 | 0.630824 | 0.019611 | 1.820278 | 0.022698 | 0.130512 |
| Feeding | 4 | 1.383682 | 0.044219 | 0.994685 | 0.480952 | 0.655051 |
| Oxygen (days) | 1 | 0.526242 | 0.017615 | 1.524152 | 0.072893 | 0.20468 |
| Vent (days) | 1 | 0.368035 | 0.01232 | 1.060221 | 0.373363 | 0.572489 |
| Multiples | 1 | 0.31171 | 0.013528 | 0.946218 | 0.484952 | 0.655051 |
| IUGR | 1 | 0.362514 | 0.015733 | 1.102904 | 0.322868 | 0.530426 |
| Maternal UTI | 1 | 0.297311 | 0.012903 | 0.901938 | 0.554045 | 0.663484 |
| Preeclampsia | 1 | 0.439966 | 0.019094 | 1.343129 | 0.161984 | 0.312769 |
| Cesarean section | 1 | 0.21574 | 0.009363 | 0.652141 | 0.838816 | 0.876944 |
| Prenatal Abx | 1 | 0.557406 | 0.026794 | 1.734477 | 0.008999 | 0.10349 |
| Postnatal Abx | 1 | 0.451244 | 0.014831 | 1.29471 | 0.163184 | 0.312769 |
| NEC | 1 | 0.309181 | 0.009881 | 0.888147 | 0.576942 | 0.663484 |
| Intubation | 1 | 0.537091 | 0.017164 | 1.55427 | 0.069293 | 0.20468 |
| Culture+ sepsis | 1 | 0.474332 | 0.015158 | 1.369859 | 0.123588 | 0.284252 |
| Culture- sepsis | 1 | 0.668253 | 0.021356 | 1.942117 | 0.015398 | 0.118055 |
| CRP | 1 | 0.329732 | 0.01037 | 0.943039 | 0.512649 | 0.655051 |

**Table S5.**
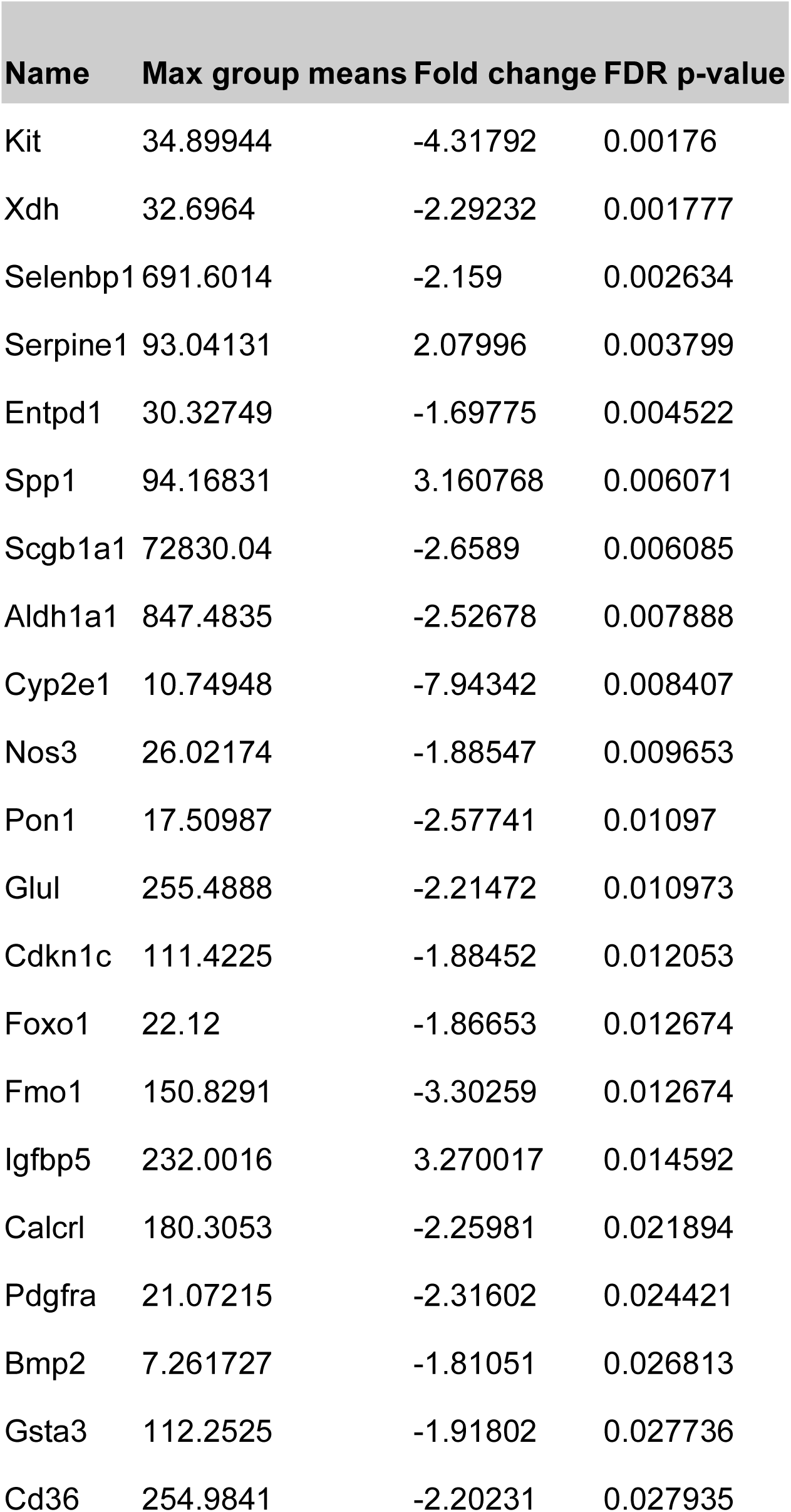
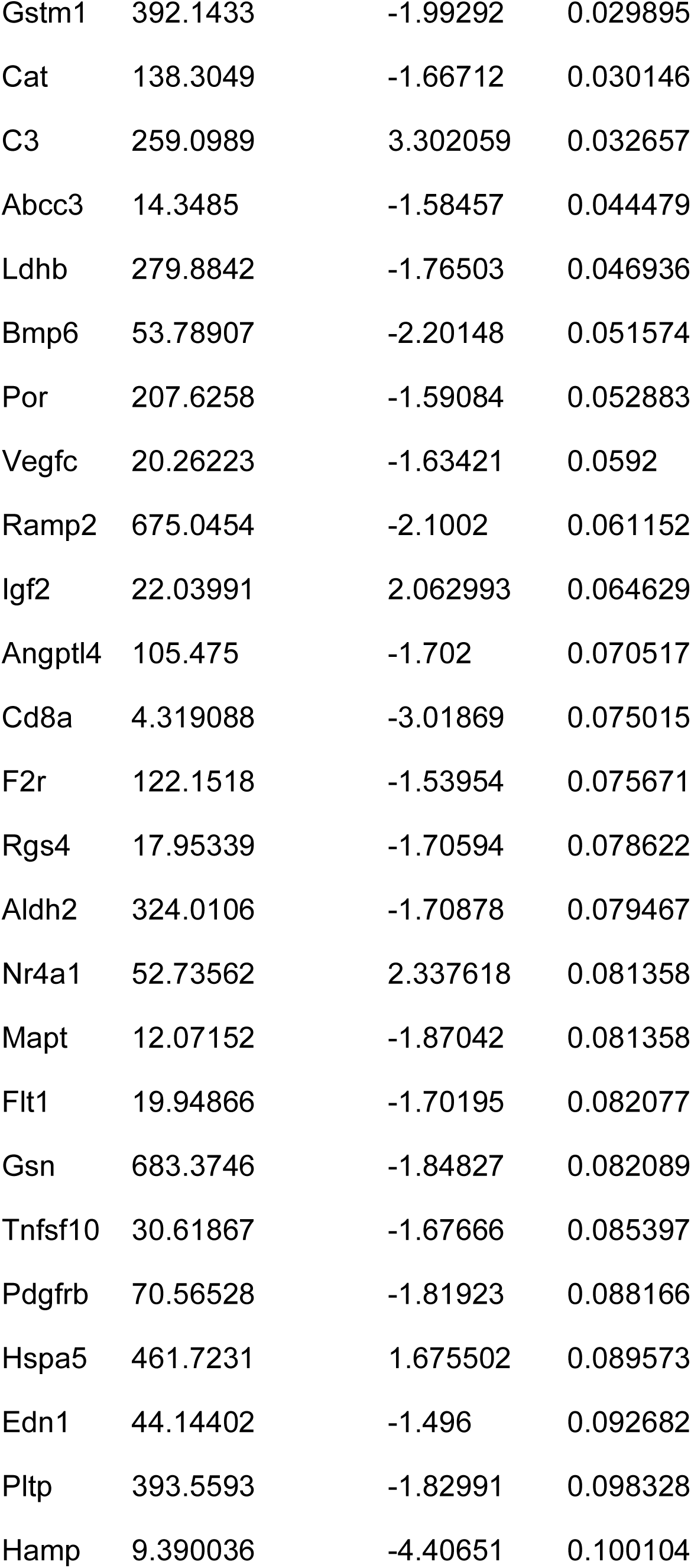
Acute lung injury-associated genes differentially expressed in the lungs of BPD-FMT of NoBPD-FMT mice after hyperoxia-exposure.

**Table S6.** Lectins and other fungal-associated genes.

| Name | Normoxia max group means | Fold change | FDR p-value | Hyperoxia max group means | Fold change | FDR p-value |
| --- | --- | --- | --- | --- | --- | --- |
| Reg3g | 125.7378 | -10.4239 | 0.003688 | 162.3476 | 1.02955 | 0.992017 |
| Clec5a | 1.08385 | 1.087852 | 0.998991 | 2.712433 | 1.686155 | 0.418715 |
| Clec4d | 3.715903 | 1.193202 | 0.998991 | 8.32751 | 1.733554 | 0.313419 |
| Clec10a | 6.247333 | -2.29367 | 0.528492 | 6.11558 | -1.05898 | 0.96846 |
| Colec11 | 3.914303 | -1.52484 | 0.998991 | 4.594983 | 2.712321 | 0.115261 |
| Cd302 | 49.55867 | -1.06473 | 0.998991 | 81.59808 | 1.272356 | 0.526733 |
| Mrc1 | 17.6641 | -1.42096 | 0.998991 | 21.61832 | 1.057407 | 0.943666 |
| Olr1 | 3.512283 | -1.07582 | 0.998991 | 5.277159 | 1.164597 | 0.831115 |
| Clec7a | 31.76774 | -1.17945 | 0.998991 | 41.95802 | 1.386064 | 0.46829 |
| Clec12a | 6.16115 | -1.15362 | 0.998991 | 7.903554 | 1.191562 | 0.811886 |
| Clec11a | 8.423013 | -1.33604 | 0.998991 | 8.940581 | 1.384451 | 0.59313 |
| Clec3b | 365.5351 | -1.45895 | 0.906358 | 346.9807 | 1.054824 | 0.942974 |
| Sftpd | 802.4012 | -1.40873 | 0.998991 | 896.616 | -1.17443 | 0.768305 |
| Clec2l | 1.301569 | -1.14745 |  | 1.483454 | 1.178183 | 0.921223 |
| Pla2r1 | 4.668342 | 1.038428 | 0.998991 | 5.543161 | 1.070186 | 0.925283 |
| Clec1a | 35.80718 | -1.11514 | 0.998991 | 48.076 | -1.24911 | 0.638181 |
| Clec9a | 15.46983 | -1.45215 | 0.998991 | 15.75296 | -1.29182 | 0.63289 |
| Layn | 8.169843 | -1.13391 | 0.998991 | 9.311215 | -1.21783 | 0.653337 |
| Mrc2 | 96.92931 | -1.12035 | 0.998991 | 88.82867 | 1.156192 | 0.82189 |
| Klrd1 | 2.481198 | -1.00763 |  | 2.508489 | 1.181422 | 0.907133 |
| Vcan | 7.76357 | 1.374094 | 0.998991 | 7.116964 | 1.141553 | 0.898861 |
| Dgcr2 | 53.08562 | 1.12667 | 0.998991 | 48.12952 | -1.03824 | 0.952087 |
| Colec12 | 101.9319 | -1.03197 | 0.998991 | 90.5154 | 1.105169 | 0.864535 |
| Atrn | 11.93377 | 1.021454 | 0.998991 | 11.03505 | -1.10897 | 0.81083 |
| Pkd1 | 25.75555 | 1.208233 | 0.998991 | 22.74798 | -1.08504 | 0.894302 |
| Frem1 | 21.77959 | 1.210365 | 0.998991 | 16.5999 | 1.258854 | 0.692306 |
| Cd72 | 5.412205 | 1.243655 | 0.998991 | 7.491691 | -1.71869 | 0.436089 |
| Klrg1 | 1.821741 | -2.17122 |  | 0.771923 | -1.20194 | 0.92452 |
| Sftpa1 | 973.3715 | -1.26977 | 0.998991 | 995.9875 | -1.57053 | 0.330538 |
| Bcan | 1.006777 | -1.13803 | 0.998991 | 0.866189 | -1.20371 | 0.902223 |
| Cd93 | 106.9558 | 1.084508 | 0.998991 | 125.5295 | -1.65012 | 0.149331 |
| Atrnl1 | 16.04558 | 1.051649 | 0.998991 | 11.66754 | -1.02127 | 0.973212 |
| Cd69 | 1.811218 | 1.340058 |  | 1.5302 | -1.18015 | 0.923925 |
| Clec2d | 219.5001 | -1.18767 | 0.998991 | 152.6552 | -1.06741 | 0.952087 |
| Clec14a | 114.9554 | -1.33834 | 0.998991 | 132.072 | -1.99149 | 0.036064 |
| Clec12b | 4.053577 | 1.105698 | 0.998991 | 2.915567 | -1.11494 | 0.934748 |
| Clec1b | 5.444281 | 1.058832 | 0.998991 | 4.29238 | -1.21047 | 0.85957 |
| Thbd | 261.6676 | -1.12082 | 0.998991 | 259.302 | -1.67356 | 0.114563 |
| Prg2 | 8.766499 | -2.14827 | 0.998991 | 2.055661 | 1.309011 | 0.910762 |
| Sell | 7.236587 | 1.584014 | 0.998991 | 5.452646 | -1.77671 | 0.261235 |
| Cd248 | 106.8494 | 1.15852 | 0.998991 | 42.13223 | -1.20353 | 0.759667 |
| Mbl2 | 1.115486 | -1.74598 |  | 0.077049 | 1.439069 | 0.951451 |
| Slpi | 42.21049 | -1.56242 | 0.998991 | 109.266 | 3.009375 | 3.74E-05 |
| Hist1h2bc | 125.2418 | -1.0633 | 0.998991 | 195.9711 | 1.239957 | 0.50988 |
| Clec7a | 31.76774 | -1.17945 | 0.998991 | 41.95802 | 1.386064 | 0.46829 |
| Rarres2 | 387.2024 | -1.2211 | 0.998991 | 472.3454 | 1.744679 | 0.032897 |
| Senp8 | 1.319852 | 1.09805 | 0.998991 | 1.783109 | 1.418531 | 0.663881 |
| Tlr4 | 3.687062 | -1.0488 | 0.998991 | 5.190117 | -1.04613 | 0.949032 |
| Tlr2 | 6.261399 | -1.09921 | 0.998991 | 7.540487 | 1.112188 | 0.889589 |
| Hscb | 26.01099 | -1.23587 | 0.998991 | 25.40008 | 1.162098 | 0.817415 |
| Malt1 | 4.614531 | 1.090377 | 0.998991 | 4.951084 | 1.039782 | 0.964061 |
| Hif1a | 36.45999 | 1.205099 | 0.998991 | 39.2115 | 1.024301 | 0.972832 |
| Camp | 12.08179 | -1.3274 | 0.998991 | 9.823168 | 1.48817 | 0.715914 |
| Gps1 | 85.19948 | 1.189032 | 0.998991 | 85.20754 | 1.013453 | 0.983507 |
| Hist2h2be | 7.226456 | 1.001625 | 0.998991 | 7.375492 | -1.0413 | 0.969961 |
| Rel | 1.266513 | 1.633626 |  | 1.45833 | -1.17561 | 0.912653 |
| Colec12 | 101.9319 | -1.03197 | 0.998991 | 90.5154 | 1.105169 | 0.864535 |
| Hist1h2bf | 1.079463 | -5.85598 |  | 0.129765 | 5.356575 |  |
| Hist1h2bg | 3.325025 | 3.199163 |  | 2.524179 | 1.206217 | 0.931258 |
| Prg2 | 8.766499 | -2.14827 | 0.998991 | 2.055661 | 1.309011 | 0.910762 |

**Table S7.**
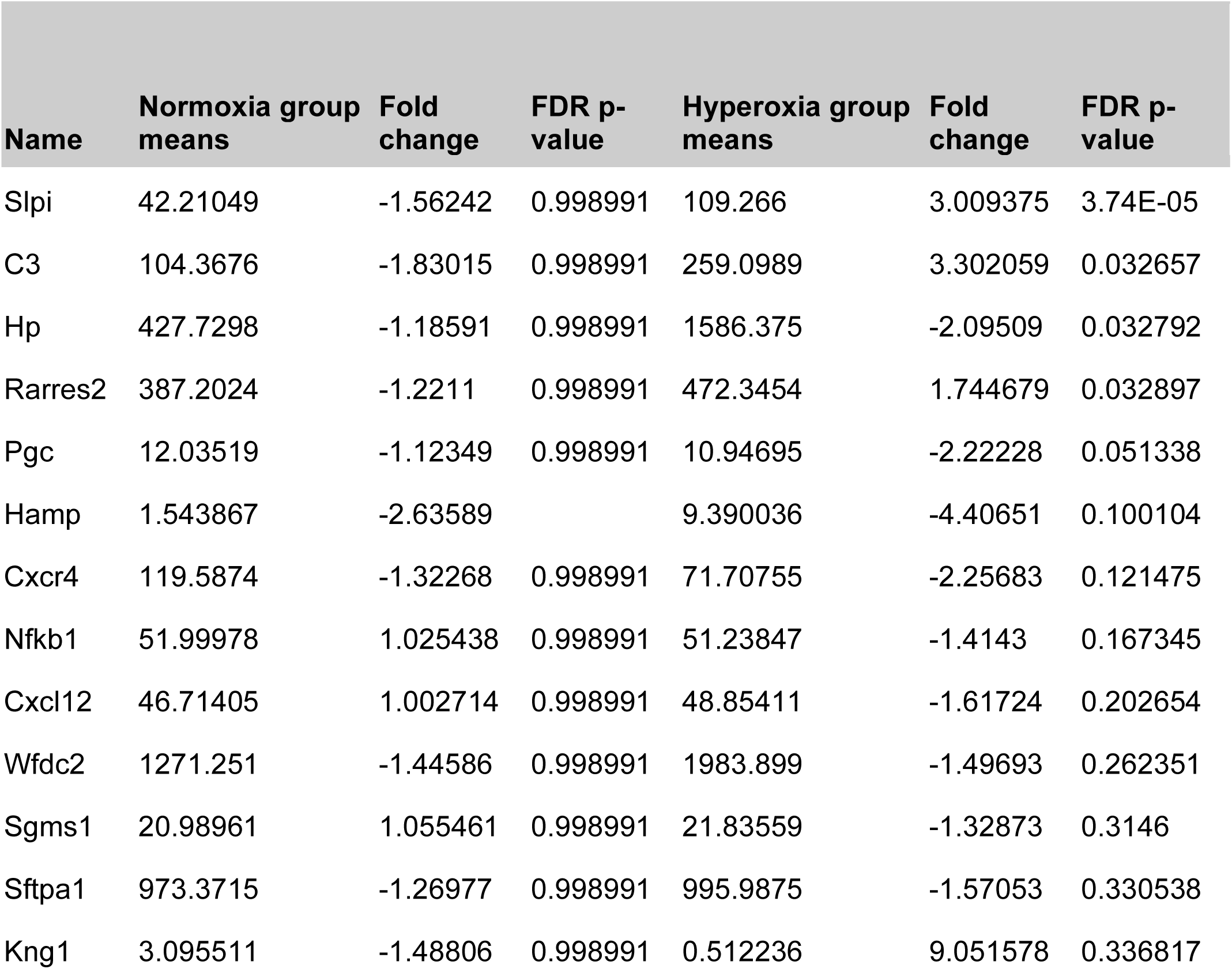
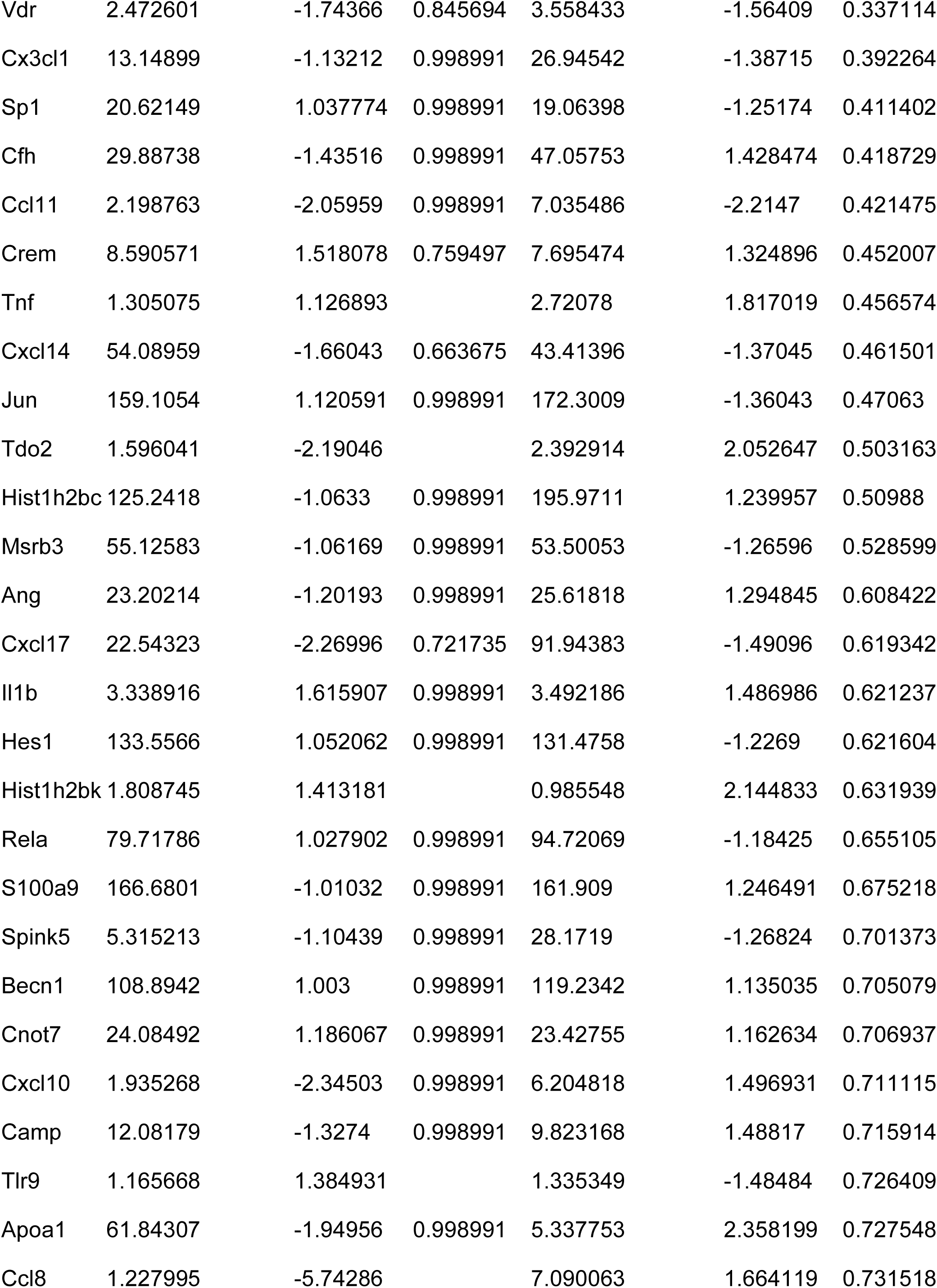
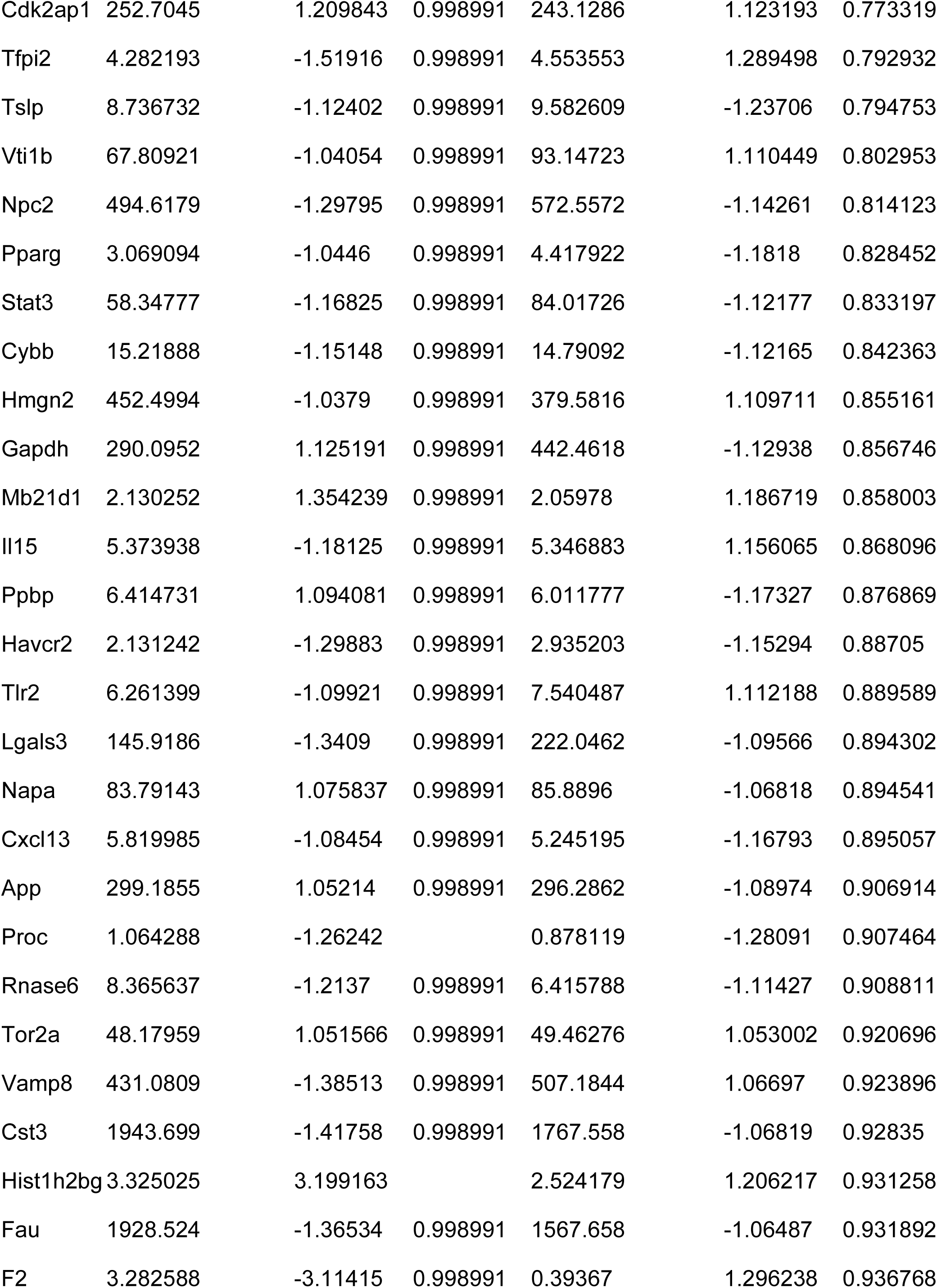
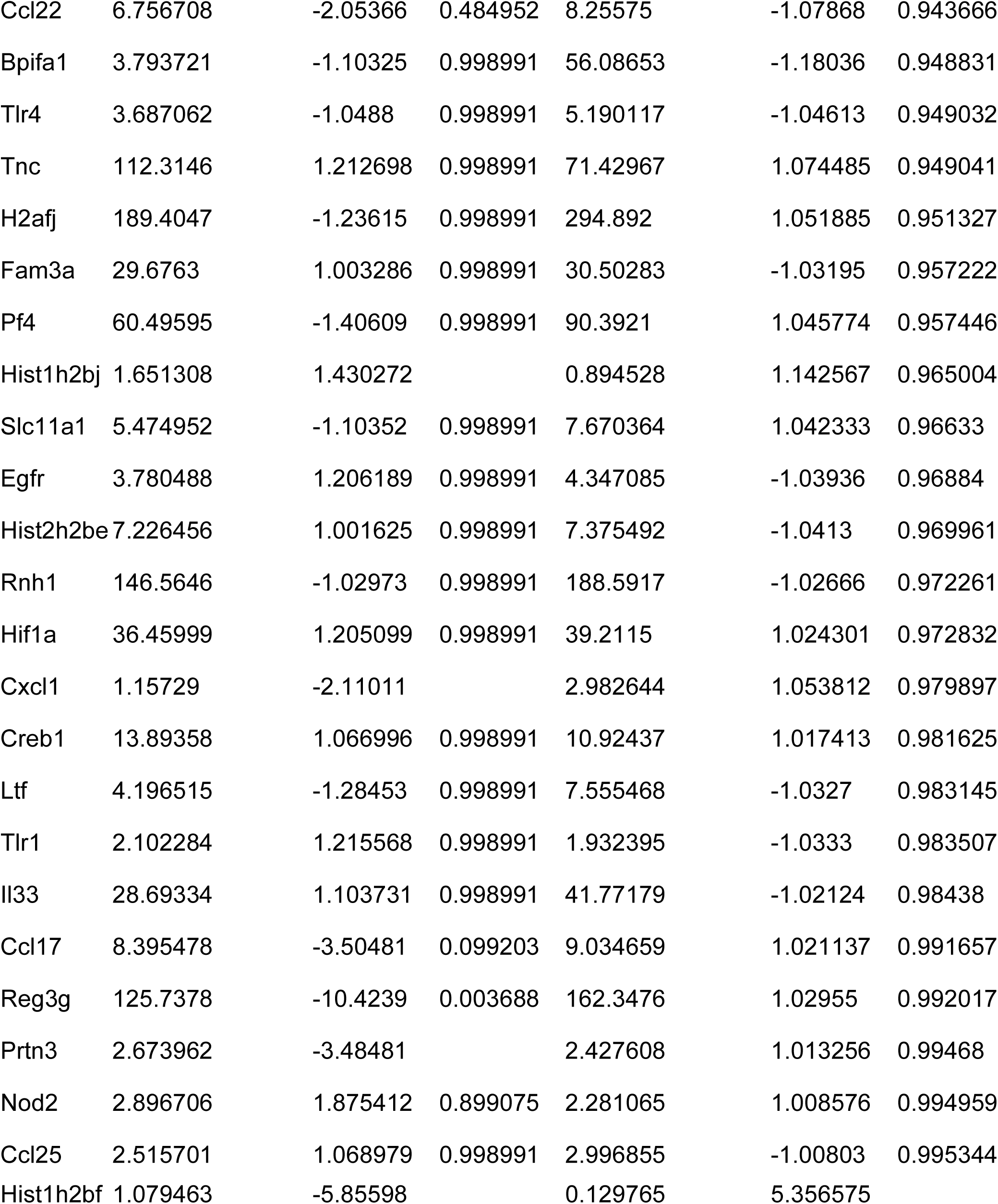
Antimicrobial peptides differentially expressed after FMT.

**Table S8.**
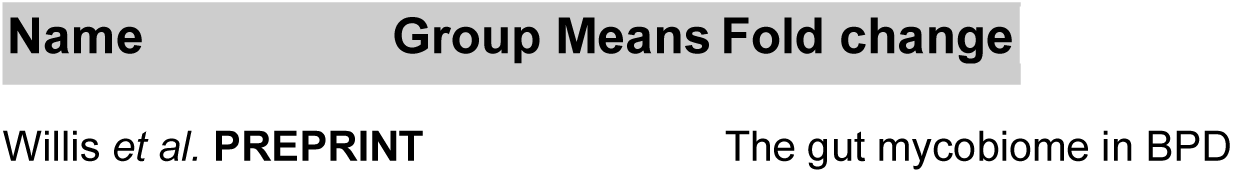
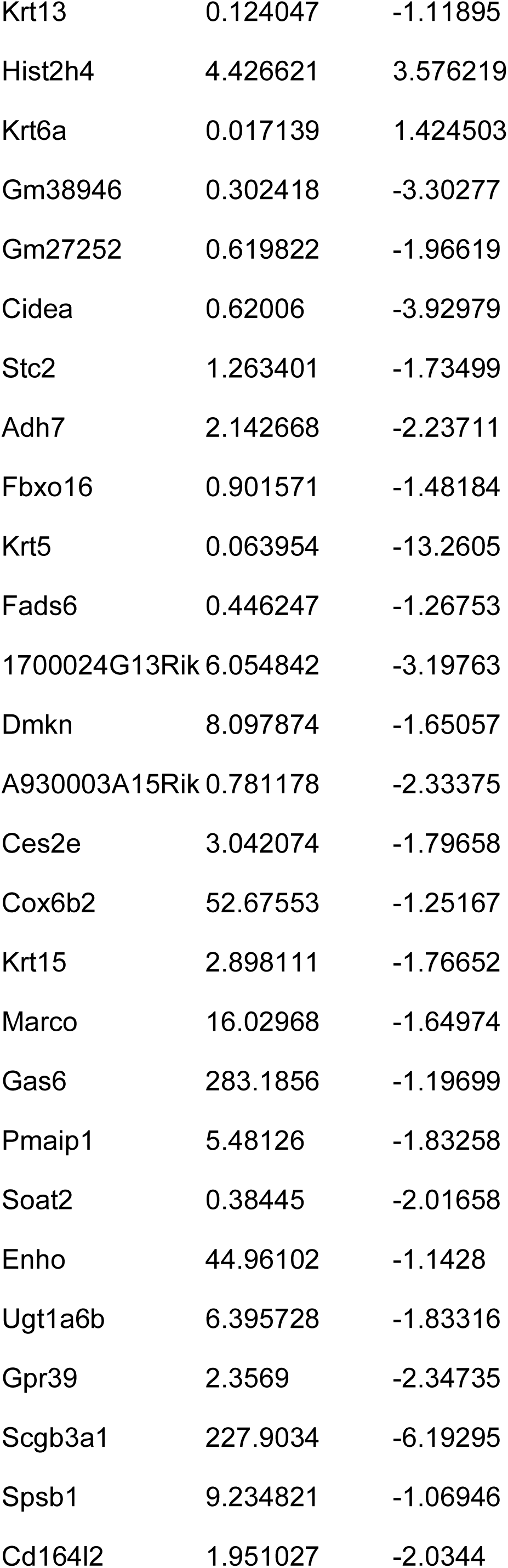
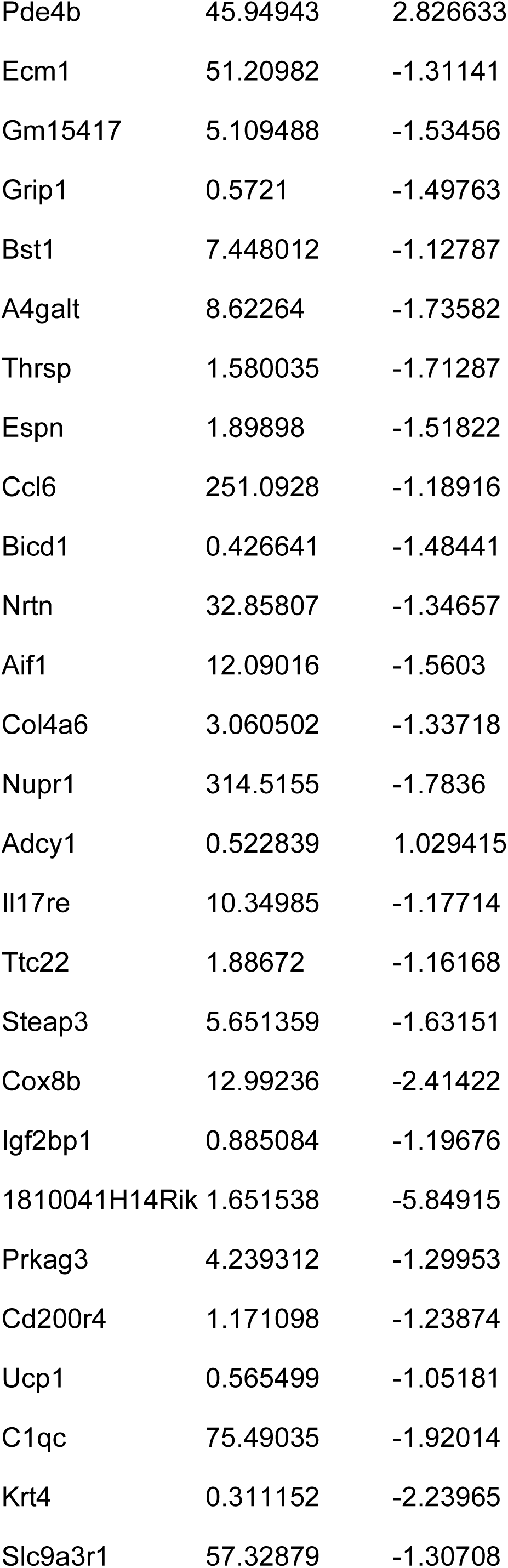
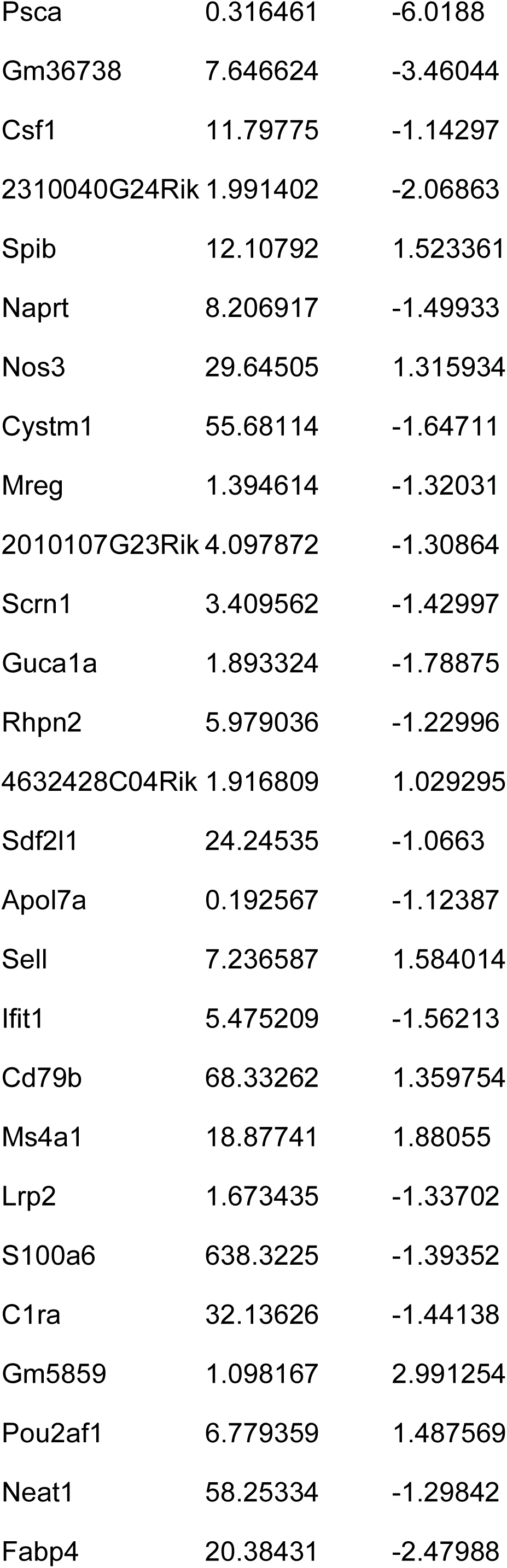
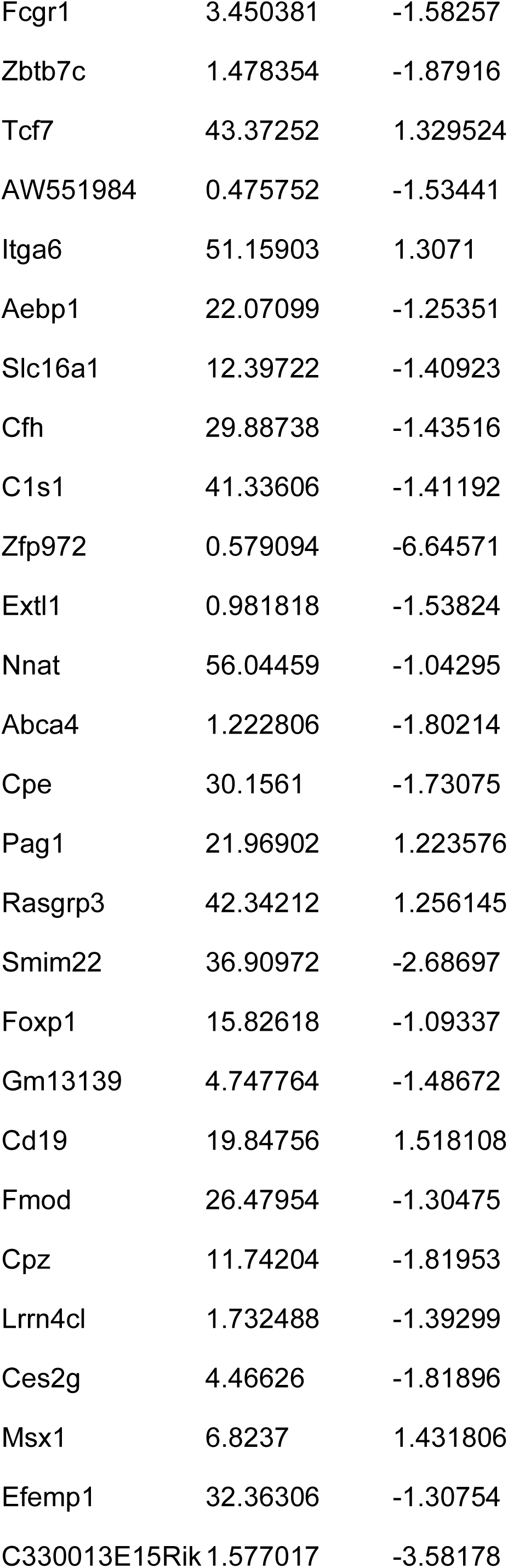
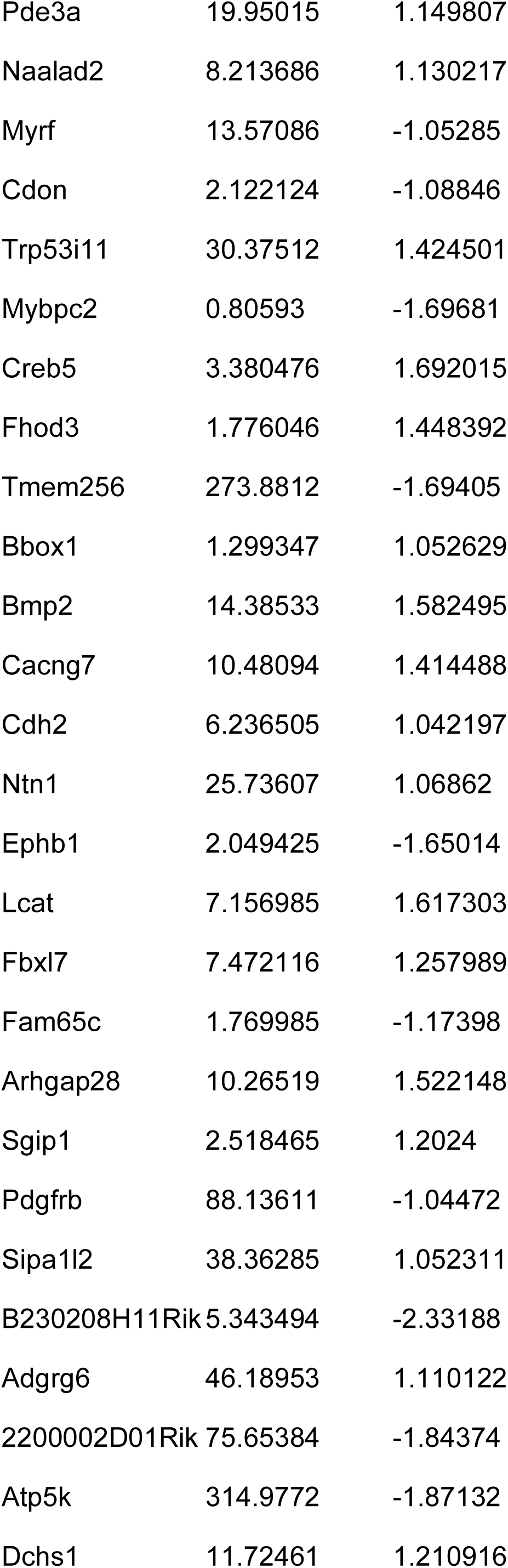
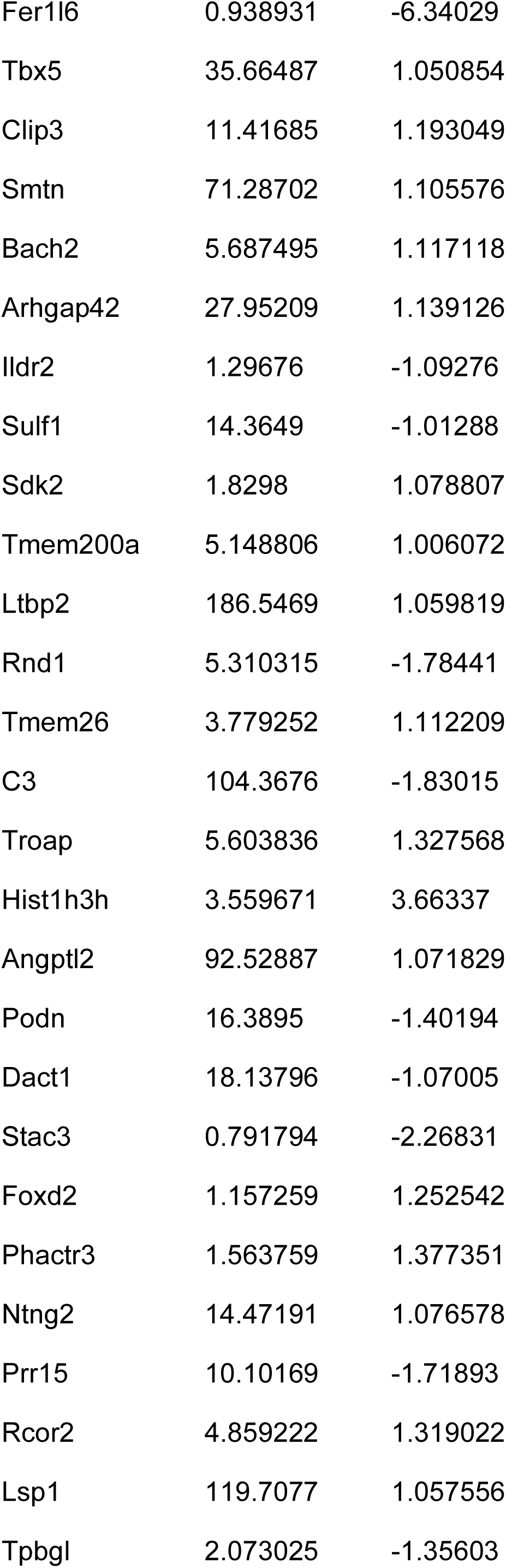
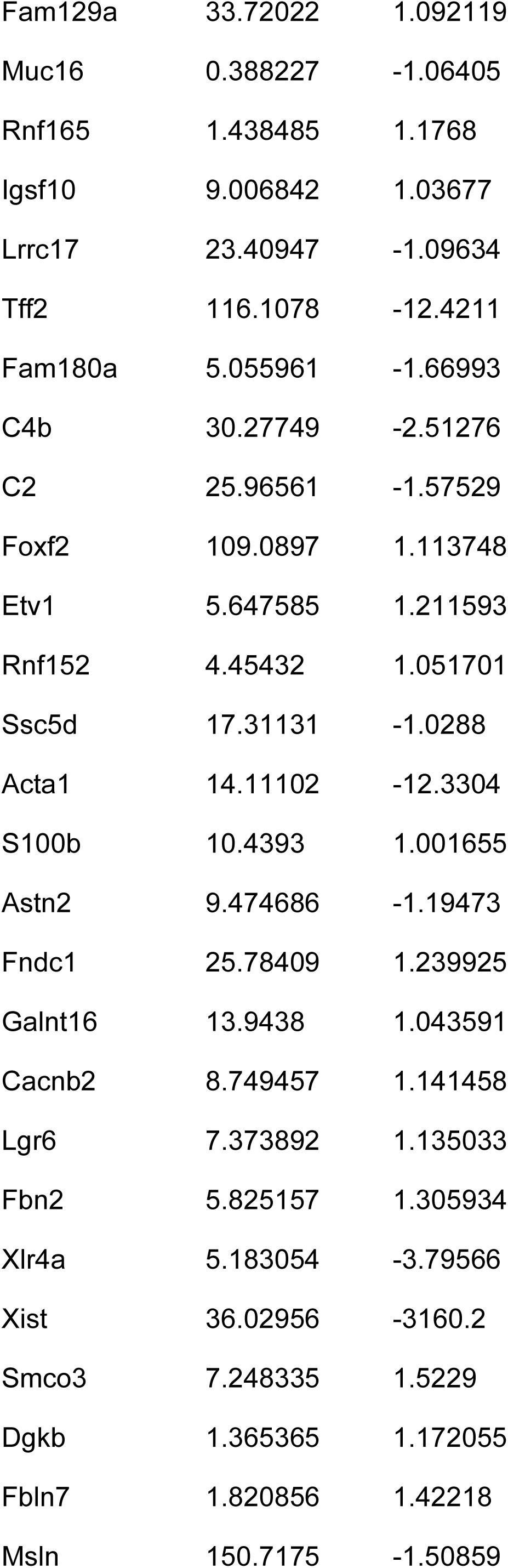
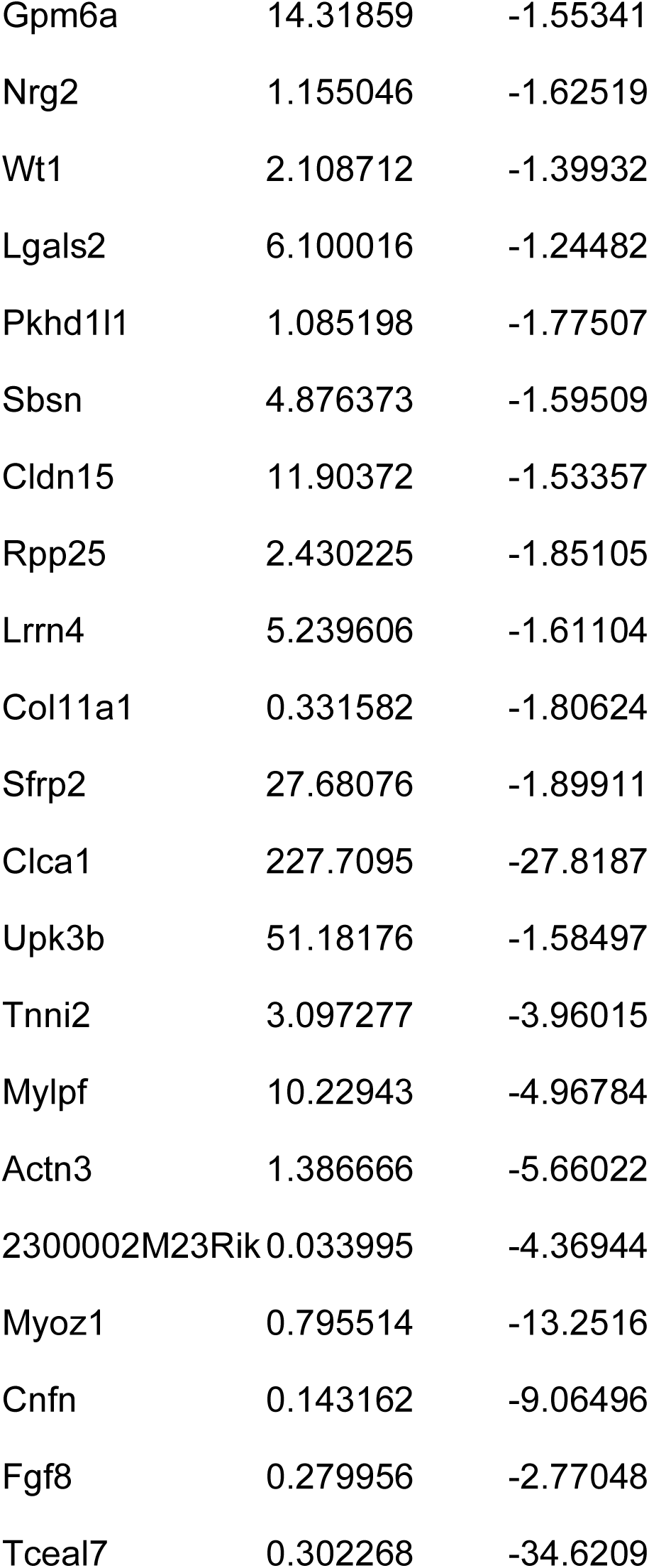
Genes uniquely regulated in the lungs of BPD-FMT mice as compared to conventional specific pathogen-free controls.

**Table S9.**
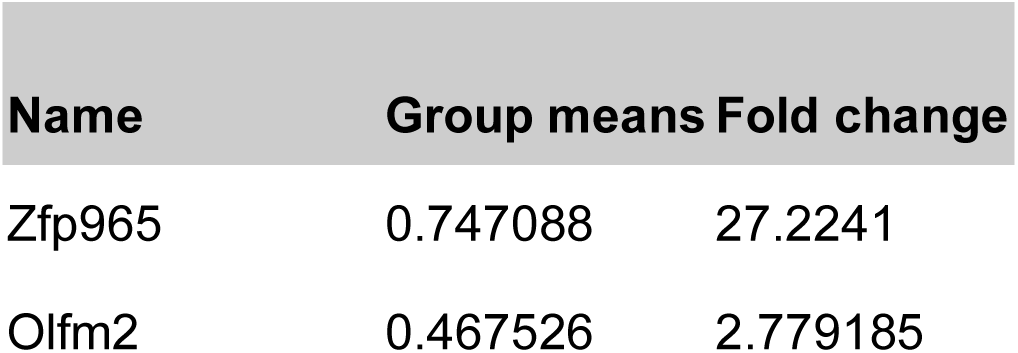
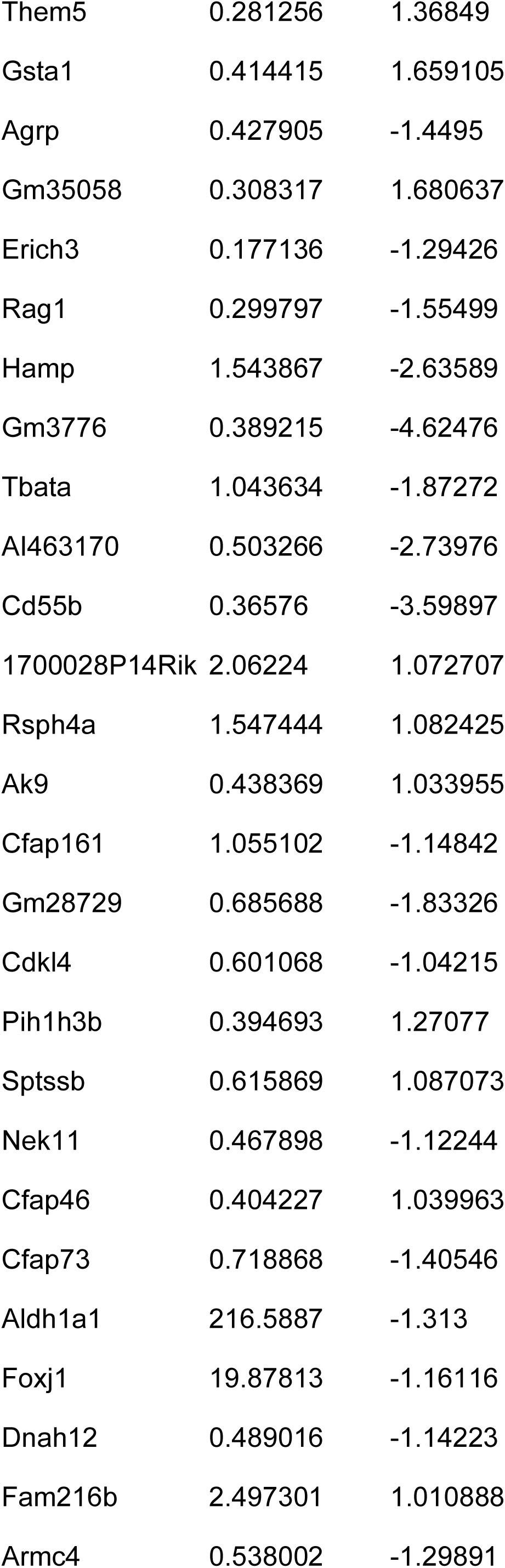
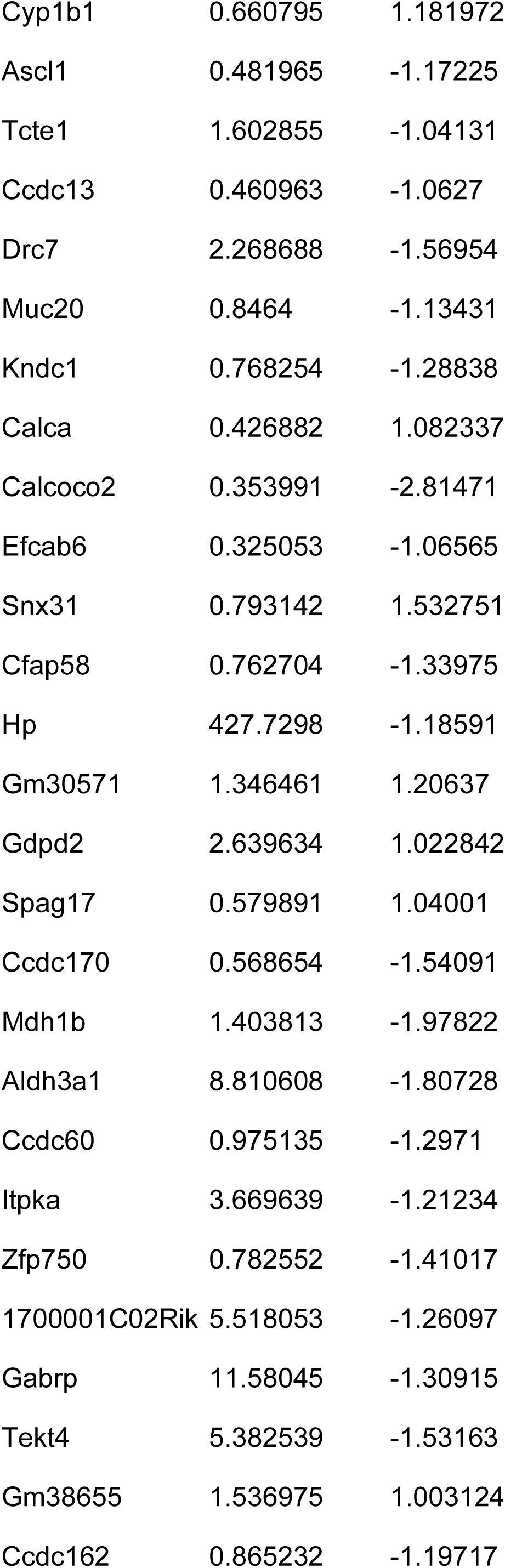
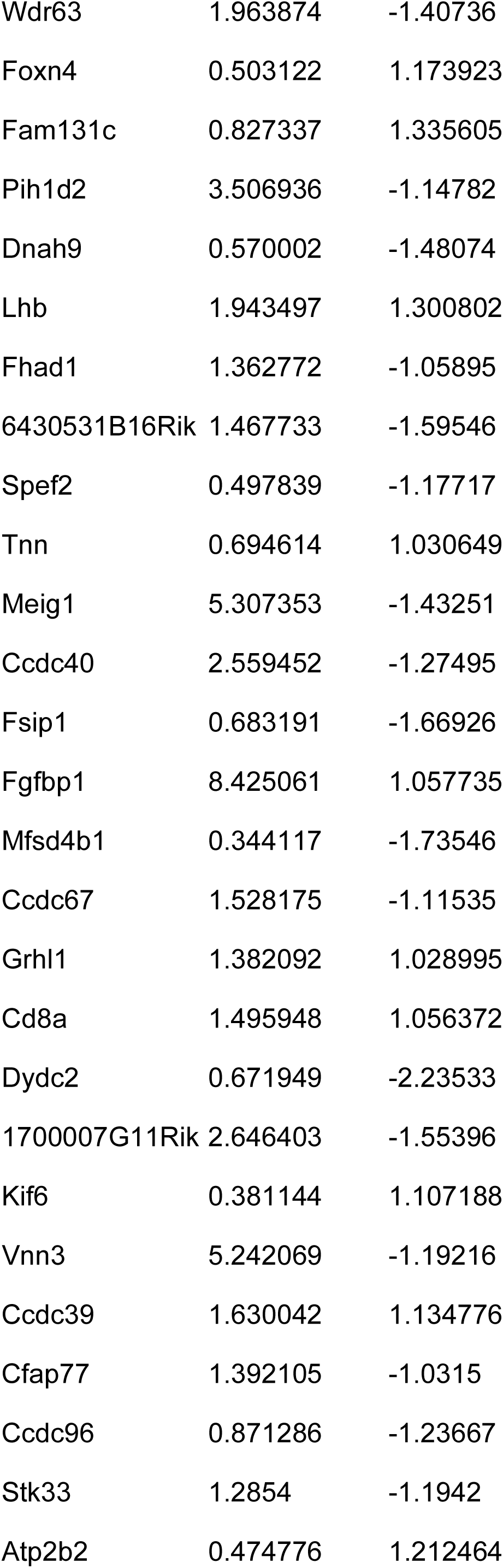
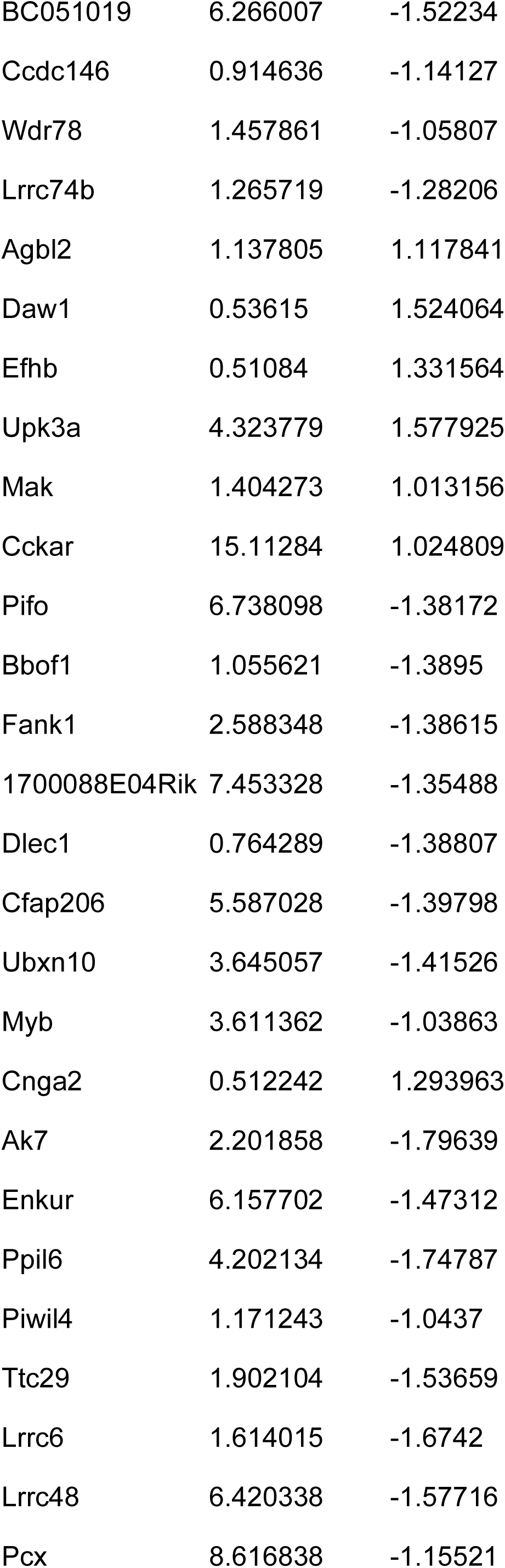
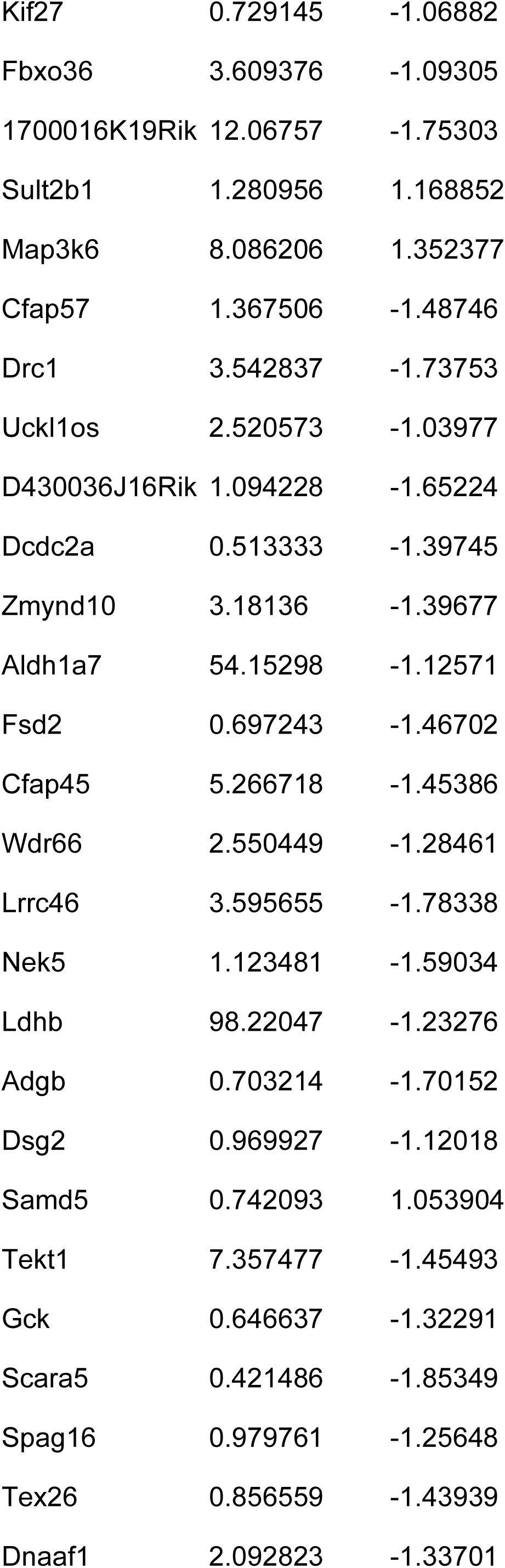
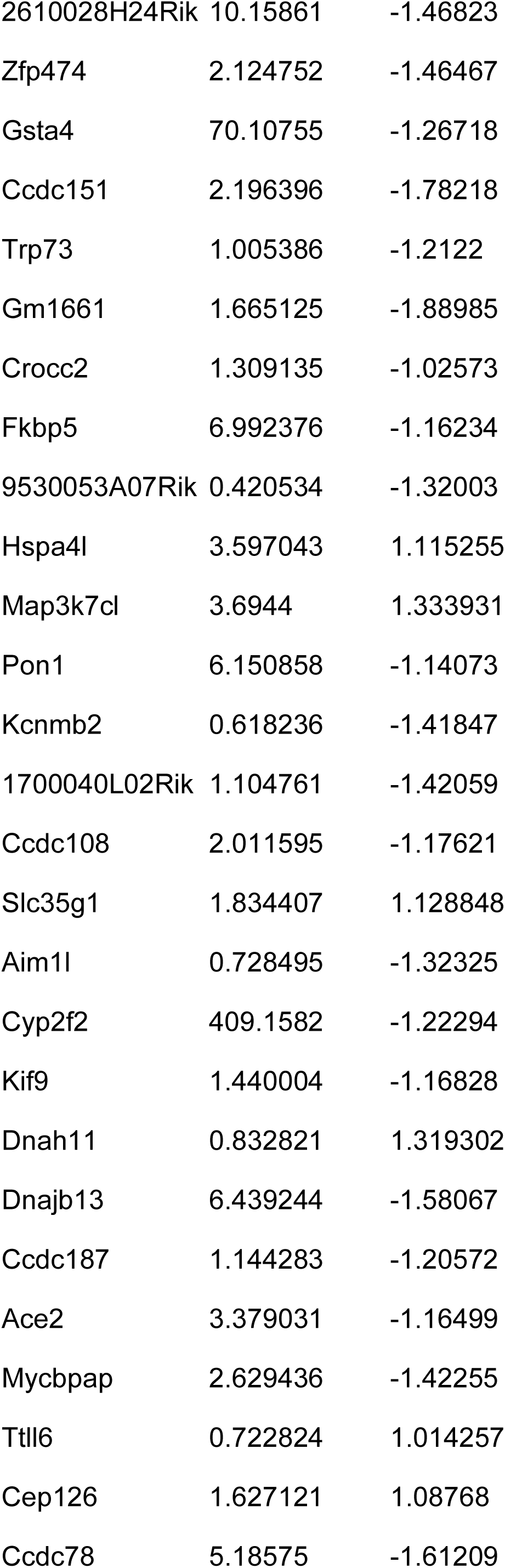
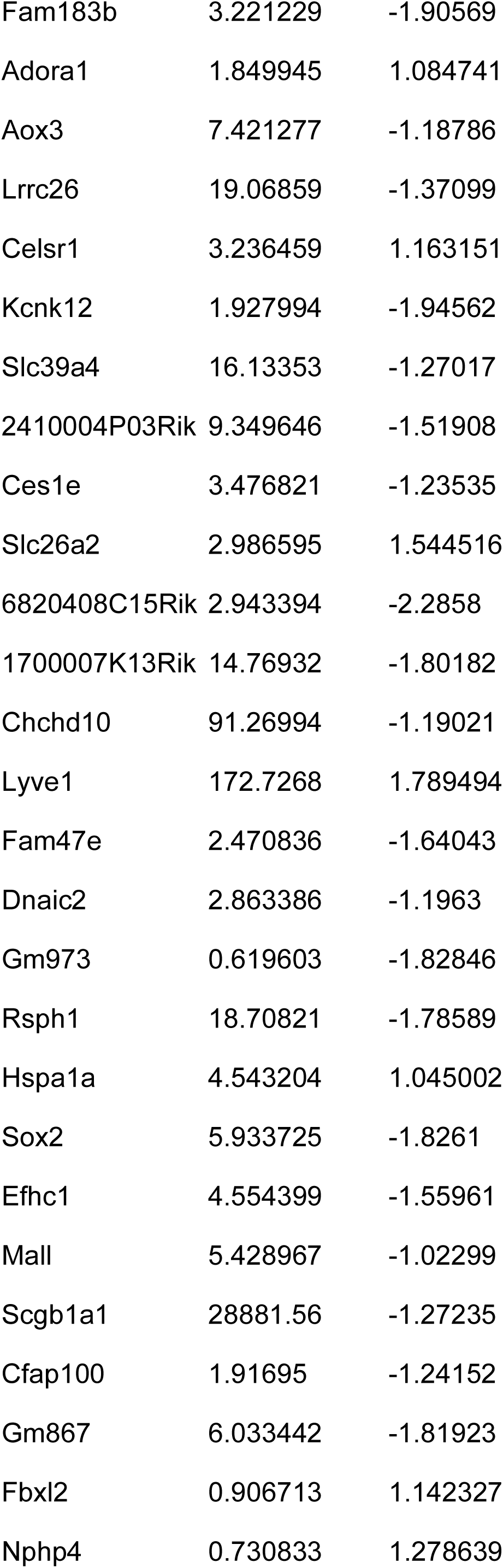
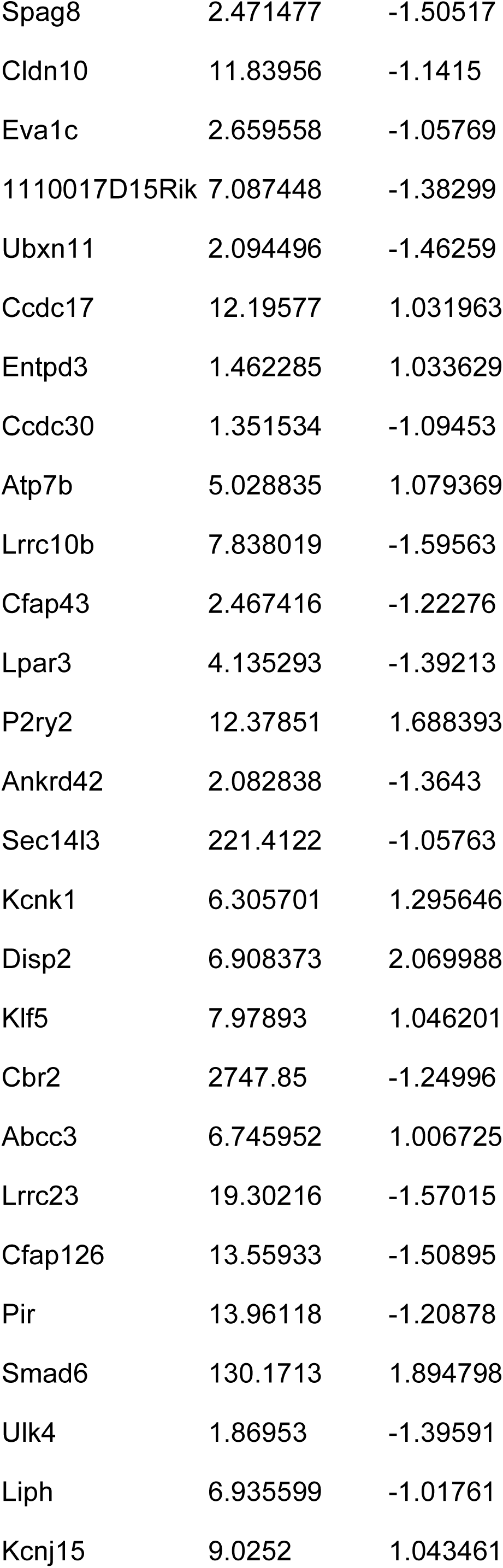
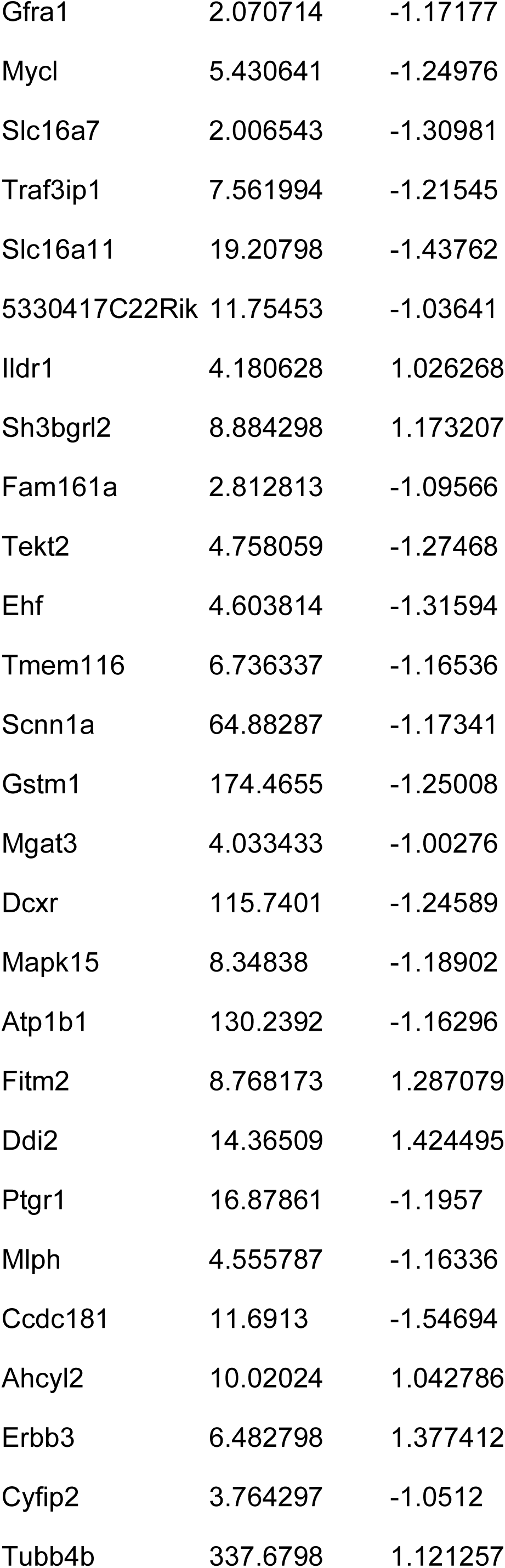
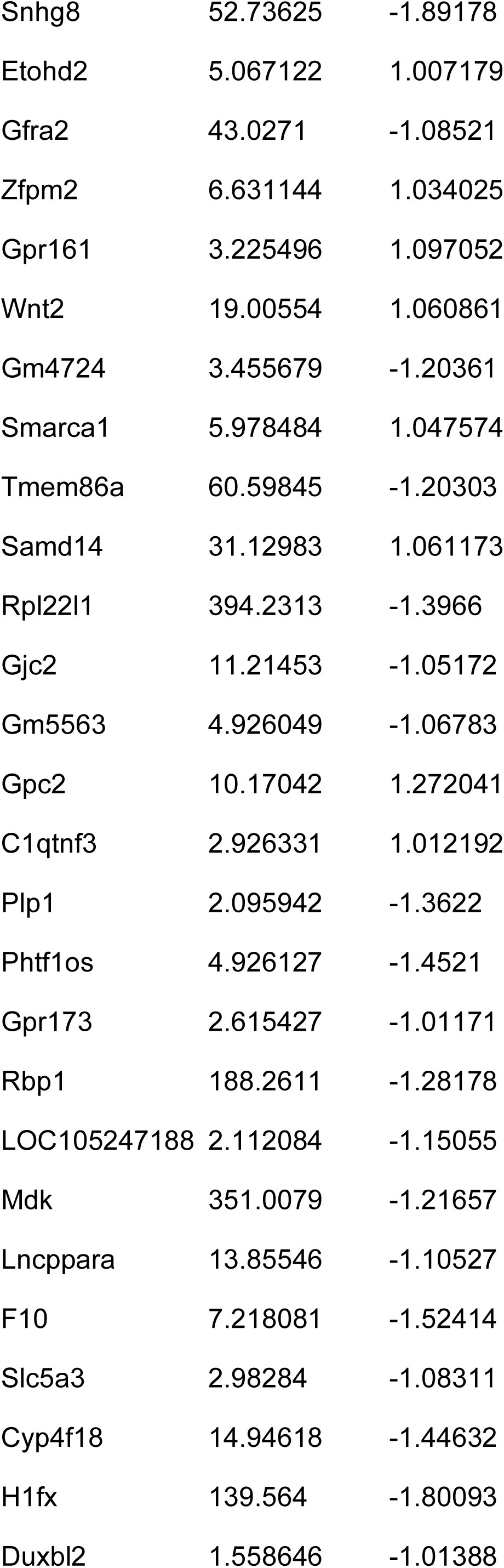
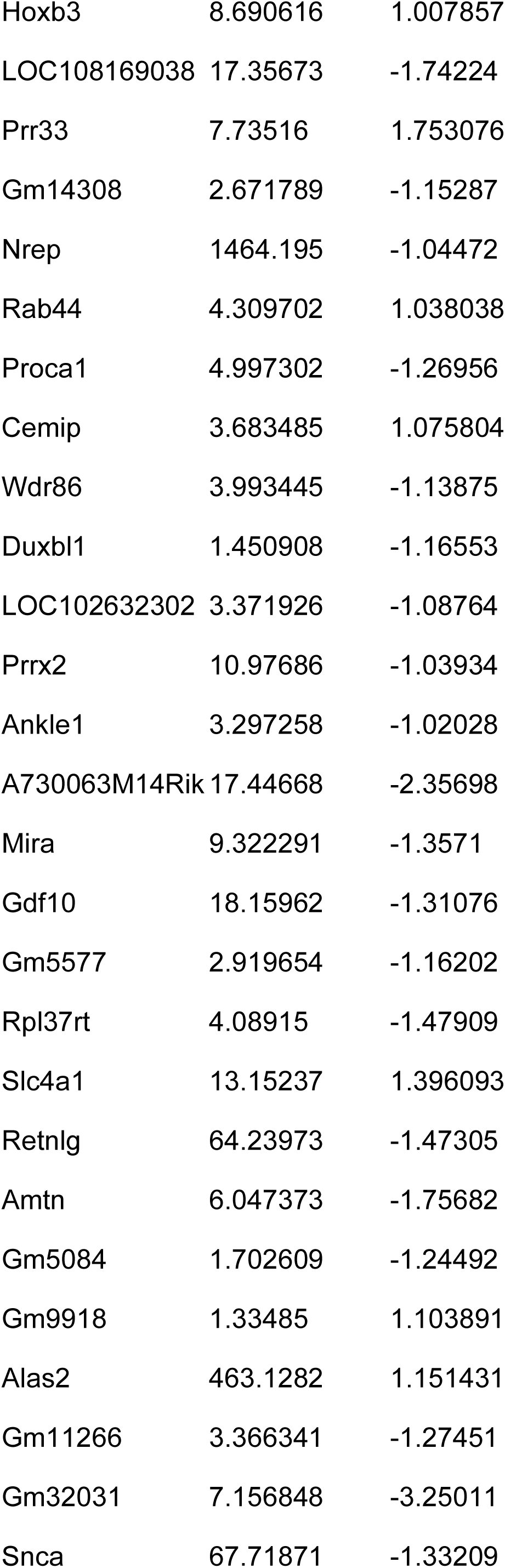
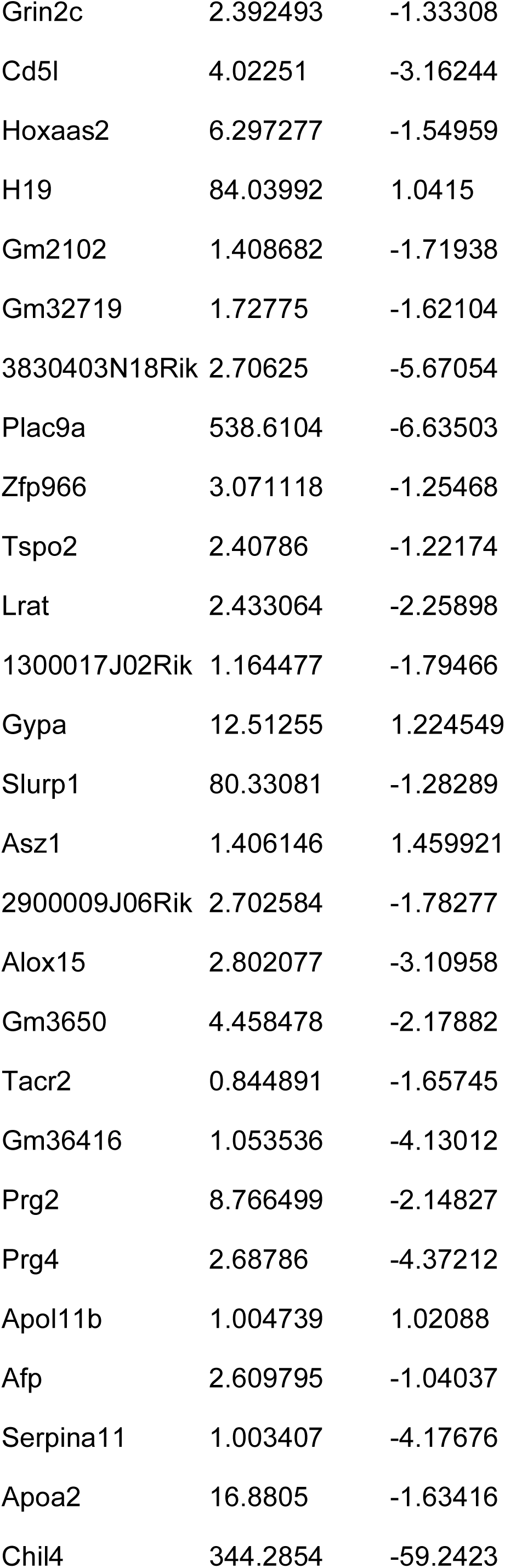
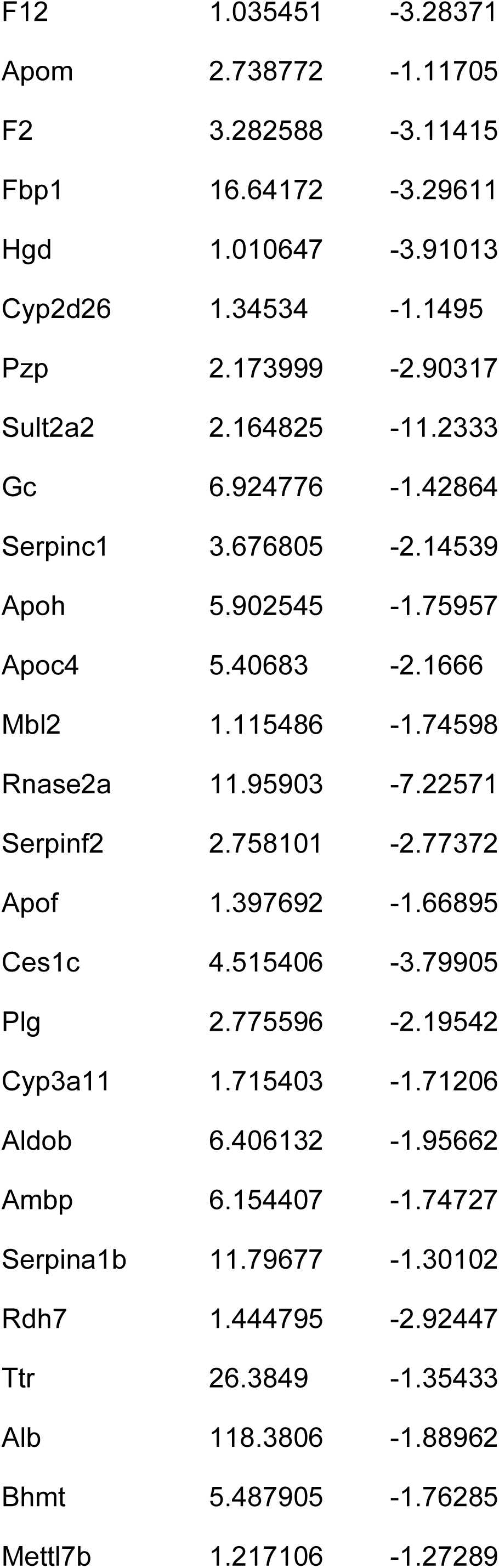
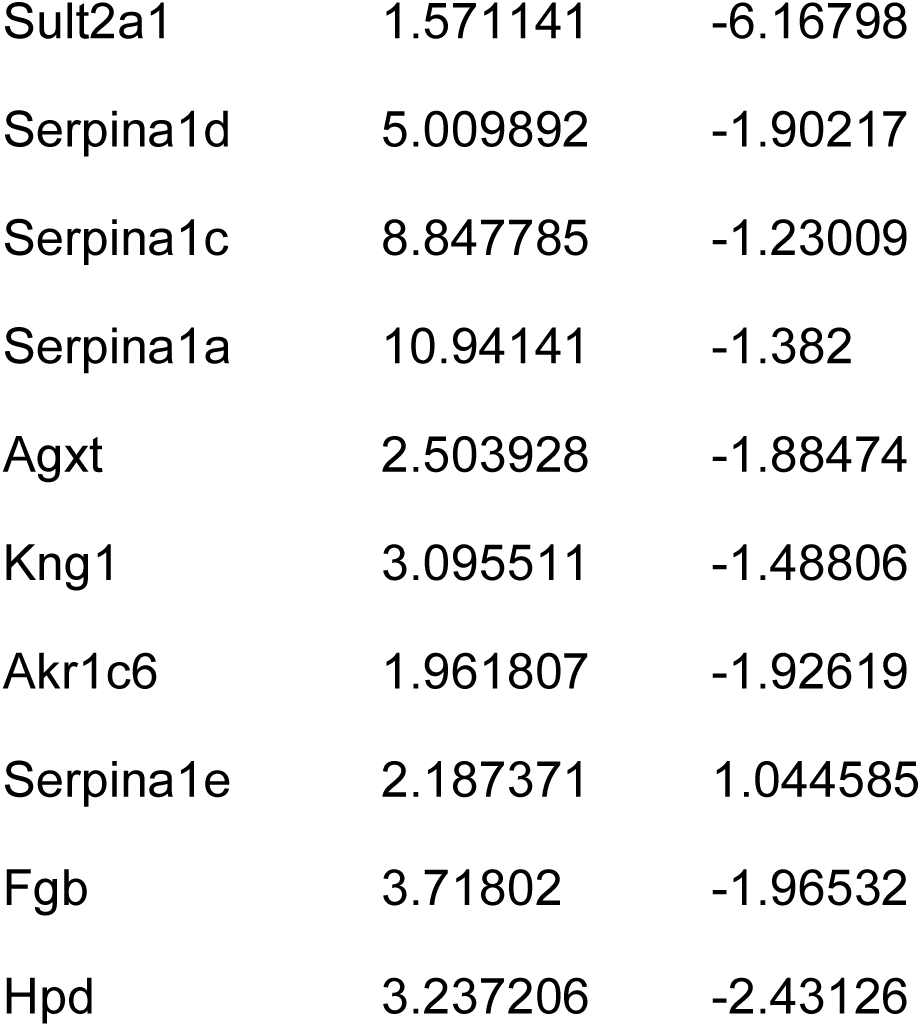
Genes uniquely regulated in the lungs of NoBPD-FMT mice as compared to conventional specific pathogen-free controls.

